# Assessing and improving the validity of COVID-19 autopsy studies - a multicenter approach to establish essential standards for immunohistochemical and ultrastructural analyses

**DOI:** 10.1101/2022.01.13.22269205

**Authors:** S Krasemann, C Dittmayer, S v. Stillfried, J Meinhardt, F Heinrich, K Hartmann, S Pfefferle, E Thies, R v. Manitius, T Aschman, J Radke, A Osterloh, S Schmid, EM Buhl, J Ihlow, S Elezkurtaj, D Horst, AC Hocke, S Timm, S Bachmann, V Corman, HH Goebel, J Matschke, S Stanelle-Bertram, G Gabriel, D Seilhean, H Adle-Biassette, B Ondruschka, M Ochs, W Stenzel, FL Heppner, P Boor, H Radbruch, M Laue, M Glatzel

## Abstract

**Background:** Autopsy studies have provided valuable insights into the pathophysiology of COVID-19. Controversies remain whether the clinical presentation is due to direct organ damage by SARS-CoV-2 or secondary effects, e.g. by an overshooting immune response. SARS-CoV-2 detection in tissues by RT-qPCR and immunohistochemistry (IHC) or electron microscopy (EM) can help answer these questions, but a comprehensive evaluation of these applications is missing.

**Methods:** We assessed publications using IHC and EM for SARS-CoV-2 detection in autopsy tissues. We systematically evaluated commercially available antibodies against the SARS-CoV-2 spike protein and nucleocapsid, dsRNA, and non-structural protein Nsp3 in cultured cell lines and COVID-19 autopsy tissues. In a multicenter study, we evaluated specificity, reproducibility, and inter-observer variability of SARS-CoV-2 nucleocapsid staining. We correlated RT-qPCR viral tissue loads with semiquantitative IHC scoring. We used qualitative and quantitative EM analyses to refine criteria for ultrastructural identification of SARS-CoV-2.

**Findings:** Publications show high variability in the detection and interpretation of SARS-CoV-2 abundance in autopsy tissues by IHC or EM. In our study, we show that IHC using antibodies against SARS-CoV-2 nucleocapsid yields the highest sensitivity and specificity. We found a positive correlation between presence of viral proteins by IHC and RT-qPCR-determined SARS-CoV-2 viral RNA load (r=-0.83, p-value <0.0001). For EM, we refined criteria for virus identification and also provide recommendations for optimized sampling and analysis. 116 of 122 publications misinterpret cellular structures as virus using EM or show only insufficient data. We provide publicly accessible digitized EM and IHC sections as a reference and for training purposes.

**Interpretation:** Since detection of SARS-CoV-2 in human autopsy tissues by IHC and EM is difficult and frequently incorrect, we propose criteria for a re-evaluation of available data and guidance for further investigations of direct organ effects by SARS-CoV-2.

**Key messages:**

- Detection of SARS-CoV-2 proteins by IHC in autopsy tissues is less sensitive in comparison to SARS-CoV-2 RNA detection by RT-qPCR.
- For determination of SARS-CoV-2 protein positive cells by IHC in autopsy tissues, detection of spike protein is less sensitive than nucleocapsid protein.
- Correct identification of SARS-CoV-2 particles in human samples by EM is limited to the respiratory system.
- Interpretation of IHC and EM should follow substantiated consensus criteria to enhance accuracy.
- Existing datasets describing SARS-CoV-2 presence in human autopsy tissues need to be critically re-evaluated.

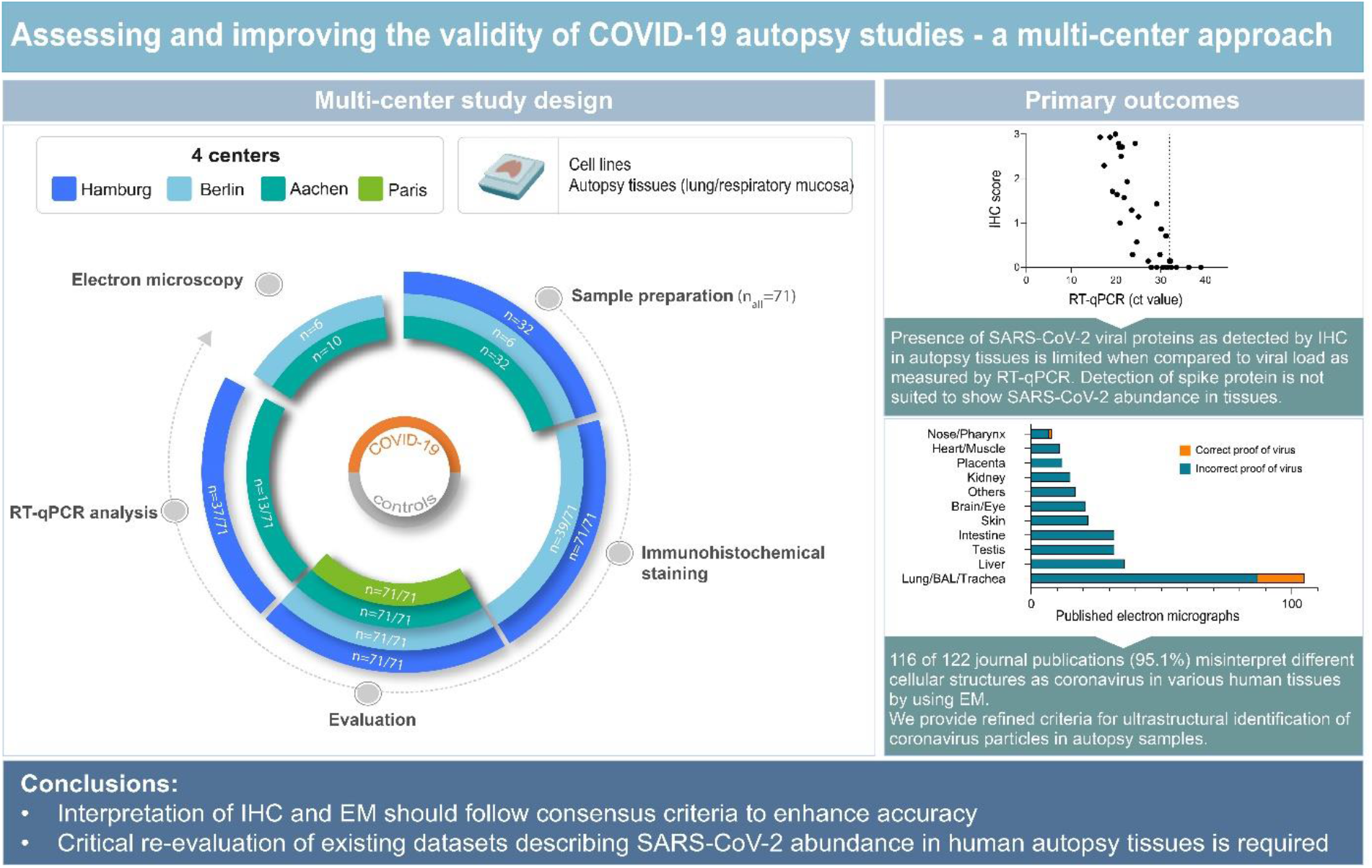

## Introduction

In clinical routine, quantitative reverse transcriptase (RT)-qPCR and rapid antigen detection tests from nasopharyngeal swabs are robust, standardized, and validated tools for screening or diagnosing severe acute respiratory syndrome coronavirus 2 (SARS-CoV-2) infections. In contrast, in situ SARS-CoV-2 detection methods such as immunohistochemistry (IHC) and electron microscopy (EM) in patient tissues are much less investigated and validated, yet autopsy studies use these methods to investigate mechanisms of organ damage and organ tropism of SARS-CoV-2 ^1-6^.

Detection of pathogen-specific antigens by immunohistochemical staining is a potent diagnostic tool allowing for spatial correlation of pathological changes with presence of the pathogen. At the beginning of the COVID-19 pandemic, positive controls for this newly emerging disease were not available, neither for diagnostic nor for research applications. The urgent need to publish data on the distribution of SARS-CoV-2 in tissues of deceased patients with mentioned technical limitations led to inconsistencies in interpretation of SARS-CoV-2 localization and distribution by IHC and EM ^1-10^.

Diagnostic EM is the only technique to directly visualize and detect intact SARS-CoV-2 particles ^11,12^. It thereby validates other in situ viral-detection techniques, detecting viral proteins or RNA, and enables cellular and subcellular localization of virus particles ^11^. EM of virus molecules or particles in model systems expanded our understanding of SARS-CoV-2 structural and cellular biology, yet did not provide information on the distribution of the virus in human tissues ^13-17^.

Due to complex sample processing procedures in electron microscopy and challenges in recording and interpretation of micrographs, misinterpretation of structures as SARS-CoV-2 particles in patient tissues occurred ^18-20^. In autopsy tissues, virus structures have to be distinguished from other structures of cells. For this task, sufficient structural preservation of the tissue and a suitable sampling strategy to detect infected cells are needed ^8,18,21^. Recommendations for identifying SARS-CoV-2 particles by EM are mainly based on virus particles in cell cultures ^22,23^ which only partially reflect the situation in autopsy tissues. In fact, misinterpretations or findings with insufficient evidence have still been published ^3,24^. Here we collated available data on in situ detection of SARS-CoV-2 in human tissue samples focussing on immunohistochemical detection of SARS-CoV-2 proteins and EM detection of intact SARS-CoV-2 particles. Furthermore, we determined optimally suited SARS-CoV-2 antibodies and evaluated their sensitivity and specificity in a multicenter approach, which allowed a correlation between the viral load detected by RT-qPCR and by IHC. Additionally, we refined criteria for identifying SARS-CoV-2 by IHC and EM and provide a publicly accessible repository of entirely digitized light- and electron microscopical sections showing examples and pitfalls for in situ detection of SARS-CoV-2.

## Material and methods

### Sample processing for immunohistochemistry

We generated paraffin cell blocks from SARS-CoV-2-infected and un-infected Vero cells (see Supplementary Methods for details) and processed these as if they were autopsy samples regarding fixation time in 10% formalin and paraffin embedding. Furthermore, SARS-CoV-2-infected and un-infected Vero cells were grown on coverslips and fixed in 4% formaldehyde solution. Autopsy samples from lungs and respiratory mucosa from COVID-19 patients and controls were formalin-fixed and paraffin-embedded (FFPE) using standard laboratory procedures from three study centers. Beside hematoxylin and eosin staining, immunohistochemistry with several antibodies against different SARS-CoV-2 proteins (see Supplementary Table 2) were performed using a Ventana Benchmark XT autostainer at two centers. For the multicenter study evaluating IHC to SARS-CoV-2 nucleocapsid, we used antibody N#9. Staining specificity and intensity were evaluated by eight independent pathologists/neuropathologists at four different centers (Pathology Aachen, Neuropathology Berlin, Neuropathology Hamburg, Pathology Paris). Observers received instructions and a set of stained slides and were then asked to categorize each slide in a blinded fashion, using a published four-tiered semiquantitative approach (none (0), slight (+), moderate (++), and severe (+++) ^25^ (Figure 3). Training slides were annotated, uploaded, and are publicly available using OMERO ^26^. Correlation of RT-qPCR (Pearson r) values and scoring was analyzed using Graphpad Prism (GraphPad Software Version 8, La Jolla, USA) with 34 pairs. Every case was rated by seven investigators. Cases with ≥70% agreement amongst raters (at least 5 from 7 raters) were classified as positive (“+” or more) or negative, respectively. Cases with unclear classification into positive or negative due to inconsistent ratings (less than 60% agreement) were defined as non-classifiable (n=2; one control / one COVID-19). Sensitivity was calculated as *Sensitivity = True positive/True positive + False negative*. Specificity was calculated as *Specificity = True negative/True negative + False positive* (see Supplementary Table 5).

### Sample processing for electron microscopy

29 autopsy samples derived from different tissues of 16 patients and SARS-CoV-2-infected Vero cells were processed according to standard protocols (Supplementary Methods; Supplementary Table 6). For qualitative and quantitative analyses of infected cells and viral particles, autopsy lung tissue (Supplementary Table 6, case 1) and FFPE re-embedded olfactory mucosa (case 2) with a high viral RNA load, as determined by RT-qPCR, and successful detection of particles by EM were used.

### Large-scale electron microscopy and transmission electron microscopy

Four large sections prepared from different resin blocks of two autopsy lung samples (case 1) and two sections prepared from different resin blocks from SARS-CoV-2-infected Vero cells were completely digitized at 3-4 nm pixel size as recently described ^27^ to screen for SARS-CoV-2 particles (Supplementary Material). The screening datasets were processed via Fiji software/TrakEM2 plugin ^28^ and nip2 software to generate high-resolution tif files ^27^ for in-depth analysis using QuPath software 0.2.0. ^29^. In the autopsy lung, in total, 15 infected cells were detected, all located within two of the four digitized sections (dataset 1 and 2 on www.nanotomy.org). Subsequently, the infected cells were annotated and digitized at 1 nm pixel size for quantitative analysis in QuPath. Selected regions showing extracellular particles and non-infected cells were also digitized. To assess the heterogeneity of coronavirus (CoV) particles and their distribution in infected cells, four different categories of particles were specified; dark type 1 particles, bright type 2 particles, deformed type 3 particles, and extracellular type 4 particles (Figure 5D-S). These types were counted manually in QuPath. Also, total area (in μm^2^) of infected cells, area of the nucleus, and area of luminal structures were measured with the polygon tool to determine the density of intracellular CoV particles per μm^2^ cytoplasmic area. Intracellular CoV particles were analysed for particle diameter (largest diameter without spikes), ribonucleoprotein (RNP) diameter (smallest diameter, granular or elongated profiles), and diameter of tubular structures (cells 2 and 15). Measurements were performed with the line annotation tool, analysis was done in Excel (Office Home and Student 2019; Microsoft Corporation, Redmond, USA). Average, standard deviation, minimal and maximal values were determined for CoV diameter, RNP diameter, and tubular structure diameter. No measurements were excluded during data processing.

In FFPE re-embedded olfactory mucosa (case 2), we restricted quantitative analysis to measurement of virus particle diameter due to limited preservation. The maximal diameter of intracellular particles (without spikes), based on TEM images, was measured using Fiji ^30^ and the “straight line” tool.

We roughly compared viral RNA load measured by RT-qPCR with virus particle number per section and cell to estimate the likelihood of finding CoV particles via thin section EM (Supplementary Methods).

### Search and analysis of publications demonstrating immunohistochemical and/or ultrastructural detection of SARS-CoV-2

We defined specific search strategies to find scientific publications using immunohistochemical datasets from autopsy studies with antibodies to SARS-CoV-2 proteins. Additionally, we searched for all scientific publications claiming ultrastructural proof of SARS-CoV-2 particles in infected human tissues (see Supplementary Methods).

## Results

### Analysis of publications demonstrating immunohistochemical detection of SARS-CoV-2 in human samples

We curated papers with immunohistochemical datasets for inclusion into our overview table (Supplementary Table 1). Detection of SARS-CoV-2 spike and nucleocapsid by immunohistochemistry in autopsy tissues showed differences regarding virus protein amounts and interpretation of data. Furthermore, results of adequate positive and negative controls were rarely reported (Supplementary Table 1).

### Assessing commercially available antibodies against SARS-CoV-2 proteins

Of the 13 commercially available anti-SARS-CoV-2 protein antibodies that were evaluated in this study, three target the spike protein, two target non-structural protein 3 (Nsp3), seven target the nucleocapsid protein, and one targets double-strand RNA (dsRNA) (Supplementary Table 2).

Firstly, we tested all antibodies on FFPE SARS-CoV-2 infected and un-infected Vero cells (Supplementary Figure 1) simultaneously processed to avoid batch effects. Of the three different spike-protein targeting antibodies, one gave a specific signal with no background, one showed signal, but also produced nonspecific staining. Of note, one widely used antibody (#3) did not produce a signal at all. Both antibodies against Nsp3 generated nonspecific high background staining in un-infected cells. An antibody against dsRNA generated a faint signal in infected cells. Of the seven antibodies detecting SARS-CoV-2 nucleocapsid, five produced specific staining on infected cells with minimal background in un-infected cells. One widely used antibody showed high nonspecific background in un-infected cells (#12), while one antibody did not produce any signal (#13) (Supplementary Figure 1).

To further investigate the specificity of antibodies that have been widely used before in research, but did not perform well in our FFPE cell-based evaluation, we infected Vero cells with SARS-CoV-2, fixed them in formalin, and stained them without prior embedding into paraffin. To determine viral protein distribution, cells were double-stained with one of the specific nucleocapsid antibodies (#7 or #9) and one of the antibodies in question against spike #3, Nsp3 #5, dsRNA #6, and nucleocapsid #12, respectively (Supplementary Figure 2). Un-infected cells served as control. While antibodies #5, #6, and #12 produced specific staining, antibody #3 did not produce a detectable signal again (Supplementary Figure 2). Since antibodies #4 and #13 did not produce specific signals in FFPE cell blocks or fixed cells, they were not further considered.

To assess the performance of anti-SARS-CoV-2 antibodies in COVID-19 autopsy tissues, we tested nine of the antibodies on FFPE autopsy tissues from COVID-19 patients and controls. We chose lung and respiratory mucosa as these tissue compartments harbor high SARS-CoV-2 viral loads and we stained consecutive sections with our panel of antibodies (Figure 1; Supplementary Figure 3). Again, two out of three anti-spike antibodies produced specific staining, while one gave no signal (#3). Of the six different nucleocapsid antibodies, five antibodies showed specific and robust staining. However, the background staining in COVID-19-positive tissues was higher in the polyclonal (#7) and one monoclonal (#11) compared to the other monoclonal antibodies tested in our study. Background staining caused by presence of SARS-CoV-2 proteins in cellular debris or tissue artifacts was higher in lung tissue when compared to that of respiratory mucosa of COVID-19 patients (Figure 1; Supplementary Figure 3). Interestingly, signals obtained with antibodies targeting nucleocapsid protein were more abundant in tissues when compared to signals obtained using anti-SARS-CoV-2 spike protein antibodies (Figure 1; Supplementary Figure 3). To evaluate this further we performed double immunofluorescence staining on Vero cells, lung, and respiratory mucosa and consistently observed more nucleocapsid positive cells than spike-protein positive cells. Moreover, nucleocapsid signals were more abundant than spike-protein within double-positive cells and they are evenly distributed throughout individual infected cells (Figure 2). To exclude an antibody-specific effect caused by spike antibody targeting spike 2 (#1), we evaluated several anti-spike antibodies. However, reduced abundance of spike SARS-CoV-2 in contrast to nucleocapsid in cells and tissues could also be confirmed using antibodies against spike S1 and receptor-binding domain (RBD) (Supplementary Figure 4).

**Figure 1:**
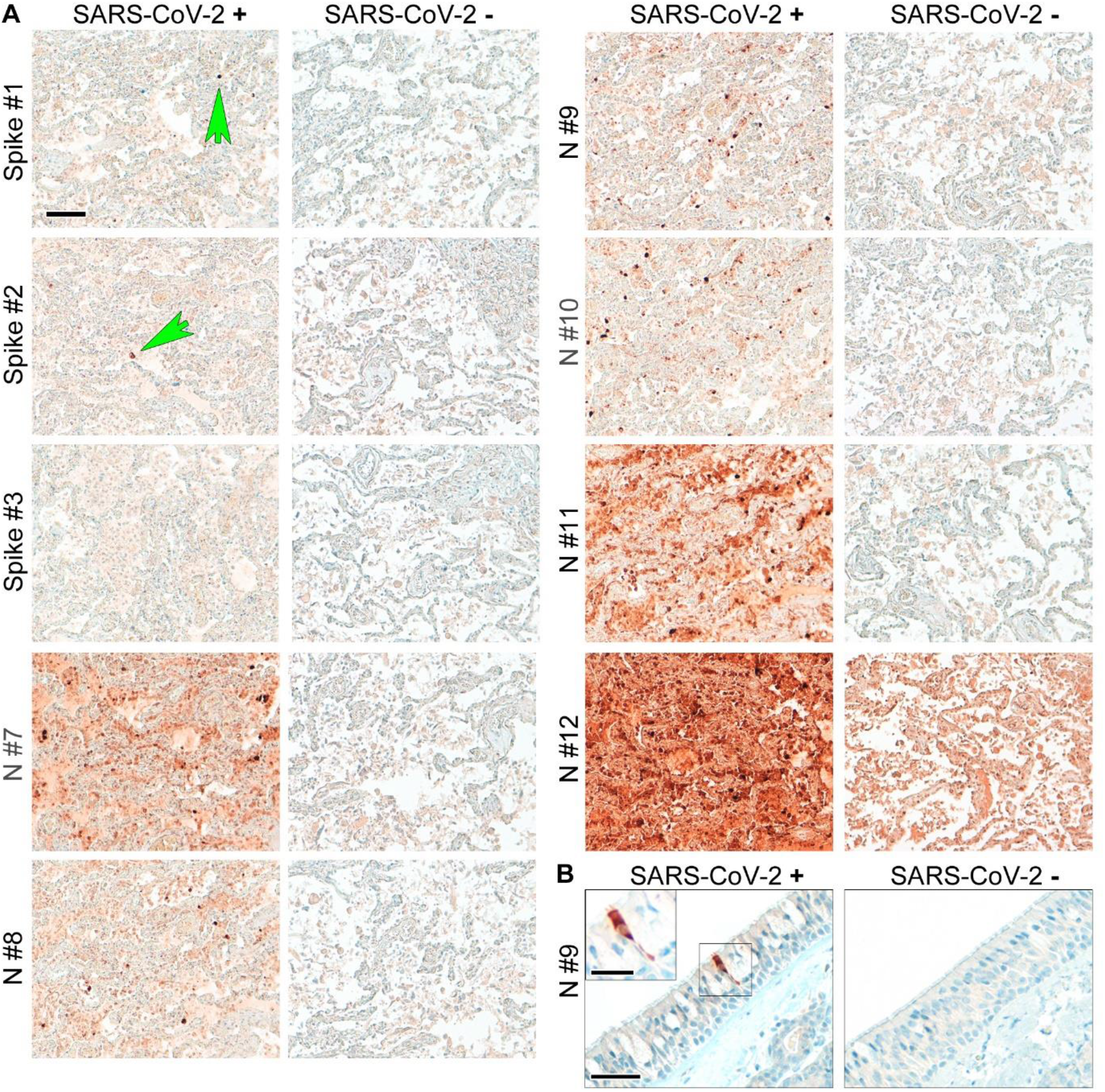
Suitable antibodies for SARS-CoV-2 protein detection show an optimal signal to background ratio in human autopsy tissues. (A) Formalin-fixed paraffin-embedded lung tissues of fatal COVID-19 and non-COVID-control were stained with different anti-spike (spike) and anti-nucleocapsid (N) antibodies (see Supplementary Table 2). Consecutive sections were used to perform the staining to enable comparability of the different antibodies. Green arrows point to single spike-protein positive cells. One widely used antibody (Spike #3) did not detect spike protein in SARS-CoV-2 positive lung tissue. Moreover, one other widely used antibody (N#12) produced very high background staining in non-COVID-control lung tissue. Of note, abundance of spike protein is much lower than that of nucleocapsid. Representative images are shown; scale bar: 100 μm. (B) Formalin-fixed paraffin-embedded respiratory mucosa from two COVID-19 patients were stained for SARS-CoV-2 nucleocapsid (antibody #9). The SARS-CoV-2-positive cell is evenly stained by the antibody and does not show single dots; scale bar: 50 μm, close-up: 25 μm.

**Figure 2:**
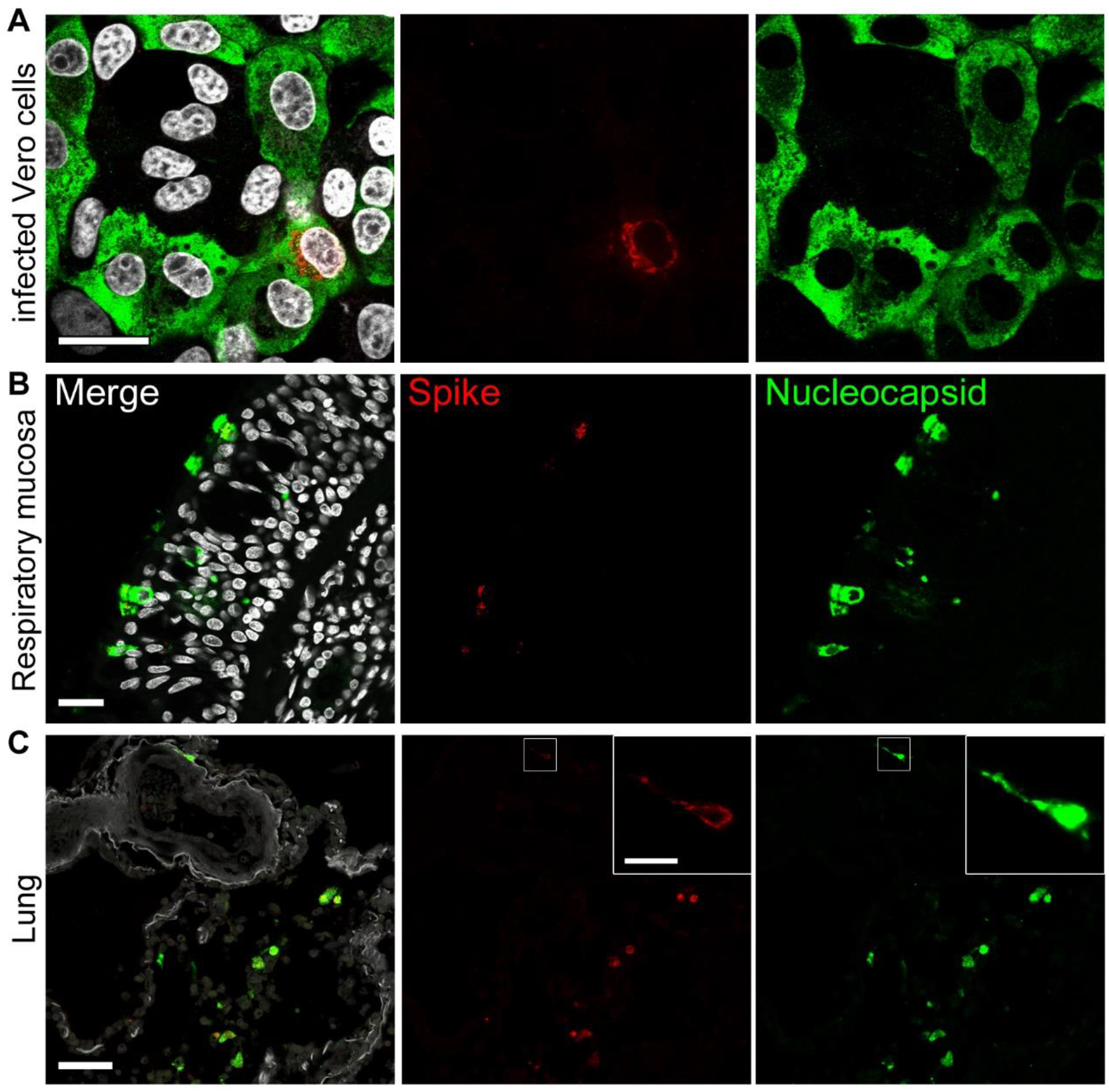
Higher abundance of nucleocapsid in comparison to spike in infected cells and tissues from COVID-19 patients. (A) Vero cells were infected with SARS-CoV-2 (MOI 0.5; 24 h), fixated in formalin, and directly stained. Representative staining of SARS-CoV-2 spike (red; antibody spike#1) and nucleocapsid (green; antibody N#7); nuclei/DAPI in white; scale bar: 25 μm. (B) FFPE sections of respiratory mucosa of a COVID-19 patient; scale bar: 25 μm or (C) lung tissue of a COVID-19 patient were stained as above for spike (red), nucleocapsid (green), nuclei/DAPI in white; scale bar: 50 μm, close-up: 10 μm. Note that the signal strength and abundance of spike protein and spike-positive cells is always less than that for nucleocapsid. Thus, we recommend using antibodies against SARS-CoV-2 nucleocapsid for the detection of SARS-CoV-2 protein positive cells in human autopsy tissues.

Of note, two widely used and published antibodies did not perform well in our analyses. One antibody (anti-N, #12) produced unacceptably high background staining even in control tissues (Figure 1; Supplementary Figures 1,3), while another widely used antibody (anti-spike, #3) did not result in measurable staining in SARS-CoV-2-infected Vero cells or COVID-19 tissues (Figure 1; Supplementary Figures 1-3). In summary, six anti-SARS-CoV-2 protein antibodies reliably work on FFPE autopsy tissues from COVID-19 patients, with antibodies against nucleocapsid protein providing better results. Our recommendations for the usage of antibodies for the detection of anti-SARS-CoV-2 proteins in human autopsy tissues are summarized in Table 1 and Supplementary Table 3.

**Table 1.** Recommendations for detection of SARS-CoV-2 proteins by IHC in formalin-fixed paraffin-embedded (FFPE) human autopsy tissues

|  |  |
| --- | --- |
| 1 | Determination of virus load by RT-qPCR may give an overestimated picture of the tissue burden; SARS-CoV-2 positive cells might be comparatively rare. |
| 2 | IHC is not suited to detect low virus amounts in cells due to unfavourable signal to background ratio in human autopsy tissues; positive cells in tissues carry a rather high virus protein load (see Figures 1,3; Supplementary Figures 3,5). |
| 3 | Always use positive/negative controls (e.g. infected versus un-infected FFPE cells (see Supplementary Methods for generation of cell blocks)). If using autopsy tissues control for fixation time and degree of inflammation. |
| 4 | Some antibodies are not recommended for detection of SARS-CoV-2 in human autopsy tissues. They either produce high background in control tissue or do not stain specifically for SARS-CoV-2 proteins (compare Figure 1; Supplementary Figures 1,3). |
| 5 | Home-made or novel antibodies need to be evaluated using FFPE cell blocks of SARS-CoV-2 infected versus un-infected cells and appropriate positive and negative control tissues. To improve comparability of different COVID-19 datasets, we recommend using one of the established antibodies in addition to the potential new one. |
| 6 | Consult an experienced pathologist to avoid misinterpretation of typical tissue artefacts (e.g. lipofuscin in neurons; carbon deposition in lung; formalin-induced artefacts). Use polarized light to identify formalin-induced artefacts in FFPE tissues (e.g. punctate or dark precipitates). |
| 7 | Nucleocapsid has a higher abundance in virus-protein positive cells, thus, usage of anti-nucleocapsid antibodies is recommended to increase the sensitivity of detection (see Figure 2; Supplementary Figure 4). Anti-spike antibodies are not recommended for detection, but are suitable as additional tools to confirm specificity in double stainings. |
| 8 | COVID-19 tissues often present with high inflammatory changes which are prone to produce higher background staining. Keep in mind that more (nonspecific) signal in tissues could be infection specific, but might not be a SARS-CoV-2 virus protein staining. It is highly recommended to employ a secondary antibody only control or isotype control in IHC. |
| 9 | We recommend to evaluate staining results on a microscope (possibility to focus in z-plane) and not on a scanned image to avoid misinterpretation of nonspecific staining artefacts on the tissue surface. |
| 10 | Some autopsy tissues provide exceptional high background staining such as kidney or placenta and should only be validated together with RT-qPCR results and comparable positive and negative tissues. |
| 11 | Nucleocapsid staining is planar and intracellular, but does not produce single punctuate dots. |
| 12 | Fluorescence microscopy is more prone to background signal than chromogenic IHC due to autofluorescence in human autopsy tissues. Check tissues in several channels to exclude autofluorescent "dots". |

### A multicenter study assessing SARS-CoV-2 immunohistochemistry

We selected one monoclonal antibody against nucleocapsid (#9) which showed a very reliable signal in cells and human autopsy tissues with minimal background staining (Figure 1A,B; Supplementary Figures 1,3), to perform a blinded multi-centric study. The goals were (1) to assess, whether IHC staining in human lung autopsy tissue by experienced pathologists is a suitable method to detect SARS-CoV-2 protein and (2) to investigate the correlation between SARS-CoV-2 load as measured by RT-qPCR and IHC (Figure 3).

**Figure 3:**
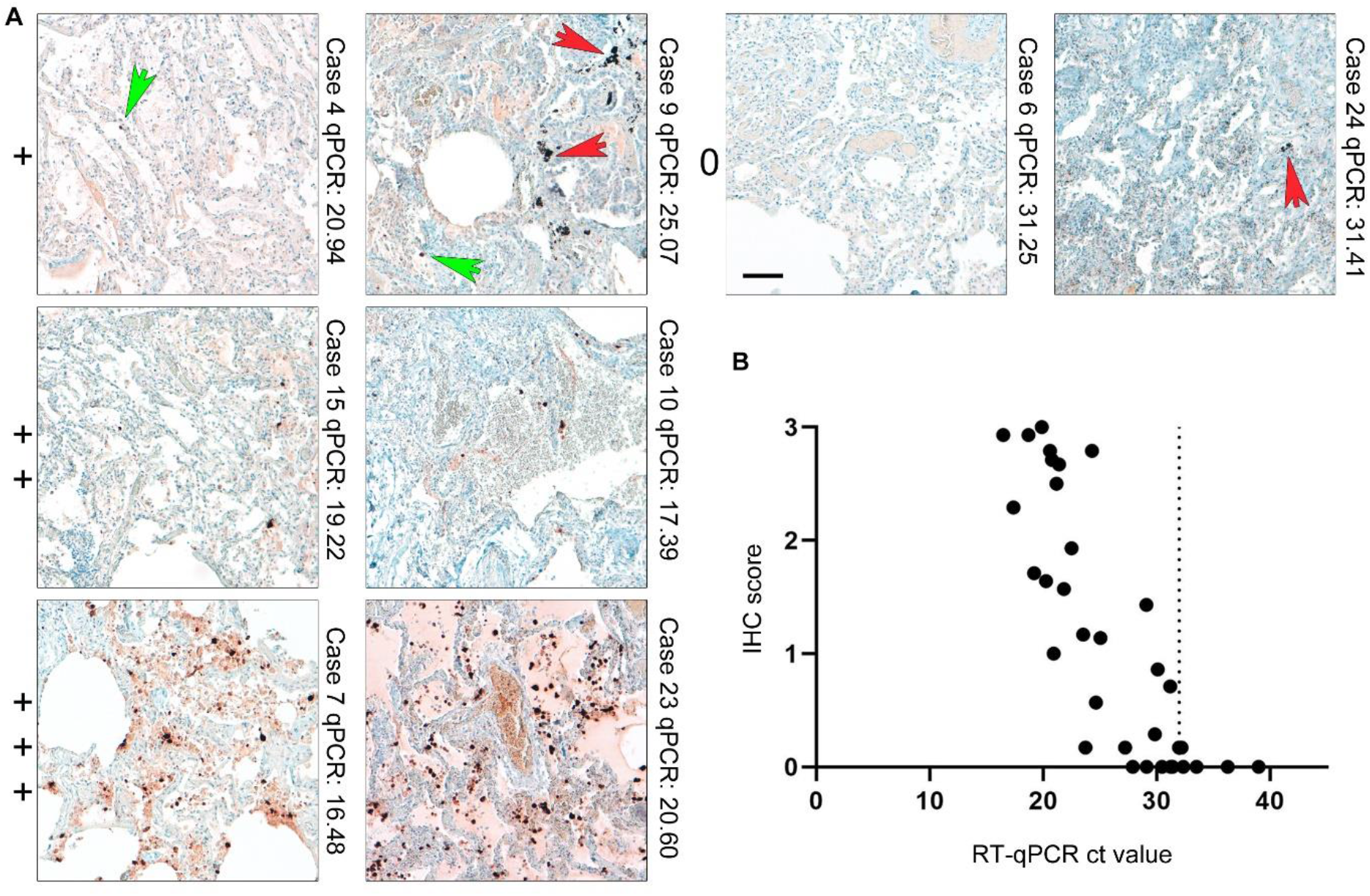
IHC for SARS-CoV-2 nucleocapsid correlates with SARS-CoV-2 viral RNA load determined by RT-qPCR. (A) Lung tissues of COVID-19 patients and respective controls were stained with an antibody against SARS-CoV-2 nucleocapsid (#9) and scored in a blinded fashion by pathologists from four different centres (for overview of patient details see Supplementary Tables 4a-c; for scoring results see Supplementary Table 5). We defined four categories for the scoring of SARS-CoV-2 nucleocapsid abundance: 0= no detection; += detection of single and/or regionally separated positive cells; ++= several positive cells and/or cluster of cells in a regionally restricted manner; +++= high abundance of positive cells and/or several highly positive cluster. Representative images of lung tissues for all four scoring categories are shown together with the case number and the RT-qPCR value. Green arrows point towards single positive cells; red arrows mark typical pigmentation due to formalin or anthracosis as further pitfalls for IHC interpretation in lung tissue. Green arrows point to nucleocapsid positive signal, while red arrow point to background typical for lung tissue. Scale bar: 100 μm. (B) SARS-CoV-2 viral RNA load was determined by RT-qPCR of consecutive tissue block paraffin sections. Viral RNA loads correlate with detection of SARS-CoV-2 nucleocapsid by IHC (r= -0.83, p-value <0.0001). Of note, widespread detection (score+++) of nucleocapsid in lung tissue is only associated with high RNA loads/low ct values, whereas at high ct values (low RNA) detection of positive cells is comparatively low.

Three centers contributed human autopsy lung tissues from COVID-19 deceased individuals which were stained against nucleocapsid. Anonymized patient details are summarized in Supplementary Tables 4a-c. Abundance of SARS-CoV-2 nucleocapsid in lung tissue in COVID-19 patients is highly variable and present in a clustered, inhomogenous pattern (Figure 3; Supplementary Figure 5). SARS-CoV-2 viral loads were determined by RT-qPCR of consecutive sections of the same paraffin blocks of the stained tissue samples (Supplementary Table 5). Lung tissues from control patients without pathological lung changes, from patients dying of non-COVID-19 related Acute Respiratory Distress Syndrome (ARDS), and from patients dying with Influenza infections were included since lung tissue intrinsically presents with high background staining (Figure 5; patients in Supplementary Tables 4a-c). Slides were evaluated blinded by pathologists from four different centers and scored in a semiquantitative manner (see Figure 3 for overview and examples; Supplementary Table 5).

We assessed false positive and false negative ratings in lung tissues. The utilized monoclonal antibody (#9) had a sensitivity of 0.71 (Supplementary Table 5). Extensive tissue damage seen in COVID-19 lungs, where pre-necrotic epithelial cells and hyaline membranes obscured interpretation of signals. The specificity for antibody #9 was high at 0.98 (40 True negative/ 40 True negative + 1 False positive; see Supplementary Table 5). A correlation between RT-qPCR-defined SARS-CoV-2 viral loads and the presence of immunohistochemically detected SARS-CoV-2 nucleocapsid protein was seen in tissues with high SARS-CoV-2 viral loads (Figure 3). COVID-19 tissues with low viral RNA loads were often rated false negative (n=7 with a mean ct value of 28.7) which could be true negative as, in these cases with only a low RNA signal there is probably no SARS-CoV-2 protein, however, in our calculations these figure as false negative. Only two cases out of 71 were not classifiable with an interrater agreement of less than 60%. The overall interrater reliability documented as interrater agreement frequency was 62% and reliability increased up to 83% if only trained raters with experience in SARS-CoV-2 IHC were included (Supplementary Table 5) ^31^. The majority of cases with interrater discrepancy (all raters) were controls (n=14) compared to COVID-19 (n=9), and almost always these cases were rated with single positive cells (Controls and COVID-19 cases). In contrast, COVID-19 cases with a high viral RNA load and a high IHC score were classified correctly.

Thus, our multicenter study showed that detection of SARS-CoV-2 proteins in human autopsy tissues is feasible yet is tainted with technical and interpretational difficulties, highlighting the importance of proper controls in autopsy studies and training of evaluators.

### Ultrastructural analysis of SARS-CoV-2 in human tissues

Guidance for identifying SARS-CoV-2 has been provided previously, however without detailed consideration of the specific challenges of autopsy tissues ^20,22,23^. Key elements guiding ultrastructural identification of SARS-CoV-2 include the presence of a membrane envelope, surface projections (spikes), a granular interior (ribonucleoprotein), and diameters from 60 to 140 nm ^20^. We searched for SARS-CoV-2 viral particles in the lung, olfactory mucosa (removed directly under the lamina cribrosa, but ultrastructurally only parts with ciliated and goblet cells were identifiable), medulla oblongata, kidney, trachea, and myocardium in autopsy tissues of 16 RT-qPCR-positive COVID-19 patients (Supplementary Table 6).

Overall, virus particles could be identified in 15 cells within the large-scale screening datasets of lung tissue of one patient. All virus-containing cells were found in two of the four digitized sections. Due to simplicity, we refer to these cells as infected cells, albeit intracellular virus particles in e.g. macrophages may be mainly composed of phagocytosed particles. Virus particles were well-preserved with distinct substructures (Figure 4A,B) and virus particles containing cells could be identified as type 2 pneumocytes and alveolar macrophages based on morphological criteria (Supplementary Table 7). Potential viral mimics such as swollen mitochondria, vesicles of rough endoplasmic reticulum (rER), and coated vesicles could be distinguished from viral particles (Supplementary Figure 6) ^23,32-34^. We recorded all 15 infected cells in the lung at very high resolution (Figure 5, repository datasets on www.nanotomy.org) for morphometric analyses and found 1557 intracellular and 144 extracellular viral particles. Infected cells showed virus loads ranging from 4 to 620 intracellular particles per sectioned cell profile (or 0.07 to 5.44 intracellular particles per μm^2^ cytoplasmic area, Supplementary Figure 7). Intracellular particles showed a mean diameter of 87 nm (±13 nm; n=1369; 55 to 177 nm). The RNP showed a mean diameter of 7.2 nm (± 1.6 nm; n=433; 3.6 to 13 nm). According to our quantitative analysis and calculations (see Supplementary Methods), individual cells may contain thousands (possibly up to 40,000; cell 7) of CoV particles. Based on a rough approximation from viral RNA load, we can expect 0.006 (case 5) or 500 (case 1) CoV particles/ mm^2^ ultrathin section area for low and high RNA load, respectively, which implies a huge difference in the likelihood of particle detection (see calculations in Supplementary Methods). These calculations also reflect our findings acquired by IHC analyses. No typical replication compartments such as double-membrane vesicles and budding viruses were found, most probably because of the limited structural preservation. However, four cells of the autopsy lung demonstrated peculiar tubular structures which are possibly associated with CoV infection (Supplementary Figure 8) ^32^.

**Figure 4:**
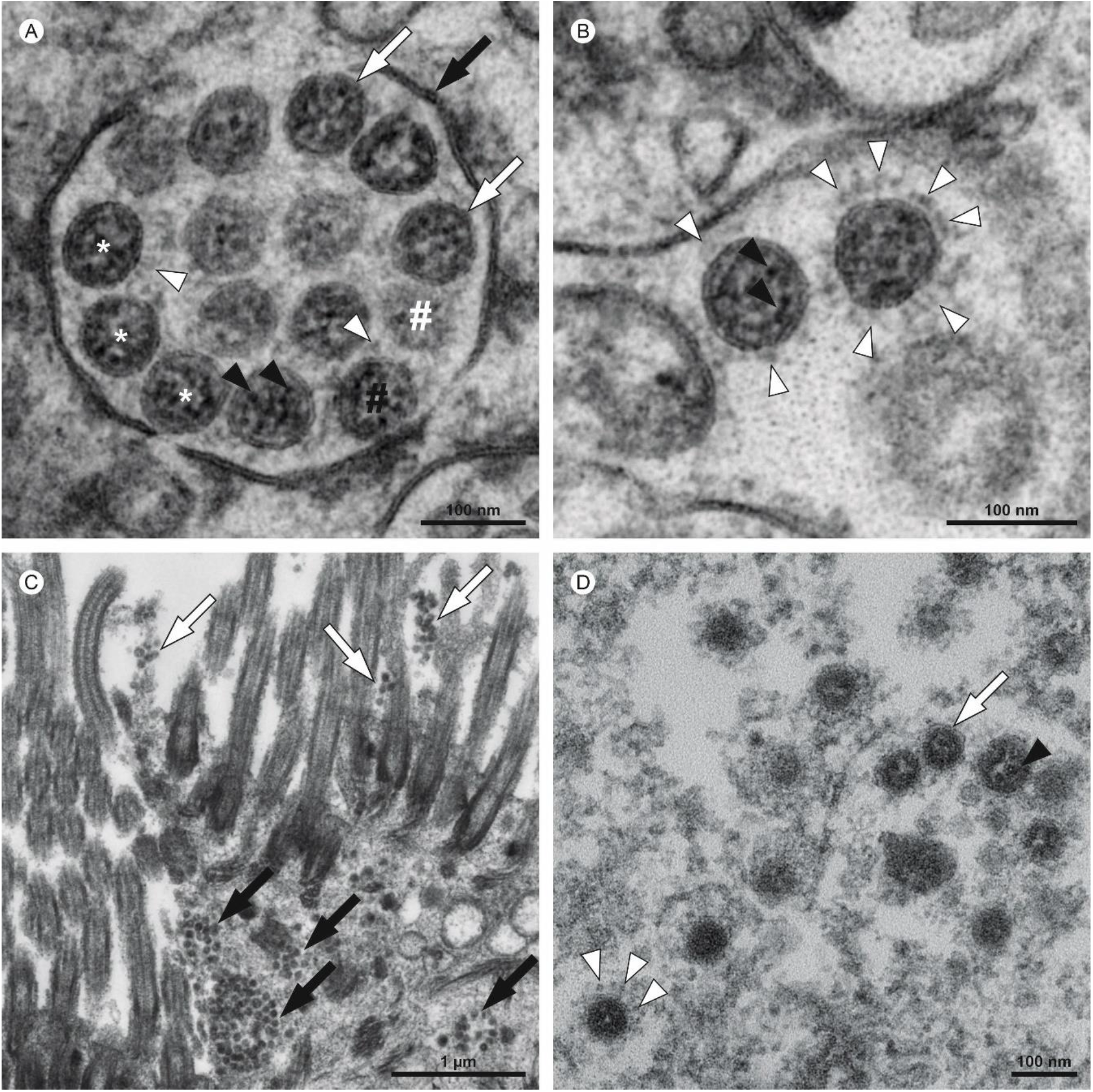
Ultrastructural characteristics of coronavirus particles in different types of autopsy samples. (A,B) Autopsy lung tissue, (C,D) FFPE-re-embedded olfactory mucosa. (A) Well-preserved intracellular coronavirus (CoV) particles (some indicated by white asterisks) in autopsy lung tissue, located within a membrane compartment (black arrow) and showing a distinct biomembrane (white arrows) and, some of them also, faint surface projections (white arrowheads). Note the granular and relatively fine dispersed ribonucleoprotein (RNP; black arrowheads). The appearance of a different electron density between the individual particles (black vs. white #) is a result of different particle volumes (about 90 nm in diameter) captured within the section volume (about 60 nm thickness); a larger particle volume within the section results in higher electron density, a phenomenon that is typical for spherical particles with an electron dense interior (as compared to e.g. empty vesicles). (B) Very few particles demonstrate prominent (well-visible) surface projections, e.g. the right CoV particle (white arrowheads) as compared to the left CoV particle. The two particles show the heterogeneous nature of the RNP in such preparations, with distinct granular and some elongated profiles (black arrowheads) in the left particle and a more granular luminal matrix with few distinct profiles in the right particle. (C) Infected cells within the olfactory mucosa also show membrane compartments (black arrows) with numerous coronavirus particles and also grouped extracellular particles that typically adhere to kinocilia and microvilli (white arrows), but individual particles are less well-preserved and more difficult to identify than particles in A and B. Virus particles appear as groups of electron dense particles of rather uniform size. (D) Virus particles at higher magnification (another region as shown in C). The particles are surrounded by a biomembrane (white arrow) which only rarely show surface projections (white arrowheads; note the globular shape of their peripheral part and also the relatively low electron density as compared to e.g. the RNP). The granular luminal matrix, formed by the RNP, is only visible in a few particles (black arrowhead). Individual images were acquired manually at high resolution by scanning transmission electron microscopy (STEM; A,B) and transmission electron microscopy (TEM; C,D).

**Figure 5:**
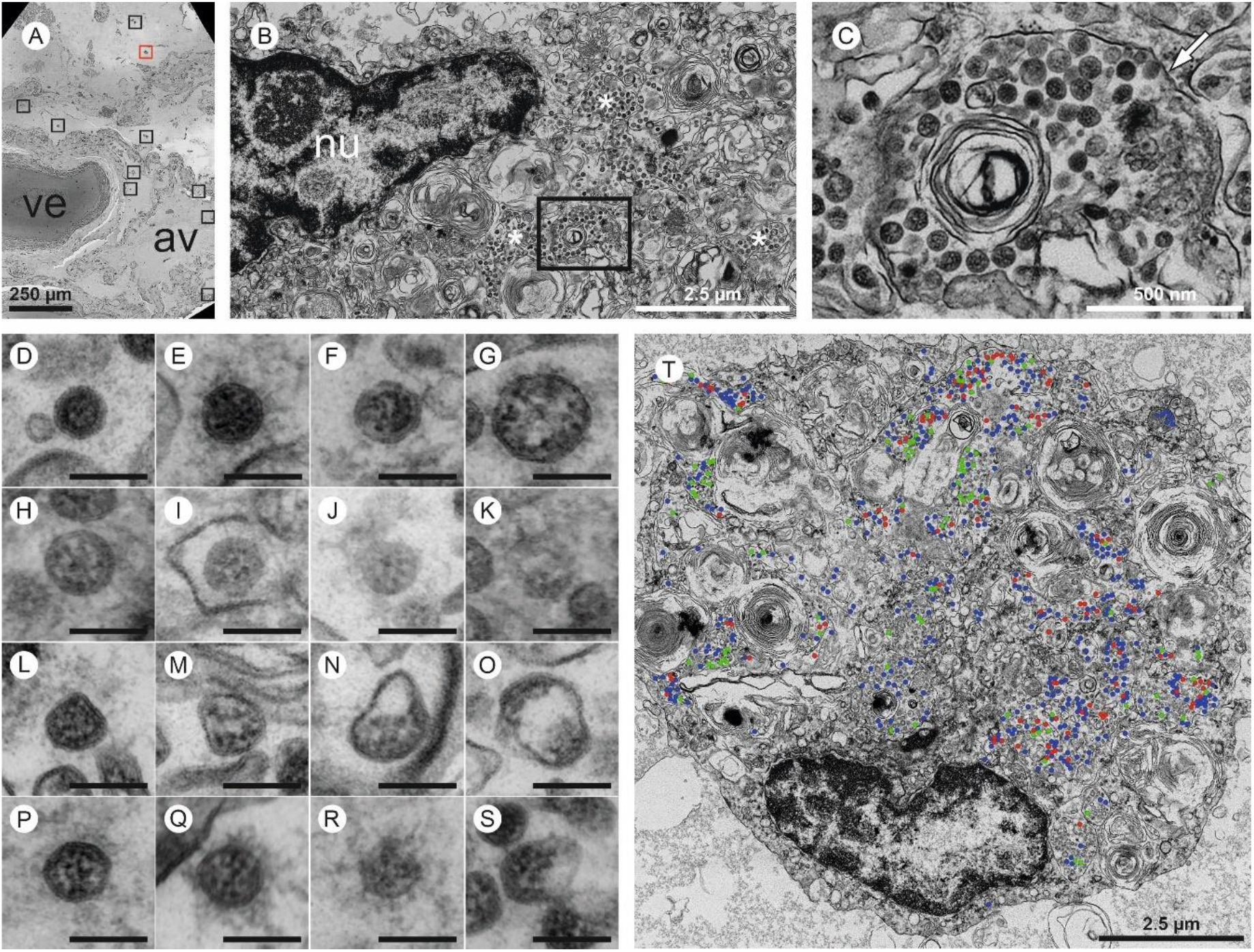
Large-scale electron microscopy of SARS-CoV-2 in human autopsy lung tissue. Four entire sections of EM case 1 were automatically digitized (dataset 1 is depicted here) at 3-4 nm pixel size to screen for SARS-CoV-2 infected cells (A-C). In the overview (A), a large vessel (ve) and an alveolus (av) are readily detectable. Preserved microanatomy allows to precisely locate regions of interest at the nanometer scale within the histological context at the millimeter scale. The red box in (A) indicates the position of the infected cell shown in (B; cell 6), the nine black boxes indicate locations of other infected cells. Numerous coronavirus (CoV) particles are located in the region next to the nucleus (nu), note several well-preserved membrane compartments with numerous CoV particles (white asterisks in B, and box in B that is further magnified and marked by the white arrow in C). The screening resolution is sufficient to detect typical CoV morphology (C) with prominent ribonucleoprotein (RNP) within the interior of particles. All infected cells were digitized at a very high resolution of 1 nm pixel size (D-S; T, cell 7) to resolve CoV substructure with improved image quality for validation and quantitative analysis. (D-S) Ultrastructural types of coronavirus particles; for quantitative analysis of CoV particles, different morphological types were defined. (D-G) Type 1 CoV particles are electron dense, corresponding to a large particle volume within the section and are relatively well preserved with a round to oval shape. Different representative appearances are shown here; small particle (D), a standard-sized particle with evenly distributed granular-appearing RNP (E), partly elongated-appearing RNP (F), a larger particle with slightly irregular interior (G). (H-K) Less electron dense type 2 CoV particles, corresponding to a smaller particle volume within the section, while being also relatively well preserved; relatively electron dense particle with well-recognizable biomembrane (H), less electron dense particles with also less distinct biomembrane (I-K). Note that the partly granular and partly elongated RNP profiles are still visible. (L-O) Type 3 CoV particles have the electron density of type 1 or type 2 CoV particles, but show more bizarre shapes. (P-S) Type 4 CoV particles were defined as all extracellular particles next to infected cells; well preserved dark particle (P), some particles showed prominent “fuzzy” coats (Q), less electron dense (R) and deformed (S) particles. (T) Visualization of different particle types in QuPath; type 1 (blue), type 2 (red) and type 3 (green). See also www.nanotomy.org for internet browser-based open access pan-and-zoom analysis of the full resolution datasets and for our Supplementary Video demonstrating how large-scale electron microscopy facilitates ultrastructural analysis and visual pattern recognition.

Virus particles could also be identified in several ciliated cells within olfactory mucosa of one patient (Supplementary Table 6; Figure 4C,D). However, virus particles were less well-preserved due to the FFPE-embedding and paraffin extraction procedure prior to re-embedding for EM. Thus, their identification relied on comparison of more particles within each cell as well as presence of typical membrane compartments with multiple isomorphic particles enclosed. Virus particles appeared more condensed than virus particles after standard preparation for thin section EM with a mean diameter of 73 nm (± 7 nm; n=175; 58 to 108 nm; intracellular particles).

Of note, no virus particles were found in 26 samples of 14 patients in different tissues (Supplementary Table 6).

Based on reliable reference publications on ultrastructural in situ detection of CoV in human samples ^21,32,35-39^, our previous work on SARS-CoV-2 in autopsy tissues ^8,18^, cell culture ^16^, MERS ^40^, and the results presented here, we developed refined criteria for identification of CoV particles in autopsy samples (Table 2).

**Table 2.** Recommendations for detection of intact SARS-CoV-2 particles using electron microscopy in human autopsy tissues

| Criteria for ultrastructural identification |  |  |
| --- | --- | --- |
| General considerations |  | It is sufficient if all of these criteria are met by a group of closely associated and similar particles within one individual cell, but individual particles of different cells should not be combined. Identification of cell types in autopsy tissues is challenging and often not possible, complicated also by pathological and virus-induced alterations that may mimic e.g. lamellar bodies of type 2 pneumocytes. |
| 1 | Shape | Round to oval. |
| 2 | Size | 50-180 nm (mean = $87 \pm 13$ nm; without spikes), with smaller particles in re-embedded FFPE material (mean = $73 \pm 7$ nm, 58-108 nm). In the range also described for cell culture <sup>16,68</sup> and autopsy lung <sup>20</sup>. |
| 3 | Membrane | At least partially visible around the particle. |
| 4 | Surface projections | Thin stalk and a globular component (in total about 20 nm long <sup>16</sup>), at least the globular component needs to be discernible with some distance from the biomembrane. The electron density is considerably lower than surface structures of e.g. coated vesicles and the particle surface usually is not entirely covered <sup>16</sup>. Note that the visibility of surface projections may be heterogeneous within the sample and also depends |
|  |  | on additives applied during tissue embedding such as *en bloc* treatment with tannic acid and uranyl acetate<sup>16,32</sup>. |
| 5 | Interior structure | Inhomogeneous granular (never empty or homogeneous at low electron density), ribonucleoprotein (RNP) profiles are round/aggregated or oval/longitudinal structures. Based on our findings, the RNP profile diameter is generally between 3.6-13 nm (mean = 7.2 nm ± 1.6 nm), as published<sup>20,23</sup>. |
| 6 | Number | Particles must be present at higher number and should often occur in groups within cells<sup>8,21,35</sup>. |
| 7 | Location | Extracellular: individual particles or small groups, sometimes attaching to outer surface of membranes. Intracellular: within small compartments with e.g. 1 particle up to very large compartments with dozens of particles, sometimes attaching to the inner surface of the membranes, but compartments with more than 1 particle should be identifiable as different structures such as swollen mitochondria may produce a "one-particle within a membrane compartment" appearance. |
| **Recommendations for sampling and analysis** |  |  |
| General considerations |  | Prioritize analysis of few, carefully selected samples with a high viral load. Controls are not required for virus identification, because EM allows a direct (label-free) proof of the respective particles. |
| 1 | Sampling | Analyse multiple blocks of the most strongly RT-qPCR-positive samples (see 3), facultatively at different levels to locate infection foci, even in apparently suboptimal samples, such as samples with a relatively long post mortem interval, or samples that have been frozen or paraffin embedded. Identify virus particles in the best specimen to get an internal "positive control". |
| 2 | Correlation: IHC/ISH | Identify infection foci by using IHC or ISH and process corresponding FFPE regions via a punch biopsy or paraffin sections for EM as previously described<sup>8,21,36</sup>. Serial paraffin sections may be processed to stain virus antigens and cell type markers and detect intact virus particles of the same cell. |
| 3 | Correlation: RT-qPCR | Try to roughly estimate the likelihood of finding virus particles based on RT-qPCR data as shown in our work with 0.006 or 500 expected particles per mm<sup>2</sup> section area for low or high viral load, respectively. |
| 4 | Structural preservation/ image quality | Adjustment of fixation, embedding, sectioning and staining may be required for sufficient preservation of virus<sup>11,53</sup>. Use adequate magnification to clearly resolve all relevant details of possible virus particles (0.2-1 nm pixel size) and adjust the respective EM settings correctly<sup>53</sup>. |
| 5 | Labelling techniques | Pre-embedding or post-embedding techniques should be used and interpreted with caution, as structural preservation usually is negatively affected. Morphological features of virus particles should still be identifiable and adequate controls should be used. Additional conventional EM for virus detection should be used. |
| 6 | Screening | Screen individual sections systematically at a medium resolution at which groups of viruses are easily detected (as can be tested by our large-scale datasets), and in some regions also at higher resolution to avoid missing of single virus particles present in the cells. |
| 7 | Pattern recognition | Learn the visual pattern of virus particles in tissue samples by using correctly identified virus particles (see large-scale repository datasets on www.nanotomy.org). |
| 8 | Figures | Ensure adequate image size, resolution, color profile, brightness and contrast when publishing figures. |

### Analysis of publications demonstrating ultrastructural evidence for the presence of SARS-CoV-2 in human samples

We surveyed publications (published April 2020 to November 2021) using ultrastructural findings as proof of virus particles in human samples (Supplementary Tables 8,9) and re-evaluated the data using our refined criteria for virus identification. Six publications presented sufficient data to prove the presence of virus particles, while 116 publications misinterpreted different cellular structures as virus or showed only insufficient evidence for the presence of virus particles (Supplementary Figure 9). In total, only 63 of 292 electron micrographs (22%) showed sufficient structural preservation and image quality necessary for identifying structural details such as enveloped viruses. Structures misinterpreted as virus particles were mostly compatible with coated vesicles, vesicles of rough endoplasmic reticulum, multivesicular bodies, and autolytic mitochondria. Thirty-eight publications discussed the challenges of SARS-CoV-2 identification by EM (Supplementary Table 10).

## Discussion

One key question regarding severe to lethal COVID-19 is whether the clinical presentation including considerable organ damage is due to direct organ targeting of SARS-CoV-2 or downstream effects such as an overshooting immune response. During the COVID-19 pandemic, autopsy-driven research, using multimodal approaches, attempted at defining organ tropism of the virus and at unraveling organ-specific pathomechanisms ^8,9,25,41-43^, yet a critical and systematic study investigating the limitations of in situ detection of SARS-CoV-2 in autopsy tissues has not been performed.

Looking into the discrepancy of results from studies using autopsy material to determine organ tropism and especially identification of organ-specific target cell types of SARS-CoV-2 for most organ systems led to conflicting results hampering research progress ^3,8-10,44,45^. For instance, published data include studies revealing direct infection of neurons by SARS-CoV-2 with substantial neuroinvasion ^46^, single infected cells in a subset of patients ^25^, but also the absence of virus and COVID-19-specific alterations ^47^. Validations are complicated by the use of various, often not well-evaluated antibodies in different studies and the lack of sufficient positive and negative controls.

Here we determined the limitations of SARS-CoV-2 detection by IHC and thin section EM using a defined set of control tissues including FFPE highly susceptible human cell lines and autopsy tissues with expectable high SARS-CoV-2 viral load. Finally, we performed a multicenter study assessing how well IHC performs in detecting SARS-CoV-2 proteins in autopsy tissues.

Assessment of a wide range of commercially available antibodies directed at SARS-CoV-2 proteins showed that only a subset of these can be reliably used in autopsy tissues (Supplementary Figure 3). Interestingly, antibodies against nucleocapsid protein showed highest sensitivity. This may be since, of all SARS-CoV-2 proteins, nucleocapsid protein is produced at the highest levels during the lifecycle of SARS-CoV-2 in cells ^48^. Correspondingly, we found much less spike protein than nucleocapsid protein-based on both, the amount per cell and the general abundance in affected tissues. This should be considered when interpreting studies proving multi-organ tropism and claiming specific target cell types for SARS-CoV-2, especially by using anti-spike antibodies ^1,3,46,49,50^.

We found a discrepancy between viral RNA loads as determined by RT-qPCR and detection of SARS-CoV-2 proteins by immunohistochemistry. In fact, in tissues with low viral RNA loads immunohistochemistry is not a reliable method to determine organ tropism or target cell type as the interpretation of the rare immunosignals is difficult and nonspecific staining may be falsely interpreted as a positive signal. In agreement with this, we found low inter-observer reliability in tissues with low viral RNA loads (Supplementary Table 5). This may have to do with the fact that the distribution of the virus is uneven, even in highly affected organ systems such as lungs (Figure 3). The absence of viral proteins, on the other hand, cannot and should not be used as an argument for the absence of SARS-CoV-2-related tissue pathology, as autopsy tissues can only provide an incomplete snapshot of what has occurred in the sometimes very long clinical phase of the disease ^51^. Recent studies have tried to address this point by studying autopsy tissues at different stages of COVID-19 ^44^. Our study also disclosed that it is crucial to choose suitable control tissues for immunohistochemistry not only based on high viral RNA loads but also on tissue integrity. Lung, for example, the tissue with the highest viral RNA loads cannot be considered an optimal control tissue as it tends to produce false-positive signals and difficult to interpret staining patterns. This may be because extensive tissue damage seen in COVID-19 lungs leads to pre-necrotic epithelial cells and hyaline membranes, both prone to false-positive signals. Also, this study shows that lungs often contain SARS-CoV-2 protein-harboring cell debris and mucus leading to difficulties to interpret signals (Figure 3). However, these limitations may also be valid for other (autopsy) tissues. Thus, it is warranted to carefully discriminate between direct SARS-CoV-2 virus presence or infection and inflammation-related tissue pathology, since the latter might considerably contribute to false-positive signals in IHC.

Using EM to detect intact SARS-CoV-2 particles in autopsy tissues comes along with specific challenges such as a relatively low sensitivity as compared to e.g. RT-qPCR, limitations in cell-type identification, and high inter-observer variability depending on EM expertise. We found virus particles only in a minor fraction of patient samples (with comparatively high viral RNA load), also, virus particles were spatially highly confined. In fact, individual cells may contain up to tens of thousands CoV particles and hundreds of thousands of RNA copies. Thus, even in samples with a high SARS-CoV-2 load, few infected cells (∼20 or less per 10,000 cells), possibly also mobile cells, could make up a significant fraction of the total viral load. This result aligns with light microscopic findings suggesting a focal infection and argues for the complementarity of both methods so as not to misguide research ^18,21^. However, quantity of viral load may be cell type-specific and also depend on the disease phase. Moreover, presence of intracellular particles in e.g. macrophages may variably be a result of phagocytosis and not infection, thus indicating a need for further research on this correlation.

Based on our data, we provide recommendations on a suitable strategy for identifying virus-infected cells in Table 2. We slightly expanded and detailed previously published criteria ^20,22,23^ primarily based on examinations of virus particles produced by cell culture.

If these refined criteria for SARS-CoV-2 identification were applied to journal publications, 116 of 122 publications do not sufficiently prove the presence of viral particles in various human tissues. This problem has already been discussed ^20,23^ and resulted in specific recommendations for the correct detection of CoV particles. However, a general decline of diagnostic EM over the last decades with loss of expertise occurred ^18,52^, further aggravated by the unfamiliarity of most EM facilities with in-situ-detection of viruses ^23^. Both probably complicate transfer of these recommendations into practice, as also indicated by the lack of general quality standards ^53^ of many published EM data. This unfortunate and long-standing decline of diagnostic EM is further illustrated by the fact that also during the SARS-CoV pandemic in the early 2000s, detection of the virus by EM was tainted with technical and interpretational difficulties, and non-viral particles in different organs were used to propose a multi-organ tropism of SARS-CoV ^54-56^. These misinterpretations were then perpetuated early on in the SARS-CoV-2 pandemic ^57-64^. Importantly, misinterpretations of different cellular structures as virus also occurred in cell culture and organoids ^46,65^.

However, we emphasize that diagnostic EM is a valuable method for virus detection if appropriate standards are applied ^20^. It should always be pursued to validate other techniques of virus detection to gain reliable information on tropism and virus-induced alterations. Generally, virus detection by EM in a routine diagnostic setting is achievable if the recommendations and criteria provided in our work are considered. Moreover, it is necessary to learn visual patterns of virus within complex tissue samples to speed up the screening process. Our large-scale datasets, corresponding to approximately 130,000 conventional electron micrographs, may help in acquiring these visual pattern recognition skills, which cannot be acquired using small sets of preselected conventionally published electron micrographs lacking cellular and microanatomical context. As demonstrated by our Supplementary Video, virus particles can be found and identified in the complex structural matrix of lung tissue by spanning the scales from millimeter to nanometer. This technique ^27,66^ also provides a promising approach for fast and precise ultrastructural *in silico* analysis for future pandemics, especially in light of innovative high-throughput EM imaging approaches ^67^. In summary, usage of autopsy tissues with in situ detection of SARS-CoV-2 is valuable if interpreted within the limits of all applied methods and tissues. In the early phase of the COVID-19 pandemic, for various reasons, researchers have not fully abided to this, opening the door for misinterpretation and overestimation of SARS-CoV-2 multi-organ tropism. The here formulated consensus criteria can provide guidance to improve quality autopsy-based SARS-CoV-2 research. The main limitation of our study is its limited scope regarding assessed tissues and anti-SARS-CoV-2 antibodies.

## Data Availability

All data produced in the present study are available upon reasonable request to the authors

http://www.nanotomy.org/

## Acknowledgements

SK (head) and KH (technician) are running the Core Facility for Experimental Pathology of the UKE (“Mouse Patho”). We thank Annette Gries, Hanna Jania and Gudrun Holland for excellent technical assistance and the UMIF/UKE for using their microscopes. Our condolences to the families and all those who have lost their loved ones during the ongoing pandemic. We thank all relatives who made the difficult decision to give their permission for autopsy and research.

## References

1. Bradley BT, Maioli H, Johnston R, et al. Histopathology and ultrastructural findings of fatal COVID-19 infections in Washington State: a case series. Lancet 2020; 396(10247): 320–32.

2. Araujo-Silva CA, Marcos AAA, Marinho PM, et al. Presumed SARS-CoV-2 Viral Particles in the Human Retina of Patients With COVID-19. JAMA Ophthalmol 2021.

3. Kanczkowski W, Evert K, Stadtmüller M, et al. COVID-19 targets human adrenal glands. The Lancet Diabetes & Endocrinology 2021; 10(1): 13–6.

4. Han Y, Duan X, Yang L, et al. Identification of SARS-CoV-2 inhibitors using lung and colonic organoids. Nature 2021; 589(7841): 270–5.

5. Dolhnikoff M, Ferreira Ferranti J, de Almeida Monteiro RA, et al. SARS-CoV-2 in cardiac tissue of a child with COVID-19-related multisystem inflammatory syndrome. Lancet Child Adolesc Health 2020; 4(10): 790–4.

6. Ackermann M, Verleden SE, Kuehnel M, et al. Pulmonary Vascular Endothelialitis, Thrombosis, and Angiogenesis in Covid-19. N Engl J Med 2020; 383(2): 120–8.

7. von Stillfried S, Boor P. Detection methods for SARS-CoV-2 in tissue. Pathologe 2021.

8. Meinhardt J, Radke J, Dittmayer C, et al. Olfactory transmucosal SARS-CoV-2 invasion as a port of central nervous system entry in individuals with COVID-19. Nat Neurosci 2021; 24(2): 168–75.

9. Puelles VG, Lutgehetmann M, Lindenmeyer MT, et al. Multiorgan and Renal Tropism of SARS-CoV-2. N Engl J Med 2020; 383(6): 590–2.

10. Yang AC, Kern F, Losada PM, et al. Dysregulation of brain and choroid plexus cell types in severe COVID-19. Nature 2021; 595(7868): 565–71.

11. Goldsmith CS, Miller SE. Modern uses of electron microscopy for detection of viruses. Clin Microbiol Rev 2009; 22(4): 552–63.

12. Roingeard P, Raynal PI, Eymieux S, Blanchard E. Virus detection by transmission electron microscopy: Still useful for diagnosis and a plus for biosafety. Rev Med Virol 2019; 29(1): e2019.

13. Wrapp D, Wang N, Corbett KS, et al. Cryo-EM structure of the 2019-nCoV spike in the prefusion conformation. Science 2020; 367(6483): 1260–3.

14. Turonova B, Sikora M, Schurmann C, et al. In situ structural analysis of SARS-CoV-2 spike reveals flexibility mediated by three hinges. Science 2020; 370(6513): 203–8.

15. Ogando NS, Dalebout TJ, Zevenhoven-Dobbe JC, et al. SARS-coronavirus-2 replication in Vero E6 cells: replication kinetics, rapid adaptation and cytopathology. J Gen Virol 2020; 101(9): 925–40.

16. Laue M, Kauter A, Hoffmann T, Moller L, Michel J, Nitsche A. Morphometry of SARS-CoV and SARS-CoV-2 particles in ultrathin plastic sections of infected Vero cell cultures. Sci Rep 2021; 11(1): 3515.

17. Cortese M, Lee JY, Cerikan B, et al. Integrative Imaging Reveals SARS-CoV-2-Induced Reshaping of Subcellular Morphologies. Cell Host Microbe 2020; 28(6): 853–66 e5.

18. Dittmayer C, Meinhardt J, Radbruch H, et al. Why misinterpretation of electron micrographs in SARS-CoV-2-infected tissue goes viral. Lancet 2020; 396(10260): e64–e5.

19. Goldsmith CS, Miller SE, Martines RB, Bullock HA, Zaki SR. Electron microscopy of SARS-CoV-2: a challenging task. Lancet 2020; 395(10238): e99.

20. Bullock HA, Goldsmith CS, Zaki SR, Martines RB, Miller SE. Difficulties in Differentiating Coronaviruses from Subcellular Structures in Human Tissues by Electron Microscopy. Emerg Infect Dis 2021; 27(4): 1023–31.

21. Martines RB, Ritter JM, Matkovic E, et al. Pathology and Pathogenesis of SARS-CoV-2 Associated with Fatal Coronavirus Disease, United States. Emerg Infect Dis 2020; 26(9): 2005–15.

22. Bullock HA, Goldsmith CS, Miller SE. Best practices for correctly identifying coronavirus by transmission electron microscopy. Kidney Int 2021; 99(4): 824–7.

23. Hopfer H, Herzig MC, Gosert R, et al. Hunting coronavirus by transmission electron microscopy - a guide to SARS-CoV-2-associated ultrastructural pathology in COVID-19 tissues. Histopathology 2021; 78(3): 358–70.

24. Steenblock C, Richter S, Berger I, et al. Viral infiltration of pancreatic islets in patients with COVID-19. Nat Commun 2021; 12(1): 3534.

25. Matschke J, Lutgehetmann M, Hagel C, et al. Neuropathology of patients with COVID-19 in Germany: a post-mortem case series. Lancet Neurol 2020; 19(11): 919–29.

26. Burel JM, Besson S, Blackburn C, et al. Publishing and sharing multi-dimensional image data with OMERO. Mamm Genome 2015; 26(9-10): 441-7.

27. Dittmayer C, Goebel HH, Heppner FL, Stenzel W, Bachmann S. Preparation of Samples for Large-Scale Automated Electron Microscopy of Tissue and Cell Ultrastructure. Microsc Microanal 2021; 27(4): 815–27.

28. Cardona A, Saalfeld S, Schindelin J, et al. TrakEM2 software for neural circuit reconstruction. PLoS One 2012; 7(6): e38011.

29. Bankhead P, Loughrey MB, Fernandez JA, et al. QuPath: Open source software for digital pathology image analysis. Sci Rep 2017; 7(1): 16878.

30. Schindelin J, Arganda-Carreras I, Frise E, et al. Fiji: an open-source platform for biological-image analysis. Nat Methods 2012; 9(7): 676–82.

31. McHugh ML. Interrater reliability: the kappa statistic. Biochem Med (Zagreb) 2012; 22(3): 276–82.

32. Goldsmith CS, Tatti KM, Ksiazek TG, et al. Ultrastructural characterization of SARS coronavirus. Emerg Infect Dis 2004; 10(2): 320–6.

33. Eskelinen EL. To be or not to be? Examples of incorrect identification of autophagic compartments in conventional transmission electron microscopy of mammalian cells. Autophagy 2008; 4(2): 257–60.

34. Eustaquio T, Wang C, Dugard CK, et al. Electron microscopy techniques employed to explore mitochondrial defects in the developing rat brain following ketamine treatment. Exp Cell Res 2018; 373(1-2): 164-70.

35. Afzelius BA. Ultrastructure of human nasal epithelium during an episode of coronavirus infection. Virchows Arch 1994; 424(3): 295–300.

36. Shieh WJ, Hsiao CH, Paddock CD, et al. Immunohistochemical, in situ hybridization, and ultrastructural localization of SARS-associated coronavirus in lung of a fatal case of severe acute respiratory syndrome in Taiwan. Hum Pathol 2005; 36(3): 303–9.

37. Cheung OY, Chan JW, Ng CK, Koo CK. The spectrum of pathological changes in severe acute respiratory syndrome (SARS). Histopathology 2004; 45(2): 119–24.

38. Wong KF, To TS, Chan JK. Severe acute respiratory syndrome (SARS). Br J Haematol 2003; 122(2): 171.

39. Ksiazek TG, Erdman D, Goldsmith CS, et al. A novel coronavirus associated with severe acute respiratory syndrome. N Engl J Med 2003; 348(20): 1953–66.

40. Hocke AC, Becher A, Knepper J, et al. Emerging human middle East respiratory syndrome coronavirus causes widespread infection and alveolar damage in human lungs. Am J Respir Crit Care Med 2013; 188(7): 882–6.

41. Wenzel J, Lampe J, Muller-Fielitz H, et al. The SARS-CoV-2 main protease M(pro) causes microvascular brain pathology by cleaving NEMO in brain endothelial cells. Nat Neurosci 2021; 24(11): 1522–33.

42. Ferreira-Gomes M, Kruglov A, Durek P, et al. SARS-CoV-2 in severe COVID-19 induces a TGF-beta-dominated chronic immune response that does not target itself. Nat Commun 2021; 12(1): 1961.

43. Aschman T, Schneider J, Greuel S, et al. Association Between SARS-CoV-2 Infection and Immune-Mediated Myopathy in Patients Who Have Died. JAMA Neurol 2021; 78(8): 948–60.

44. Khan M, Yoo SJ, Clijsters M, et al. Visualizing in deceased COVID-19 patients how SARS-CoV-2 attacks the respiratory and olfactory mucosae but spares the olfactory bulb. Cell 2021; 184(24): 5932–49 e15.

45. von Stillfried S, Villwock S, Bulow RD, et al. SARS-CoV-2 RNA screening in routine pathology specimens. Microb Biotechnol 2021; 14(4): 1627–41.

46. Song E, Zhang C, Israelow B, et al. Neuroinvasion of SARS-CoV-2 in human and mouse brain. J Exp Med 2021; 218(3).

47. Deigendesch N, Sironi L, Kutza M, et al. Correlates of critical illness-related encephalopathy predominate postmortem COVID-19 neuropathology. Acta Neuropathol 2020; 140(4): 583–6.

48. V’Kovski P, Kratzel A, Steiner S, Stalder H, Thiel V. Coronavirus biology and replication: implications for SARS-CoV-2. Nat Rev Microbiol 2021; 19(3): 155–70.

49. Cantuti-Castelvetri L, Ojha R, Pedro LD, et al. Neuropilin-1 facilitates SARS-CoV-2 cell entry and infectivity. Science 2020; 370(6518): 856–60.

50. Schurink B, Roos E, Radonic T, et al. Viral presence and immunopathology in patients with lethal COVID-19: a prospective autopsy cohort study. Lancet Microbe 2020; 1(7): e290–e9.

51. Fitzek A, Schadler J, Dietz E, et al. Prospective postmortem evaluation of 735 consecutive SARS-CoV-2-associated death cases. Sci Rep 2021; 11(1): 19342.

52. de Haro T, Furness P. Current and future delivery of diagnostic electron microscopy in the UK: results of a national survey. J Clin Pathol 2012; 65(4): 357–61.

53. Stirling JW, Curry A. Quality standards for diagnostic electron microscopy. Ultrastruct Pathol 2007; 31(5): 365–7.

54. Gu J, Gong E, Zhang B, et al. Multiple organ infection and the pathogenesis of SARS. J Exp Med 2005; 202(3): 415–24.

55. Ding Y, He L, Zhang Q, et al. Organ distribution of severe acute respiratory syndrome (SARS) associated coronavirus (SARS-CoV) in SARS patients: implications for pathogenesis and virus transmission pathways. J Pathol 2004; 203(2): 622–30.

56. Xu J, Zhong S, Liu J, et al. Detection of severe acute respiratory syndrome coronavirus in the brain: potential role of the chemokine mig in pathogenesis. Clin Infect Dis 2005; 41(8): 1089–96.

57. Li YC, Bai WZ, Hashikawa T. The neuroinvasive potential of SARS-CoV2 may play a role in the respiratory failure of COVID-19 patients. J Med Virol 2020; 92(6): 552–5.

58. Baig AM. Neurological manifestations in COVID-19 caused by SARS-CoV-2. CNS Neurosci Ther 2020; 26(5): 499–501.

59. Kotfis K, Williams Roberson S, Wilson JE, Dabrowski W, Pun BT, Ely EW. COVID-19: ICU delirium management during SARS-CoV-2 pandemic. Crit Care 2020; 24(1): 176.

60. Wu Y, Xu X, Chen Z, et al. Nervous system involvement after infection with COVID-19 and other coronaviruses. Brain Behav Immun 2020; 87: 18–22.

61. Zhou Z, Kang H, Li S, Zhao X. Understanding the neurotropic characteristics of SARS-CoV-2: from neurological manifestations of COVID-19 to potential neurotropic mechanisms. J Neurol 2020; 267(8): 2179–84.

62. Li Z, Liu T, Yang N, et al. Neurological manifestations of patients with COVID-19: potential routes of SARS-CoV-2 neuroinvasion from the periphery to the brain. Front Med 2020; 14(5): 533–41.

63. Paniz-Mondolfi A, Bryce C, Grimes Z, et al. Central nervous system involvement by severe acute respiratory syndrome coronavirus-2 (SARS-CoV-2). J Med Virol 2020; 92(7): 699–702.

64. Jiang RD, Liu MQ, Chen Y, et al. Pathogenesis of SARS-CoV-2 in Transgenic Mice Expressing Human Angiotensin-Converting Enzyme 2. Cell 2020; 182(1): 50–8 e8.

65. Xiao K, Zhai J, Feng Y, et al. Isolation of SARS-CoV-2-related coronavirus from Malayan pangolins. Nature 2020; 583(7815): 286–9.

66. de Boer P, Pirozzi NM, Wolters AHG, et al. Large-scale electron microscopy database for human type 1 diabetes. Nat Commun 2020; 11(1): 2475.

67. Zheng Z, Lauritzen JS, Perlman E, et al. A Complete Electron Microscopy Volume of the Brain of Adult Drosophila melanogaster. Cell 2018; 174(3): 730–43 e22.

68. Zhu N, Zhang D, Wang W, et al. A Novel Coronavirus from Patients with Pneumonia in China, 2019. N Engl J Med 2020; 382(8): 727–33.

