## Supplementary Material for "Assessing and improving the validity of COVID-19 autopsy studies - a multicenter approach to establish essential standards for immunohistochemical and ultrastructural analyses"

### Supplementary Methods

#### Infection of cells with SARS-CoV-2

Vero cells (ATCC® CRL-1006) were cultivated and maintained under standard conditions in DMEM containing 3% FCS, 1% Penicillin/ Streptomycin, 1% L-Glutamine, (200 mM), 1% Sodium pyruvate, 1% non-essential amino acids (all Gibco/ThermoFisher Scientific). In infection experiments for detection of SARS-CoV-2 proteins by IHC, SARS-CoV-2 isolate HH-1 was used [1]. Cells were grown to >90% confluence and infected with SARS-CoV-2 Hamburg isolate or Berlin at different multiplicities of infection (MOIs). 24 h post infection, plates were soaked for 30 min. in 4% formaldehyde solution to allow for virus inactivation and cell fixation. Cells were then used for downstream applications.
For the evaluation of virus particles by EM, the SARS-CoV-2 isolate Italy-INMI1 was used [2].

#### Immunocytochemistry of cells to detect SARS-CoV-2 proteins

Formaldehyde-fixed cells were carefully washed, permeabilized (PBS, 0.2% Triton X-100), and blocked (blocking buffer, Active Motif). Primary antibodies against SARS-CoV-2 were incubated at 4 °C overnight (anti-spike #1, #3, anti-N #7, #9, #12, anti-Nsp3 #5, and anti-dsRNA #6 (dilutions see Supplementary Table 2). Cells were thoroughly washed and Alexa-conjugated secondary antibodies (-488, -555; Thermo Fisher Scientific) were added for 1.5 h at room temperature. Cells were washed again, stained with DAPI, and mounted in Fluoromount-G (SouthernBiotech). Images were acquired with a Leica SP5 confocal microscope with a 63x oil objective and Leica application suite software (LAS-AF-lite).

*Production of cell blocks to detect SARS-CoV-2 proteins in formalin-fixed paraffin-embedded cells*

Formaldehyde-fixed Vero cells were washed in PBS, scraped from the tissue culture surface and harvested by centrifugation. Cells were post-fixed in formaldehyde solution for at least 48 h to model the fixation time of autopsy tissues. Un-infected cells of both cell lines served as negative control, respectively. Cell pellets were then embedded in 3% agarose and processed for dehydration and paraffin embedding using a Leica ASP300S tissue processor.
Paraffin sections of cells were cut at 2 µm and processed as described for autopsy tissues. SARS-CoV-2-infected and the corresponding un-infected negative cell preparations were always cut next to each other on one slide to enable identical staining conditions. Anti-SARS-CoV-2 antibodies that were used in this study are listed in Supplementary Table 2.

*Immunohistochemistry to detect SARS-CoV-2 spike and nucleoprotein in autopsy tissues*

Autopsy tissues were fixed in 4% buffered formaldehyde solution for at least 48 h and processed for paraffin embedding. For the detection of SARS-CoV-2 proteins, sections were cut (2 µm) and mounted. After dewaxing and inactivation of endogenous peroxidases (3% hydrogen peroxide), antibody specific antigen retrieval was performed. Anti-SARS-CoV-2 antibodies that were evaluated in our study are summarized in Supplementary Table 2. Immunohistochemical staining were performed using a Ventana Benchmark XT autostainer (Ventana, Tuscon, Arizona, USA). Samples for a give analysis including negative controls were always performed in one staining run to avoid unwanted bias. All staining were developed using the Ultra View Universal 3,3´-Diaminobenzidine (DAB) Detection Kit (Ventana, Roche) which contains both secondary antibodies (anti-rabbit and anti-mouse), DAB stain and counter staining reagent. Slides were examined by experienced morphologists in a blinded fashion and representative images were taken with a Leica DMD108 digital microscope.

Immunofluorescence staining of paraffin embedded tissues were performed as followed: Tissue sections were cut at 2 µm and deparaffinised sections were boiled for 30 min. in 10 mM citrate buffer, pH 6.0, for antigen retrieval. Subsequently, sections were permeabilized (PBS, 0.2% Triton X-100), and blocked (blocking buffer, Active Motif). Primary antibodies against SARS-CoV-2 (anti-spike #1, anti-N #7 and #9) were incubated at 4 °C overnight. Slides were thoroughly washed and Alexa-conjugated secondary antibodies (-488, -555; Thermo Fisher Scientific) were added for 1.5 h at room temperature. Slides were washed again, auto-fluorescence was quenched by treatment with TrueBlack (Biotium) according to the manufacturer's protocol, and sections were mounted in Fluoromount-G/DAPI. Images were acquired with a Leica SP5 confocal microscope using a 40x oil objective.

To investigate the cross-reactivity of the here tested SARS-CoV-2 antibodies against other common human virus species, we used the commercially available cell control array (#MB-CC VIR; Zytomed) with formalin-fixed in one single paraffin block embedded cells that were infected by Cytomegalo (CMV), Herpes Simplex (HSV Type 1 and 2), Epstein Barr (EBV), and Simian Virus 40 (SV40), respectively. Sections of this cell block that also comprised a control human muscle tissue punch were stained with our panel of SARS-CoV-2 antibodies as described above. All thirteen antibodies were tested for nonspecific binding, but produced only minimal cross-reactivity.

**RNA isolation and SARS-CoV-2 RNA detection from FFPE tissues**

Total RNA was isolated from formalin-fixed, paraffin-embedded human lung tissue sections (4 x 5 µm thick sections per case) using the RNeasy@ FFPE Kit (Qiagen) according to the manufacturer's instructions. The RNA was eluted in RNase-free water and mixed with 1 U $\mu l^{-1}$ RiboLock RNase inhibitor (Thermo Fisher Scientific).

SARS-CoV-2 RNA levels were determined by quantitative reverse transcription real-time PCR (RT-q-PCR) using the RealStar® SARS-CoV-2 RT-PCR Kit RUO (Altona Diagnostics). Briefly, 2 µl RNA was added as a template to the reaction mixture. A sample preparation control provided by the kit was used as an internal control. Furthermore, an extended dry spin step was performed for 10 min. at 17,000g at room temperature.

**Sample preparation for electron microscopy**

Samples of autopsy lung, olfactory mucosa, trachea, medulla oblongata, myocardium and kidney were processed according to locally established standard protocols (see also Supplementary Table 6).

*Sample processing at the Robert Koch Institute*

The autopsy lung tissue of EM case 1 was first frozen, parts of it were then broken under liquid nitrogen and fixed in the frozen state in a 2.5% glutaraldehyde/ 2% formaldehyde solution in 0.1 M sodium cacodylate buffer for several days, similarly as previously described [3].

Foci of infection in FFPE-embedded olfactory mucosa were selected for re-embedding via positive *in-situ* hybridization signal as recently described (the same resin block was used; [4]), also, regular-sized electron micrographs of the olfactory mucosa [4] and autopsy lung tissue [5,6] were shown. Further tissue processing, including the processing of Vero E6 cells, was performed according to a standard protocol, including *en bloc* staining with 0.1% tannic acid in 0.05 M Hepes buffer for 30 min. and 2% uranyl acetate in distilled water for 120 min. [7].

Negative staining of viruses from cell culture supernatant was done according a standard protocol [7] using phosphotungstic acid as a stain.

*Sample processing at the Department of Neuropathology, Charité – Universitätsmedizin Berlin*

Samples were either directly fixed in 2.5% glutaraldehyde in 0.1 M Na-cacodylate buffer (EM case 3, 5 and 6) or first fixed in 4% buffered formaldehyde solution with secondary fixation in 2.5% glutaraldehyde/ 2% formaldehyde solution in 0.1 M Na-cacodylate buffer (EM case 4). Further processing, also of fixed autopsy lung tissue of EM case 1, was performed as recently described for diagnostic muscle and nerve samples [8], including *en bloc* staining with 1% uranyl acetate and 0.1% phosphotungstic acid during the dehydration step of 70% acetone.

*Sample processing at the Department of Pathology, RWTH Aachen*

Samples were first fixed for 7-10 d in 4% buffered formaldehyde solution and secondary fixed in 3% glutaraldehyde in 0.1 M Soerensen`s phosphate buffer for 24 h. After post-fixation with 1% $OsO_4$ samples were dehydrated with ethanol and propylenoxide and embedded in epon resin as previously described [9]. Ultrathin sections were stained with 1% uranyl acetate and 1% lead citrate.

**Large-scale electron microscopy and transmission electron microscopy**

Semithin sections (500 nm) were prepared with an ultramicrotome (Ultracut E, Reichert-Jung) and stained with Richardson's stain to select a region for large-scale digitization, measuring approximately 1 $mm^2$.

For digitization of autopsy lung tissue (case 1) and infected Vero E6 cells, we used a Gemini 300 scanning electron microscope (SEM, Zeiss) with a scanning transmission electron microscopy (STEM) detector at 29 kV accelerating voltage, SmartSEM software (Zeiss) for general control of the SEM and Atlas 5 software (Fibics) for automated acquisition of large-scale datasets as recently described [8]. Parameters for screening of coronavirus (CoV) particles were 3-4 nm pixel size and 1.5-1.8 beam dwell time. We also included three previously published large-scale datasets [4].

Additionally, all 15 infected cells that were found in dataset 1 and 2 of EM case 1 were digitized at very high resolution with 1 nm pixel size and 1.5 µs dwell time including line averaging of 3 for quantitative analysis of CoV particles. Image acquisition time for screening lung datasets was between 2-4 d per section and for screening cell culture datasets about 23 and 13 h per section. Individual cells were recorded in 15-60 min. Individual regions were recorded manually via SmartSEM software at very high resolution.

For digitization of two regions in sections of the olfactory mucosa, a SEM (Teneo VS, Thermo Fisher) was used with the STEM3+ detector and MAPS software (30 kV, 25 pA, 5 µs dwell time and 1 or 1.22 nm pixel size).

For transmission EM, thin sections (60-80 nm) were inspected with different transmission electron microscopes. Images shown here were recorded either with a CCD (Megaview III, EMSIS) or a CMOS camera (Phurona, EMSIS). The negative staining image shown in the video was acquired with a transmission electron microscope (Tecnai Spirit, Thermo Fisher) operated at 120 kV.

For repository implementation on www.nanotomy.org and open access pan-and-zoom analysis, datasets of autopsy lung and infected cell culture were stitched and then exported into a tiled browser-based file format using Atlas 5 software. The olfactory mucosa datasets were stitched with Imaris (Bitplane), exported with Zoomify free and visualized with Openseadragon. Additionally, bigtif files of screening datasets and tif (raw data) files of infected cells, extracellular particles and non-infected cells were implemented for download of files and further raw data analysis.

**Correlation of RT-qPCR and ultrastructural data**

For a rough approximation of how many coronavirus particles may be present within the volume of a 60 nm ultrathin section per mm$^2$, we used the information of the $Log_{10}$ SARS-CoV-2 RNA copies/ DNA amount of 10,000 diploid nuclei. We assumed a simplified “standard” size of nuclei of 3.5 x 3.5 x 3.5 µm based on measurements of six representative nuclear profiles in one section of an autopsy lung (case 1; dataset 1 on www.nanotomy.org). A standard nucleus of thus 3500 nm height, therefore, corresponds to 58 sections of 60 nm thickness each. The section of dataset 1 contained 657 nuclear profiles per mm$^2$ which corresponds to the volume of 11.3 nuclei (675 profiles per section/ 58 profiles per nucleus) within the section volume. 5,011,872 virus RNA copies ($Log_{10}$ 5,011,872 = 6.7; EM case 1 in Supplementary Table 6) and 52 RNA copies ($Log_{10}$ 52 = 1.72; EM case 5) correspond to a tissue volume that has the DNA amount of 10,000 diploid nuclei, resulting in 5663 or 0.06 virus RNA copies for 11.3 nuclei, respectively. It is difficult to estimate the percentage of RNA copies that are present in form of intact virus particles. In EM case 1 we found approximately 2000 CoV particles in four digitized ultrathin sections measuring each approximately 1 mm$^2$ (1701 particles were counted within and next to infected cells and 200-300 further particles were located diffusely or in small clusters without being located next to infected cells), resulting in approximately 500 CoV particles per mm$^2$ section area. If we compare this number with the expected number of virus RNA copies as calculated above, we can deduce a ratio between virus particle number and virus RNA of roughly 1:10 (500 particles vs. ~5000 virus RNA copies).

If we apply this ratio for EM case 5 with only 52 RNA copies per 10,000 diploid nuclei, only 0.006 virus particles per $mm^2$ section area can be expected (assuming a similar number of standard nuclei present in the section).

As an alternative approach to estimate the possible number of CoV particles per section, we used the relation between known particle number (i.e. 500 CoV particles per $mm^2$ section area) and RNA copy number of the sample (i.e. 5,011,872 / 10,000 nuclei) to calculate the particle number for the known RNA copy of case 5 (i.e. 52 / 10,000 nuclei) which resulted in 0.005 virus particles per $mm^2$ of an ultrathin section.

For an estimation of how many CoV particles may be present in a single cell, we used cell 7 as an example that exhibits a nearly round shape and showed 620 intracellular particles. With a cell diameter of 12.2 µm and a section thickness of 0.060 µm (60 nm), we calculated a section volume of 7.014 $\mu m^3$ of this cell profile. Assuming a spherical shape of the cell, we calculated 951 $\mu m^3$ cell volume. By calculating ((950.776/ 7.014) * 620), we came up with 84,043 CoV particles. As one CoV particle is likely present as section profile on two consecutive sections, we came up with approximately 40,000 CoV particles, corresponding to approximately 400,000 RNA copies.

Of note, our calculations remain a thought experiment correlating RT-qPCR and EM data to estimate the likelihood of finding CoV particles in autopsy samples by EM. While sufficient experimental data for correlation of RT-qPCR and the presence of virus particles in EM is still missing, these considerations helped us to deal with the challenges of in situ detection of CoV particles.

**Human tissues and Ethics approval**

Autopsies were performed at the Institute of Legal Medicine of the University Medical Center Hamburg-Eppendorf, Germany and Institute of Pathology Charité Universitätsmedizin Berlin, and Institute of Pathology and Electron Microscopy Facility, RWTH University of Aachen, Germany. Use of human tissue for post mortem studies after conclusion of diagnostic procedures has been reviewed and approved by the institutional review board of the independent Ethics Committee of the Hamburg Chamber of Physicians (WF-051/20; protocol-no. PV7311) and by the local Ethics Committee (Berlin: EA2/066/20) and by the Charité-BIH COVID-19 research board, and by local Ethics Committee (Aachen: EK 304/20, EK 119/20, and EK 092/20), and the study is in line with the Declaration of Helsinki.

**Search and analysis of publications demonstrating immunohistochemical detection of SARS-CoV-2**

In this work, we reviewed all publications available in English on immunohistochemical detection of SARS-CoV-2 in tissues (as of 06 Nov. 2021) from the literature database PubMed and curated database LitCovid of PubMed (20 publications in total) and evaluated the detection methods and the interpretation of the result as positive or negative detection of SARS-CoV-2. One case report was excluded because a similar team of authors contributed to an extended study using the same methods. Two other papers were excluded, one article in Russian and one article with a focus on ACE2 distribution. The publication by Lean et al. [10] selected by LitCovid was replaced by a more suitable study from the same group.

**Search and analysis of publications demonstrating ultrastructural findings of SARS-CoV-2**

We used PubMed and continuously searched for “SARS-CoV-2” AND “autopsy” AND “electron microscopy” and also used Google, Google news and scholar with keywords “SARS-CoV-2”, “electron microscopy”, “patient”, “case”, “particles”, “detected”, “biopsy” and “autopsy” in different combinations and facultatively excluded “cryo electron”, “cryo EM”, “cryogenic” and “culture” for the search.
We collected publications that do not sufficiently prove the presence of SARS-CoV-2 in human autopsy and biopsy samples based on our refined criteria for ultrastructural virus identification in Supplementary Table 8. Publications with ultrastructural proof of SARS-CoV-2 are listed in Supplementary Table 9 and publications such as letters, author replies and reviews that discuss the challenges of ultrastructural identification of coronavirus are listed in Supplementary Table 10. All electron micrographs of the publications listed in Supplementary Tables 8 and 9, including insets of other regions which showed areas with putative coronavirus particles, were counted and analyzed regarding their structural preservation and image quality. Overview images were not counted as separate electron micrographs in this regard. Image series of stepwise increasing magnification were counted as one EM image. Sufficient structural preservation refers to the presence of biomembranes, cytoskeletal components and organelles. Sufficient image quality refers to sufficient combined optical and image resolution, adequate image focus, adequate image contrast and the absence of image distortions and also absence of preparation artefacts such as folds or contamination. Finally, we evaluated if the criteria we have defined in Table 2 were met or not and tried to identify the presented structures. We grouped samples from different regions of an organ that were examined in these publications under common category names (Supplementary Figure 9: e.g. gyrus rectus was changed to the brain; more tissue categories were grouped for the Graphical Abstract).
We did not include preprint publications or publications demonstrating electron micrographs of SARS-CoV-2-infected cell culture, slice culture or organoids and (topographic) SEM images or images of negative staining preparations. We also did not include publications that reported not having found coronavirus particles. However, we discuss aspects of these publications in our manuscript.

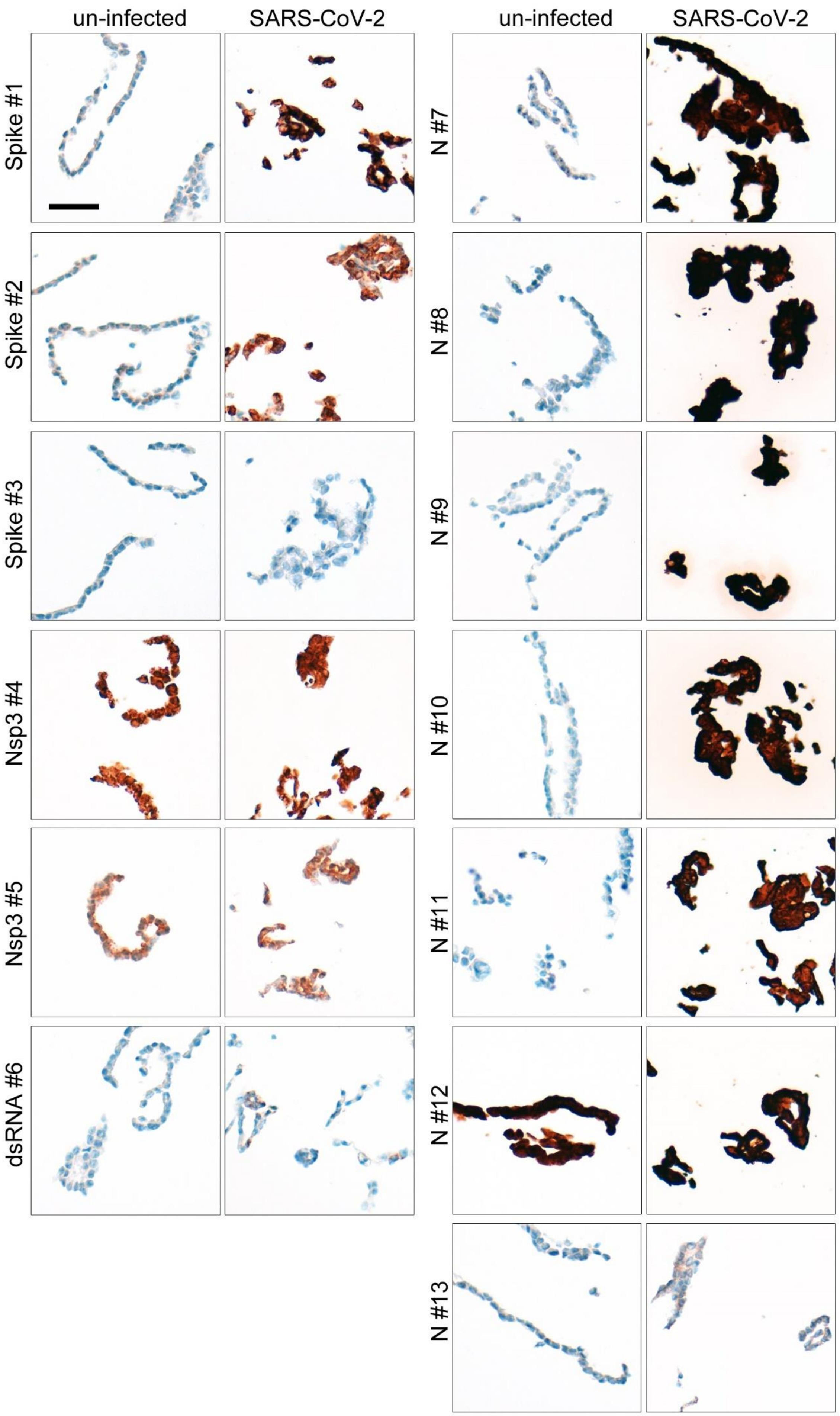

un-infected
SARS-CoV-2
un-infected
SARS-CoV-2
Spike #1
Spike #2
Spike #3
Nsp3 #4
Nsp3 #5
dsRNA #6
N #7
N #8
N #9
N #10
N #11
N #12
N #13

**Supplementary Figure 1: A subset of commercially available antibodies detect SARS-CoV-2 proteins in FFPE-cell blocks.** Evaluation of antibodies against SARS-CoV-2 proteins was performed in Vero cell blocks. Cells were infected with SARS-CoV-2, fixed in formaldehyde solution for >48 h, and embedded in paraffin in a similar way as human autopsy tissues. Sections of SARS-CoV-2- and un-infected cells were stained with commercially available antibodies against SARS-CoV-2 spike protein (Spike), nucleoprotein (N), non-structural protein 3 (Nsp3), or double-stranded RNA (dsRNA) (overview see Supplementary Table 2). Of note, some antibodies do not detect SARS-CoV-2 protein in infected cells in FFPE cell blocks or produce some or even high background staining in un-infected cells. These antibodies are not suited to detect SARS-CoV-2 in human autopsy tissue. Representative images are shown; Scale bar: 50 µm.

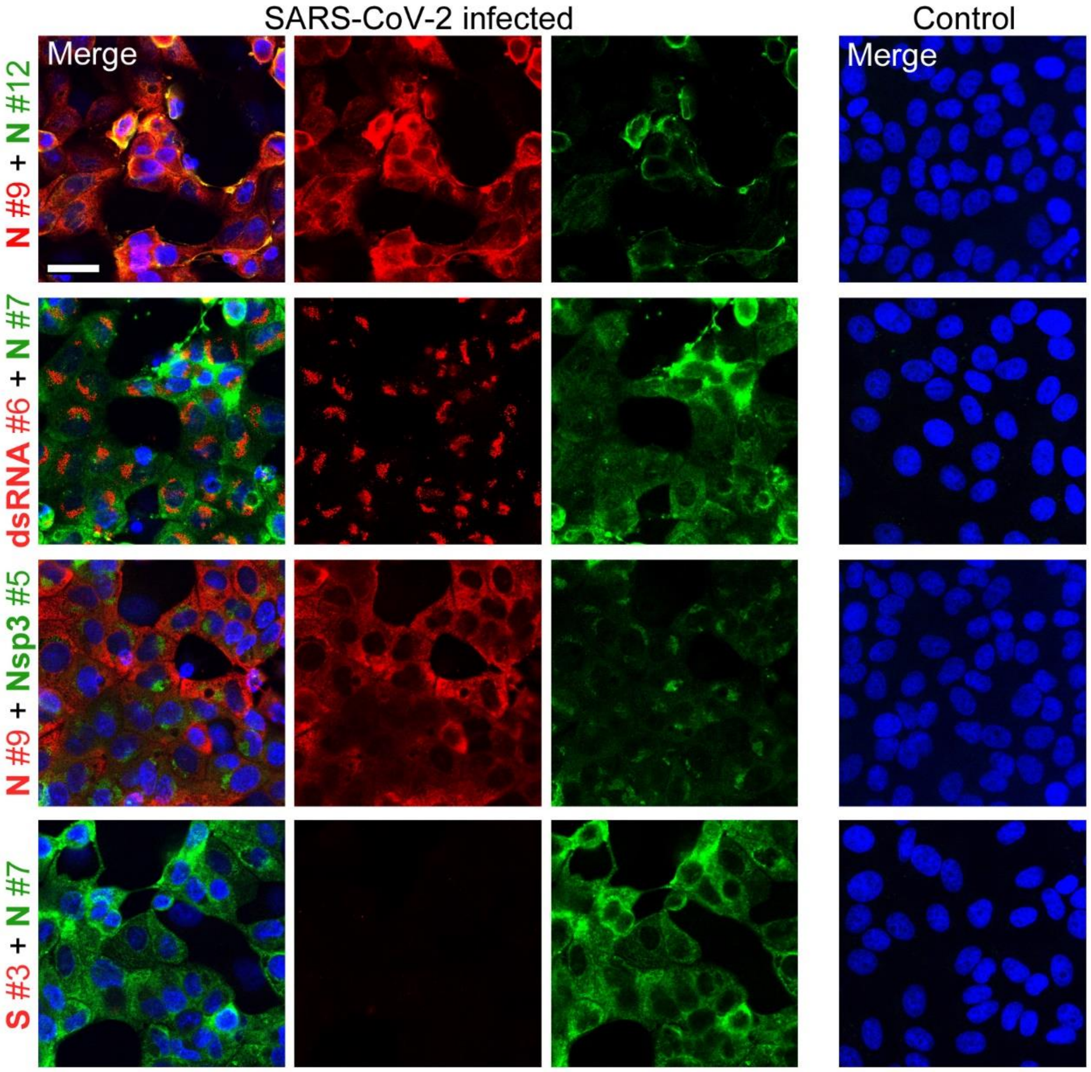

**Supplementary Figure 2: Selected antibodies detect SARS-CoV-2 proteins in infected cells, despite failure in FFPE tissues.** Antibodies that did not detect SARS-CoV-2 proteins in FFPE infected cells (antibodies #3 spike, #5 Nsp3, #6 dsRNA) or produced unfavourable signal-to-noise ratio (antibody #12 nucleocapsid) were further tested in infected Vero cells (MOI 10 24 h post infection) that were fixed in formalin, but not embedded in paraffin, thus, avoiding antigen retrieval. Cells were double-stained with the well performing antibodies #7 or #9 against nucleocapsid, respectively, to verify the infection level of the cells. Un-infected cells served as control. Nuclei were counterstained with DAPI (blue), scale bar: 25 µm. Antibody #12 show a low, but specific signal in infected cells. The non-structural marker for SARS-CoV-2 infection Nsp3 (antibody #5) and dsRNA (antibody #6) could be specifically detected in infected cells without paraffin embedding. Of note, one widely used and published antibody (#3 against spike) does not show staining of infected cells.

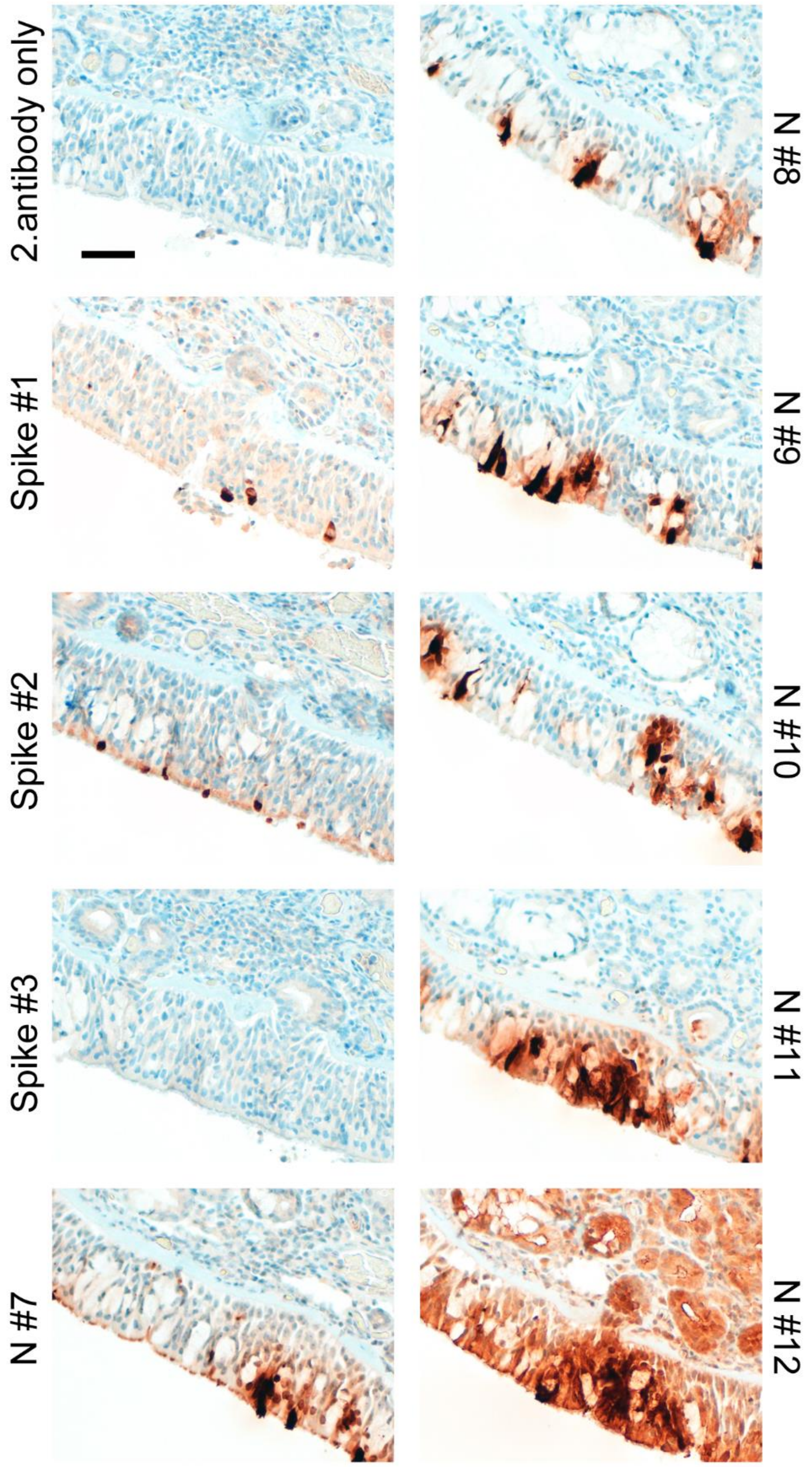

**Supplementary Figure 3: SARS-CoV-2 positive human respiratory mucosa was stained with different anti-SARS-antibodies.** Consecutive sections of formalin-fixed paraffin-embedded human respiratory mucosa was stained with selected antibodies to enable comparability (see Supplementary Table 2 for full panel of antibodies). Antibodies recognize SARS-CoV-2 spike and nucleocapsid protein (N) in respiratory mucosa of COVID-19 patients. Of note, as in lung tissue, the signal for SARS-CoV-2 spike protein is much lower than that of nucleocapsid. Staining results are much clearer in the respiratory mucosa than in lung tissue, where pathology and abundance of cellular debris is more sever. One widely used antibody (Spike #3) failed to detect spike protein in SARS-CoV-2 positive respiratory mucosa. Moreover, one other widely used antibody (N#12) produced very high background staining in surrounding tissue. Staining with the respective secondary antibody only as control show neglectable background in the respiratory mucosa and adjacent tissue. Representative images are shown; scale bar: 50 µm

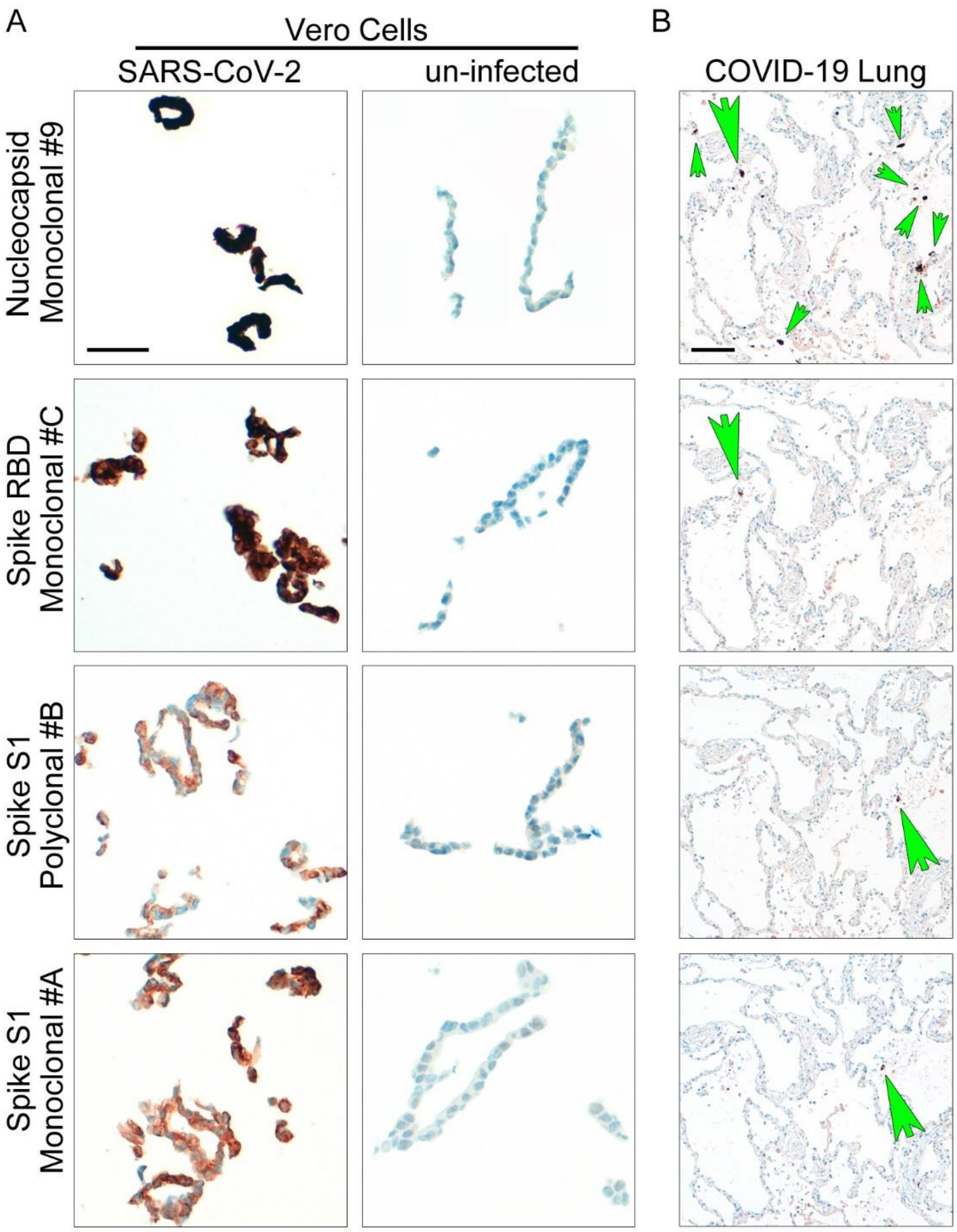

**Supplementary Figure 4: The abundance of SARS-CoV-2 spike protein in tissue is low independent of the detecting antibody.** Detection of SARS-CoV-2 spike protein in lungs of COVID-19 patients is low compared to abundance of nucleocapsid. To rule out antibody specific effects, we investigated this phenomenon with independent anti-spike antibodies that are directed against different parts of the spike protein. An antibody against nucleocapsid (#9) served as positive control. (A) Representative staining of infected and un-infected FFPE Vero cells show the specificity of the anti-spike antibodies which are directed against S1 and receptor-binding domain (RBD). Scale bar: 50 µm. (B) Representative images of staining with anti-nucleocapsid and three different anti-spike antibodies on consecutive sections of COVID-19 lung tissue confirmed our finding that abundance of spike protein is low in comparison to nucleocapsid. Small green arrows point to nucleocapsid-positive cells. Big green arrows point to single spike protein-positive cells in consecutive sections. Scale bar: 100 µm.

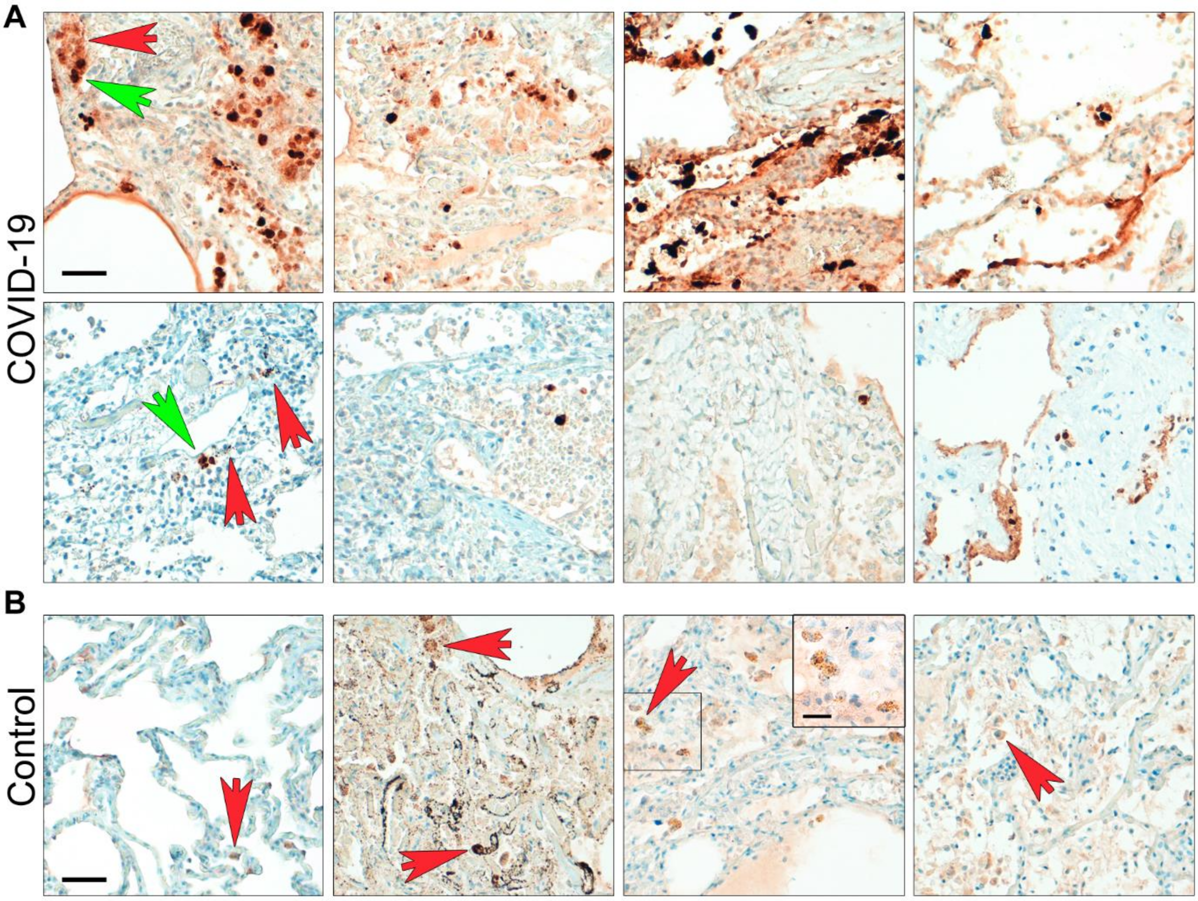

**Supplementary Figure 5: Abundance of SARS-CoV-2 nucleocapsid in lung tissue in COVID-19 patients is highly variable.** We determined monoclonal antibodies against nucleocapsid as suitable tools with a favourable signal to background ratio to determine virus abundance in human autopsy tissue of COVID-19 deceased. (A) Representative staining of SARS-CoV-2 nucleocapsid with a monoclonal antibody (N#9) is shown. The amount of virus protein+-cells in lung tissue varies highly. Moreover, presentation of staining is different which is complicating the evaluation of staining results. Green arrows point to positive staining of nucleocapsid, while red arrows point to bleeding of DAB staining to adjacent tissue in very positive tissues (upper row) and carbon deposition in lung tissue (lower row). Scale bar: 50 µm. (B) Representative images of non-COVID-19 control tissue display the problems with nonspecific background staining even with reliable antibodies. Red arrows point to nonspecific background staining and carbon depositions. Note that in contrast to the plane nucleocapsid staining, nonspecific background present in a punctate pattern, usually with a difference in colour: yellowish staining of background versus robust black-brown staining of real antibody signal. Scale bar: 50 µm; close-up: 20 µm.

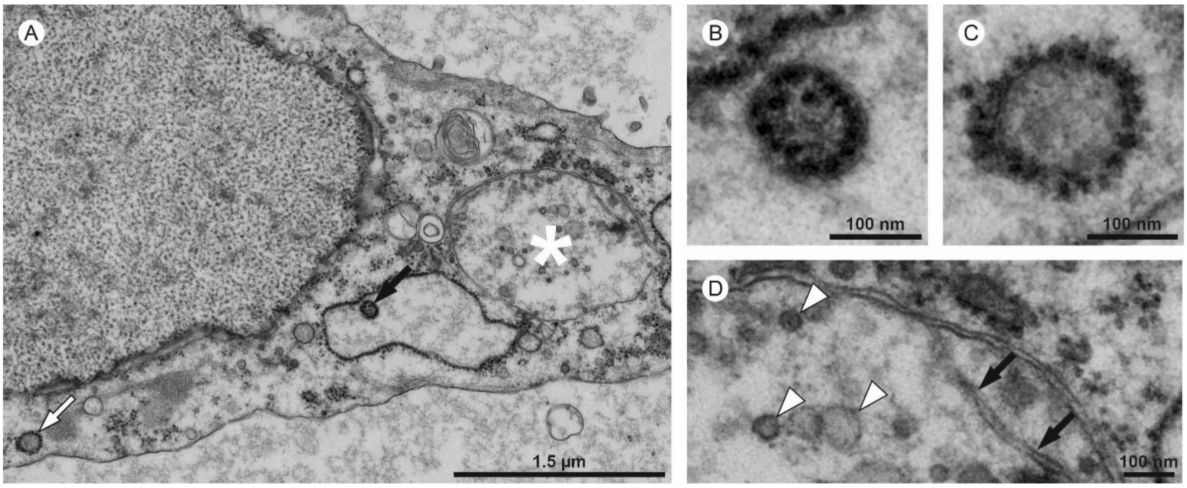

**Supplementary Figure 6: Common ultrastructural mimics of coronavirus particles**. (A) This cell within the alveolar lumen demonstrates multiple structures that are commonly misinterpreted as coronavirus (CoV) particles. Invaginations or vesicles within the rough endoplasmic reticulum (rER, A; black arrow and B) can be identified by larger, more rounded and often peripherally located electron dense dots (=ribosomes) as compared to the smaller (7 vs 20 nm) and more pleomorphic (sometimes net and sausage-shaped) electron dense ribonucleoprotein (RNP) profiles of CoV particles. rER vesicles within the cytoplasm additionally show untypical location (not enclosed in membrane compartments) as well as peripherally located electron dense dots (white arrow in A, and C). Autolytic (swollen and vesiculated) mitochondria (white asterisk in A, and D) can be identified by their limiting double membrane and the presence of cristae-like internal membranes (black arrows), less electron density (absence of RNP) and higher pleomorphism of artificially formed vesicles (white arrowheads) as compared to CoV particles. Coated vesicles (not illustrated here) contain no RNP or similar structures, are located in the cytoplasm (not enclosed in other intracellular membrane compartments) and show densely packed and shorter surface projections as compared to CoV particles. All these mimics could be separated from CoV particles, especially due to the high number of CoV particles in most infected cells as compared to mimics that are found in virtually every cell, but mostly at low numbers.

See [www.nanotomy.org](http://www.nanotomy.org) for high-resolution dataset of the demonstrated cell.

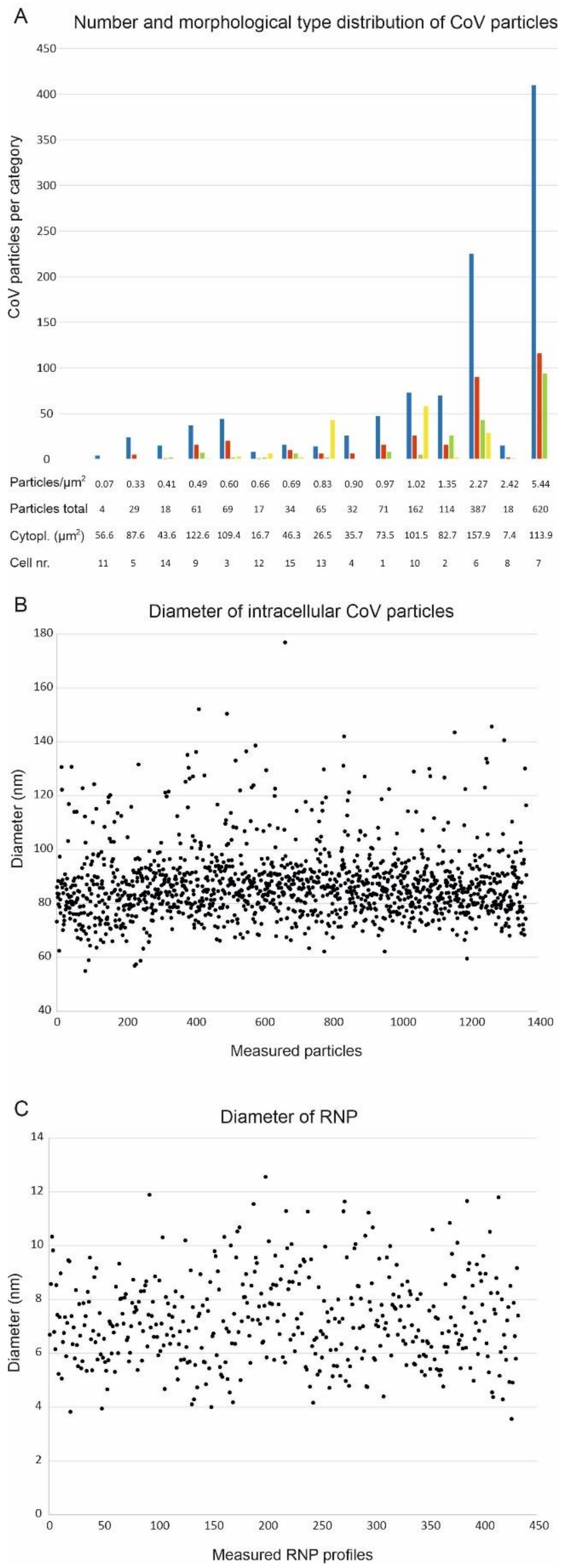

**Supplementary Figure 7: Quantitative analysis of coronavirus particles.** (A) Scoring of different coronavirus (CoV) particle types within the individual infected cells highlight that CoV type 1 particles (blue) are the most common morphological type. CoV type 2 (red), CoV type 3 (green) and CoV type 4 (yellow) (see Figure 5 for further details of CoV types). Density of CoV particles is demonstrated as intracellular particles/ µm$^2$ cytoplasmic (Cytopl.) area in the respective infected cells. Total particles also include extracellular particles. The differences in total CoV particle number clarify that all cells within a section may need to be carefully examined to detect infected cells that are concentrated very focally (11 vs. 4 vs. 0 vs. 0 infected cells per section) and many cells only show a few CoV particles. Alternatively, faster screening of multiple resin blocks and levels may be performed at lower magnification to detect only cells with many intracellular CoV particles, thereby identifying only hot spots of the infection, which are most likely of highest relevance. (B and C) Distribution of CoV particle diameter and ribonucleoprotein (RNP) diameter. Note that the RNP clearly shows a different diameter as compared to ribosomes which measure approximately 20 nm [11].

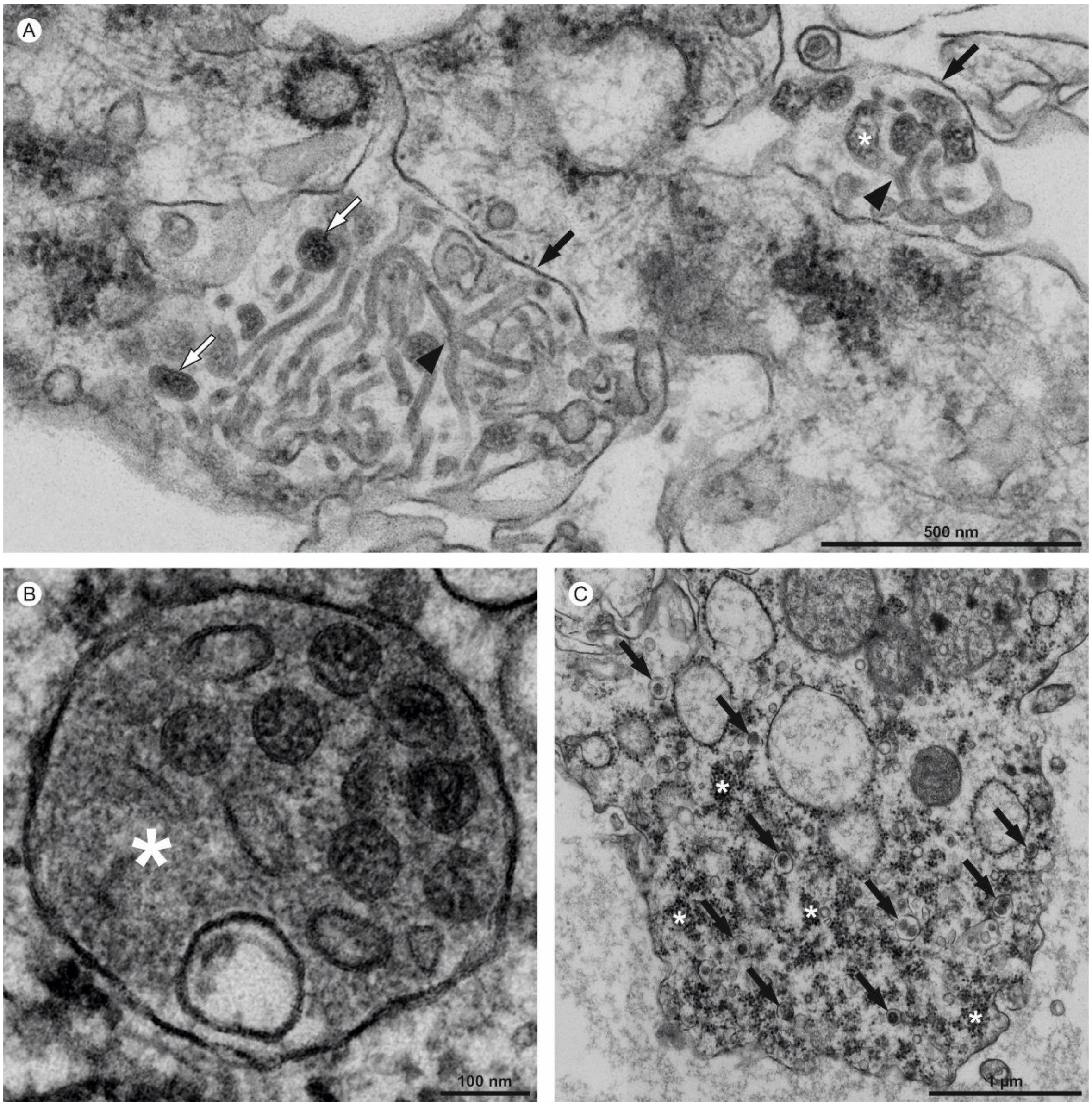

**Supplementary Figure 8: Peculiar ultrastructural findings in SARS-CoV-2-infected autopsy lung tissue.** (A; cell 15) Tubular structures, (B; cell 7) vesicle with granular material (white asterisk) and CoV particles and infected cell (C; cell 9) with a cytoplasmic area demonstrating numerous small vesicles with one or two profiles of CoV particles (black arrows) and prominent granular, ribosome-like material (white asterisks) (A-C; compare Goldsmith et al. 2004 [12]). (A) Cells 2 and 15 demonstrated prominent tubular structures, also, cells 8 and 12 showed few such profiles. The tubules appear to be located within membrane compartments (black arrows). Some branching figures are noted (black arrowheads). Interestingly, multiple CoV particles (white arrows), some with bizarre shape and some with stalk-like projections towards the tubules (white asterisk) were found, albeit with blurred transition. Cells 2 and 15 showed prominent tubular structures with a mean diameter of 29 nm (+- 4.2 nm; 25 profiles measured; 22 to 36 nm), while cells 8 and 12 only showed single profiles (not measured).

See [www.nanotomy.org](www.nanotomy.org) for high-resolution datasets of the demonstrated cells.

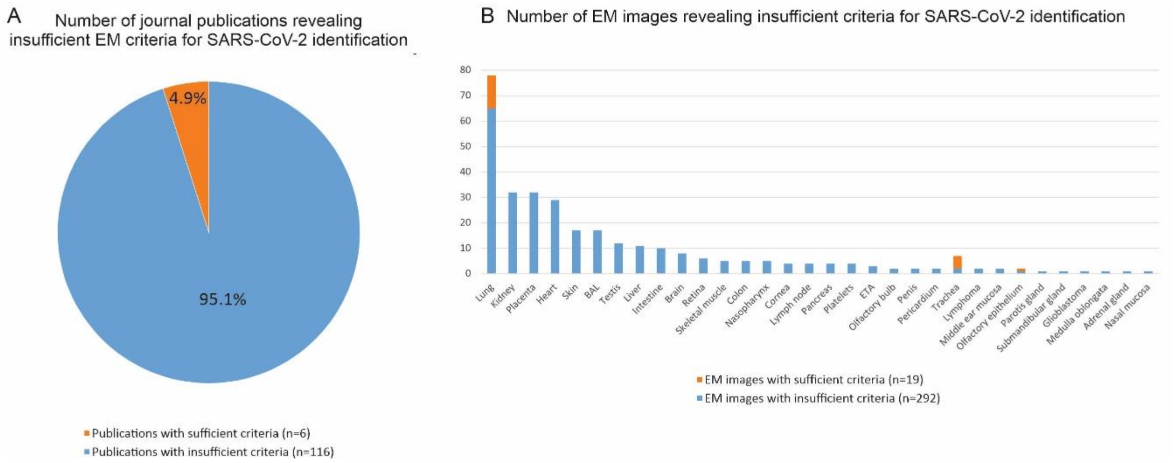

**Supplementary Figure 9: Results of the analysis of publications on the presence of putative SARS-CoV-2 particles in human autopsy and biopsy tissues.** (A) 4.9% of these publications (6 of 122) unequivocally prove the presence of coronavirus (CoV) particles. The remaining 95.1% of publications (116 of 122), according to our refined criteria for identification of CoV particles, do not prove the presence of CoV particles. A detailed discussion of all publications including the individual electron micrographs is provided in Supplementary Tables 8 and 9. (B) Among the 116 publications, in total, 292 electron micrographs of different tissues are provided; lung, kidney, placenta and heart are the most frequent. Also, electron micrographs of immunogold labelling are shown, but no morphological aspects of CoV particles are visible and no controls are shown. One publication does not provide electron micrographs [13]. Due to the limited quality of the samples and images, no exact values are given for the individual possible identity of the structures. Among the six publications that demonstrate CoV particles (two articles, one letter to the editor, one reply letter and two reviews), 19 EM images show particles in the lung, upper airway/trachea and within the olfactory mucosa. See Supplementary Table 10 for letters, replies and reviews discussing the challenges of ultrastructural SARS-CoV-2 identification.

**Supplementary Table 1.** Pubmed Search for „SARS-CoV-2“ AND „detection“ AND „IHC“

| Nr. | Original reference | Nr Antibodies used in study | Antibodies evaluated in study | FFPE cells | FFPE tissue shown | Comments/Images |
|---|---|---|---|---|---|---|
| 1 | Best Rocha et al. [14] | 3 | no | no | Lung, Kidney | Not specified, which antibody was used for images |
| 2 | Borczuk et al. [15] | 1 | * | no | Lung | Comparison of IHC and ISH |
| 3 | Carossino et al. [16] | 4 | yes | Infected Vero | no | Comparison of IHC and ISH; all antibodies shown |
| 4 | Duarte-Neto et al. [17] | 2 | no | no | Lung, Liver, Kidney, Brain, Skin, Gut, Bone marrow | Non-validated antibodies that have not been used in the other studies; no controls; should be confirmed by independent staining |
| 5 | Duarte-Neto et al. [18] | 1 | no | no | Testes | Non-validated antibodies that have not been used in the other studies; no controls; should be confirmed by independent staining |
| 6 | El Jamal et al. [19] | 2 | yes | Infected Vero | Lung | Non-validated antibodies that have not been used in the other studies; no negative cell controls shown for evaluation |
| 7 | Hong et al.[20] | 1 | partially | no | Hamster Lung | Novel antibody that has not been used in the other studies; evaluation by Western Blot; no negative controls |
| 8 | Ko et al. [21] | 1 | no | no | Skin biopsy | No negative control included in study |
| 9 | Lacavalla et al. [22] | 0 | no | no | no | Reference to other article with a non-validated antibody; should be confirmed by independent staining |
| 10 | Lean et al. [10] | 3 | yes | Infected Vero; transfected HEK 293T-17 | Infected ferret: nasal turbinates | Comparison of IHC and ISH; all antibodies shown; no negative control for ferret tissue |
| 11 | Liu et al. [23] | 7 | yes | Infected Vero | no | Comparison of IHC, fluorescence staining, and ISH; not all antibodies shown |
| 12 | Massoth et al. [24] | 1 | no | no | Lung | Comparison of IHC, RT-qPCR and ISH; non-validated antibody; article not freely available |
| 13 | McMullen et al. [25] | 1 | yes | Infected Vero | Lung | Comparison of IHC and ISH; no negative cell controls shown for evaluation; influenza lung as virus disease control |
| 14 | Ray et al. [26] | 1 | no | no | Lung | No validation; no control shown |
| 15 | Roden et al. [27] | 1 | * | no | Lung | Comparison of IHC, RT-qPCR, and ISH |
| 16 | Sauter et al. [28] | 2 | * | no | Lung | Only staining with one antibody is shown |
| 17 | Szabolcs et al. [29] | 7 | yes | Transfected HEK | Lung | Not all antibodies are shown in cell blocks; not all antibodies are shown in lung tissue |
| 18 | Teixeira et al. [30] | 1 | no | no | no | No image shown |

*antibodies were evaluated in former studies by different authors e.g. Liu et al. or Szabolcs et al.

Abbreviation: Nr number of different antibodies that were used in the respective study

**Supplementary Table 2.** Summary of commercially available antibodies against SARS-CoV-2 proteins or double-stranded RNA that were evaluated in this study

| | Antigen | Species | Dilution | Order number | Company |
|---|---|---|---|---|---|
| **1** | Spike | Mouse Mab | 1:300 | #GTX632604 | GeneTex |
| **2** | Spike | Rab Poly | 1:100 | #40150-T62-COV2 | Sino Biological |
| **3** | Spike | Mouse Mab | 1:300 | #ab272420 | Abcam |
| **4** | Nsp3 | Rab Poly | 1:500 | #100-401-A52 | Rockland |
| **5** | Nsp3 | Rab Poly | 1:200 | #GTX135589 | GeneTex |
| **6** | dsRNA | Mouse Mab | 1:200 | #RNT-SCI-10010200 | Jena Bioscience |
| **7** | N | Rab Poly | 1:1000 | #40143-T62 | Sino Biological |

| | | | | | |
|---|---|---|---|---|---|
| **8** | N | Mouse Mab | 1:300 | #40143-MM05 | Sino Biological |
| **9** | N | Mouse Mab | 1:2000 | #HS-452 011 | Synaptic Systems |
| **10** | N | Mouse Mab | 1:500 | #HS-452 111 | Synaptic Systems |
| **11** | N | Rab Mab | 1:5000 | #40143-R001 | Sino Biological |
| **12** | N | Rab Mab | 1:3000 | #A20021 | ABclonal |
| **13** | N | Rab Poly | 1:500 | #BIV-A2061 | Biozol |
| **A** | Spike S1 | Rab Mab | 1:300 | #IHC-184005 | Dianova/Biozol |
| **B** | Spike S1 | Rab Poly | 1:100 | #IHC-184008 | Dianova/Biozol |
| **C** | Spike RBD | Rab Mab | 1:1000 | #IHC-184006 | Dianova/Biozol |

Abbreviations: Spike SARS-CoV-2 spike; N SARS-CoV-2 nucleocapsid; Nsp3 SARS-CoV-2 non-structural protein 3; dsRNA double strand RNA; Rab Rabbit; Mab monoclonal antibody, Poly polyclonal antibody

**Supplementary Table 3.** Recommendations for the use of commercially available antibodies against SARS-CoV-2 proteins or double-stranded RNA (for summary see Supplementary Table 2) for staining in human autopsy tissues

| | **Antigen** | **Cell block** | **Cells** | **Lung** | **Resp. muc.** | **Recommendations** |
|---|---|---|---|---|---|---|
| **1** | Spike | ++ | ++ | ++ | ++ | Suitable for IHC |
| **2** | Spike | + | nd | + | + | Partially recommended for IHC; background staining in several non-COVID-19 tissues |
| **3** | Spike | - | - | - | - | Not recommended |
| **4** | Nsp3 | - | - | - | nd | Not recommended |
| **5** | Nsp3 | - | ++ | - | nd | Works on cells, not suitable for FFPE tissue |
| **6** | dsRNA | + | ++ | - | - | Works on cells, not suitable for FFPE tissue (probably due to insufficient signal to background ratio) |
| **7** | N | ++ | ++ | ++, bg | +++ | Partially recommended for IHC; nonspecific staining of nucleoli in some cell types (e.g. Vero cells, neurons) |
| **8** | N | +++ | +++ | +++ | +++ | Suitable for IHC |
| **9** | N | +++ | +++ | +++ | +++ | Suitable for IHC |
| **10** | N | +++ | +++ | +++ | +++ | Suitable for IHC |
| **11** | N | +++ | nd | ++, bg | +++ | Partially recommended for IHC |
| **12** | N | +, bg | + | +, bg | +, bg | Not recommended; very high background |
| **13** | N | - | nd | nd | nd | Not recommended |
| **A** | Spike S1 | ++ | nd | ++ | nd | Suitable for IHC |
| **B** | Spike S1 | ++ | nd | ++ | nd | Suitable for IHC |
| **C** | Spike RBD | ++ | nd | ++ | nd | Suitable for IHC |

Note, that some antibodies might be partially suited e.g. in specific applications e.g. in double staining where the antibody species matters such as antibody N#7, A-C, which are rabbit antibodies. Some antibodies detect their respective targets in infected cells, but produce very high background in human autopsy tissues.

Abbreviations: Resp. muc. respiratory mucosa; FFPE formalin-fixed paraffin-embedded; Spike SARS-CoV-2 spike; N SARS-CoV-2 nucleocapsid; Nsp3 SARS-CoV-2 non-structural protein 3; dsRNA double stranded RNA; bg background; nd not done

**Supplementary Tables 4a-c.** Summary of cases from Hamburg (2a), Berlin (2b), or Aachen (2c) included in the multi-centre study displaying parameters that might impact the quality of immunohistochemical analyses such as post-mortem interval (PMI) and fixation time of tissues in formalin.

Table 4a. Summary of cases from Hamburg

| Group | Case | Age | Sex | BMI ($kg/m^2$) | Location of death | PM interval (hours) | Time of formaldehyde fixation (hours) | Nasopharyngeal SARS-CoV-2 RNA load (copies/ml) |
|---|---|---|---|---|---|---|---|---|
| COVID-19 | 1 | 70-74 | M | 22.3 | Normal ward | 168 | 1872 | NA |
| COVID-19 | 2 | 95-99 | F | NA | Normal ward | 360 | 816 | $1.3 \times 10^8$ |
| COVID-19 | 3 | 70-74 | M | 23.8 | Normal ward | 144 | 528 | $7.1 \times 10^6$ |
| COVID-19 | 4 | 80-84 | F | 30.0 | Nursing home | 168 | 576 | $1.1 \times 10^7$ |
| COVID-19 | 5 | 60-64 | M | 26.6 | Home | 48 | 36 | $3.9 \times 10^5$ |
| COVID-19 | 6 | 55-59 | F | 36.5 | Home | 48 | 36 | NA |
| COVID-19 | 7 | 80-84 | M | 36.2 | Nursing home | 48 | 36 | $7.5 \times 10^7$ |
| COVID-19 | 8 | 95-99 | M | 26.3 | Normal ward | 120 | 72 | $1.7 \times 10^7$ |
| COVID-19 | 9 | 80-84 | F | NA | Normal ward | 72 | 36 | NA |
| COVID-19 | 10 | 90-94 | M | 21.1 | Nursing home | 72 | 36 | $5.2 \times 10^5$ |
| COVID-19 | 11 | 80-84 | M | 41.8 | ICU | 72 | 36 | $3.8 \times 10^4$ |
| COVID-19 | 12 | 85-89 | F | 30.5 | Normal ward | 72 | 168 | $1.1 \times 10^7$ |
| COVID-19 | 13 | 80-84 | M | 19.3 | ICU | 96 | 48 | NA |
| COVID-19 | 14 | 65-69 | M | 33.5 | Normal ward | 48 | 48 | NA |
| COVID-19 | 15 | 75-79 | M | 33.4 | Normal ward | 72 | 36 | NA |
| COVID-19 | 16 | 70-74 | F | 29.5 | Home | 144 | 528 | $3.4 \times 10^7$ |
| COVID-19 | 17 | 75-79 | M | 27.7 | Normal ward | 48 | 36 | $6.8 \times 10^4$ |
| COVID-19 | 18 | 80-84 | F | 27.0 | Nursing home | 24 | 36 | NA |
| COVID-19 | 19 | 75-79 | M | 26.6 | ICU | 120 | 408 | $1.7 \times 10^4$ |
| COVID-19 | 20 | 75-79 | F | 22.2 | Home | 144 | 36 | $8.4 \times 10^5$ |
| COVID-19 | 21 | 90-94 | M | ND | ICU | 192 | 48 | $1.1 \times 10^8$ |
| COVID-19 | 22 | 45-49 | M | 32.5 | ICU | 48 | 192 | $5.9 \times 10^3$ |
| COVID-19 | 23 | 65-69 | F | 39.6 | Home | 48 | 120 | $9.4 \times 10^3$ |
| COVID-19 | 24 | 80-84 | M | 34.4 | ICU | 72 | 168 | $2.4 \times 10^6$ |
| Normal lung | 25 | 85-89 | M | 25.2 | Other | 120 | 216 | <LOD |
| Normal lung | 26 | 40-44 | F | 24.5 | Home | 24 | 192 | <LOD |
| Normal lung | 27 | 90-94 | M | 20.8 | Home | 120 | 144 | <LOD |
| Normal lung | 28 | 85-89 | M | 27.4 | Home | 96 | 168 | <LOD |
| Normal lung | 29 | 65-69 | M | 24.5 | Home | 96 | 144 | <LOD |
| Normal lung | 30 | 35-39 | M | 28.2 | ICU | 72 | 120 | <LOD |
| Normal lung | 31 | 85-89 | F | 18.5 | Normal ward | 48 | 72 | <LOD |

Abbreviations: F female; M male; BMI body mass index; PM post mortem; ICU intensive care unit; NA not assessed; LOD limit of detection

Table4b. Summary of cases from Berlin

| Group | Case | Age | Sex | BMI (kg/m$^2$) | Location of death | PM interval (hours) | Time of formaldehyde fixation (hours) | Nasopharyngeal SARS-CoV-2 RNA load (copies/ml) |
|---|---|---|---|---|---|---|---|---|
| COVID-19 | B1 | 75-79 | M | 30.9 | ICU | 30 | 9 months! | NA |
| COVID-19 | B2 | 90-94 | M | 27 | Normal ward | 72 | 24 | 6.3 x $10^7$ |
| COVID-19 | B3 | 75-79 | F | 50.4 | ICU | 148 | 24 | NA |
| COVID-19 | B4 | 80-84 | M | ND | Normal ward | 82 | 24 | 2.7 x $10^3$ |
| ARDS | B5 | 70-74 | M | ND | ICU | ND | 24 | NA |
| ARDS | B6 | 80-84 | M | 27.4 | ICU | 35 | 24 | <LOD |

Abbreviations: F female; M male; BMI body mass index; PM post mortem; ICU intensive care unit; NA not assessed

Table 4c. Summary of cases from Aachen

| Group | Case | Age | Sex | BMI (kg/m$^2$) | Location of death | PM interval (hours) | Time of formaldehyde fixation (hours) | SARS-CoV-2 RNA load lung FFPE (copies/ml) |
|---|---|---|---|---|---|---|---|---|
| ARDS | A1 | 76-80 | M | 26.3 | ICU | 15 | 108 | NA |
| ARDS | A2 | 71-75 | F | 25.1 | Intermediate care | 109 | 56 | NA |
| ARDS | A3 | 51-55 | F | 29.3 | Intermediate care | 11 | 60 | NA |
| ARDS | A4 | 51-55 | F | >40 | Intermediate care | 36 | 32 | NA |
| ARDS | A5 | 66-70 | M | 29.3 | Intermediate care | 107 | 128 | NA |
| ARDS | A6 | 56-60 | F | 32.1 | ICU | 19 | 36 | NA |
| ARDS | A7 | 51-55 | M | 27.7 | Intermediate care | 16 | 108 | NA |
| ARDS | A8 | 51-55 | M | 29.2 | ICU | 35 | 104 | NA |
| COVID-19 | A9 | 56-60 | M | 25.1 | ICU | 24 | 320 | 12.7 |
| COVID-19 | A10 | 51-55 | M | 23 | ICU | 41 | 320 | 180.6 |
| COVID-19 | A11 | 56-60 | M | 36.1 | ICU | 183 | 224 | 199.9 |
| COVID-19 | A12 | 71-75 | M | 23.7 | Intermediate care | 19 | 176 | 651.9 |
| COVID-19 | A13 | 66-70 | F | 36.2 | ICU | 74 | 200 | 3.7 x $10^3$ |
| COVID-19 | A14 | 71-75 | F | 27 | ICU | 45 | 228 | 5.9 x $10^3$ |
| COVID-19 | A15 | 81-85 | M | 27 | ICU | 20 | 252 | 7.3 x $10^4$ |
| COVID-19 | A16 | 56-60 | M | 24.7 | ICU | 24 | 275 | 1.6 x $10^3$ |
| COVID-19 | A17 | 76-80 | M | 31.6 | Intermediate care | 29 | 275 | 6.3 x $10^4$ |
| COVID-19 | A18 | 66-70 | M | 28.7 | ICU | 30 | 300 | 235.3 |
| COVID-19 | A19 | 71-75 | F | 27 | ICU | 42 | 180 | 82.1 |
| COVID-19 | A20 | 61-65 | F | 23.8 | ICU | 48 | 275 | 2.0 |
| COVID-19 | A21 | 51-55 | F | 40.6 | ICU | 28 | 107 | 3.1 x $10^5$ |
| COVID-19 | A22 | 66-70 | M | 30.7 | ICU | 17 | 180 | NA |
| Influenza (H1N1) | A23 | 31-35 | F | 23.1 | ICU | 31 | 104 | NA |
| Influenza (H1N1) | A24 | 51-55 | M | >40 | ICU | 13 | 83 | NA |
| Influenza (H1N1) | A25 | 41-45 | F | 26.6 | Intermediate care | 12 | 131 | NA |
| Influenza (seasonal Type A) | A26 | 61-65 | F | 21 | Intermediate care | 34 | 275 | NA |
| Influenza (seasonal Type B) | A27 | 31-35 | F | 32.3 | ICU | 36 | 59 | NA |

| | | | | | | | | |
|---|---|---|---|---|---|---|---|---|
| Influenza (seasonal Type B) | A28 | 71-75 | F | 26.4 | Intermediate care | 18 | 35 | NA |
| Influenza (seasonal Type A) | A29 | 56-60 | M | 37 | Intermediate care | 15 | 35 | NA |
| Normal lung | A30 | 56-60 | F | 20.9 | Intermediate care | 24 | 59 | NA |
| Normal lung | A31 | 86-90 | M | 24.6 | Standard care | 30 | 83 | NA |
| Normal lung | A32 | 85-90 | M | 27.2 | Intermediate care | 68 | 59 | NA |
| Normal lung | A33 | 61-65 | M | 30.2 | ICU | 23 | 107 | NA |
| Normal lung | A34 | 56-60 | M | 26.9 | ICU | 65 | 59 | NA |

Abbreviations: F female; M male; BMI body mass index; PM post mortem; ICU intensive care unit; NA not assessed; FFPE formalin-fixed paraffin-embedded

**Supplementary Table 5.** Hamburg Berlin Aachen, Paris Summary of the multicenter evaluation of SARS-CoV-2 nucleocapsid staining (antibody N#9) that was set up to test the validity of SARS-Co V-2 staining recommendations

| | Center I | | Center II | | | Center III | Center IV | | | | | | | |
|---|---|---|---|---|---|---|---|---|---|---|---|---|---|---|
| Case | Path I | Path II | Path I | Path II | Path III | Path I | Path I | RT-qPCR ct value | mean score | agreement all | agreement exp only | RT-qPCR +/- | IHC +/- | FP/FN |
| 0 | 0 | 0 | 0 | 0 | 1 | 1 | 0 | 29,87 | 0,29 | 0 | 0 | 1 | 0 | FN |
| 2 | 2 | 2 | 3 | 3 | 2 | 3 | 2,5 | 21,2 | 2,50 | 1 | 1 | 1 | 1 | TP |
| 3 | 0 | 0 | 0 | 0 | 0 | 0 | 0 | xx | 0,00 | 1 | 1 | 0 | 0 | TN |
| 4 | 1 | 1 | 1 | 1 | 1 | 1 | 1 | 20,94 | 1,00 | 1 | 1 | 1 | 1 | TP |
| 5 | 1 | 1 | 0 | 1 | 1 | 0 | 0 | 24,67 | 0,57 | 0 | 0 | 1 | x | x |
| 6 | 0 | 0 | 0 | 0 | 0 | 0 | 0 | 31,25 | 0,00 | 1 | 1 | 1 | 0 | FN |
| 7 | 3 | 3 | 3 | 3 | 3 | 3 | 2,5 | 16,48 | 2,93 | 1 | 1 | 1 | 1 | TP |
| 8 | 3 | 3 | 3 | 3 | 3 | 3 | 3 | 19,9 | 3,00 | 1 | 1 | 1 | 1 | TP |
| 9 | 1 | 1 | 1 | 2 | 1 | 1 | 1 | 25,07 | 1,14 | 1 | 1 | 1 | 1 | TP |
| 10 | 2 | 2 | 2 | 3 | 3 | 2 | 2 | 17,39 | 2,29 | 1 | 1 | 1 | 1 | TP |
| 11 | 1 | 1 | 1 | 2 | 2 | 2 | 2 | 21,84 | 1,57 | 1 | 1 | 1 | 1 | TP |
| 12 | 3 | 2 | 3 | 3 | 3 | 3 | 2,5 | 24,31 | 2,79 | 1 | 1 | 1 | 1 | TP |
| 13 | 0 | 0 | 0 | 0 | 0 | 0 | 0 | xx | 0,00 | 1 | 1 | 0 | 0 | TN |
| 14 | 1 | 1 | 1 | 1 | 1 | 1 | 1 | xx | 1,00 | 1 | 1 | 0 | 1 | FP |
| 15 | 2 | 1 | 1 | 2 | 2 | 2 | 2 | 19,22 | 1,71 | 1 | 1 | 1 | 1 | TP |
| 16 | 1 | 0 | 1 | 1 | 1 | 1 | 1 | 30,1 | 0,86 | 0 | 0 | 1 | 1 | TP |
| 17 | 1 | 1 | 1 | 2 | 2 | 2 | 1 | 29,1 | 1,43 | 1 | 1 | 1 | 1 | TP |
| 18 | 2 | 2 | 3 | 3 | 3 | 3 | 3 | 20,78 | 2,71 | 1 | 1 | 1 | 1 | TP |
| 19 | 0 | 0 | 0 | 0 | 0 | 0 | 0 | xx | 0,00 | 1 | 1 | 0 | 0 | TN |
| 20 | 0 | 0 | 0 | 1 | 0 | 0 | 0 | xx | 0,14 | 0 | 0 | 0 | 0 | TN |
| 21 | 1 | 1 | 1 | 2 | 2 | 3 | 1,5 | 20,25 | 1,64 | 1 | 1 | 1 | 1 | TP |
| 22 | 0 | 0 | 0 | 0 | 0 | 0 | 0 | xx | 0,00 | 1 | 1 | 0 | 0 | TN |
| 23 | 3 | 2 | 3 | 3 | 3 | 3 | 2,5 | 20,6 | 2,79 | 1 | 1 | 1 | 1 | TP |
| 24 | 0 | 0 | 0 | 0 | 0 | 0 | 0 | 31,41 | 0,00 | 1 | 1 | 1 | 0 | FN |
| 25 | 0 | 0 | 0 | 0 | 0 | 0 | 0 | xx | 0,00 | 1 | 1 | 0 | 0 | TN |
| 26 | 0 | 0 | 0 | 0 | 0 | 1 | 0 | xx | 0,14 | 0 | 1 | 0 | 0 | TN |
| 27 | 0 | 0 | 0 | 0 | 0 | 1 | 0 | xx | 0,14 | 0 | 1 | 0 | 0 | TN |
| 28 | 0 | 0 | 0 | 0 | 0 | 1 | 0 | xx | 0,14 | 0 | 1 | 0 | 0 | TN |
| 29 | 0 | 0 | 0 | 0 | 0 | 1 | 0 | xx | 0,14 | 0 | 1 | 0 | 0 | TN |
| 30 | 0 | 0 | 0 | 0 | 0 | 1 | 0 | xx | 0,14 | 0 | 1 | 0 | 0 | TN |
| 31 | 0 | 0 | 0 | 0 | 0 | 0 | 0 | xx | 0,00 | 1 | 1 | 0 | 0 | TN |
| B1 | 2 | 2 | 2 | 2 | 2 | 2 | 1,5 | 22,51 | 1,93 | 1 | 1 | 1 | 1 | TP |
| B2 | 0 | 0 | 0 | 0 | 0 | 0 | 0 | 0,00 | 0,00 | 1 | 1 | 0 | 0 | TN |
| B3 | 1 | 1 | 1 | 1 | 0 | 0 | 1 | 31,22 | 0,71 | 0 | 0 | 1 | 1 | TP |
| B4 | 3 | 3 | 3 | 3 | 3 | 3 | 2,5 | 18,72 | 2,93 | 1 | 1 | 1 | 1 | TP |
| B5 | 1 | 1 | 0 | 0 | 0 | 3 | 0 | xx | 0,71 | 0 | 0 | 0 | x | x |
| B6 | 0 | 0 | 0 | 0 | 0 | 0 | 0 | xx | 0,00 | 1 | 1 | 0 | 0 | TN |
| A1 | 0 | 0 | 0 | 0 | 0 | 0 | 0 | 0,00 | 0,00 | 1 | 1 | 0 | 0 | TN |
| A2 | 0 | 0 | 0 | 0 | 0 | 1 | 0 | 0,00 | 0,14 | 0 | 1 | 0 | 0 | TN |
| A3 | 0 | 0 | 0 | 0 | 0 | 0 | 0 | 0,00 | 0,00 | 1 | 1 | 0 | 0 | TN |
| A4 | 0 | 0 | 0 | 0 | 0 | 2 | 0 | 0,00 | 0,29 | 0 | 1 | 0 | 0 | TN |
| A5 | 0 | 0 | 0 | 0 | 0 | 0 | 0 | 0,00 | 0,00 | 1 | 1 | 0 | 0 | TN |
| A6 | 0 | 0 | 0 | 0 | 0 | 1 | 0 | 0,00 | 0,14 | 0 | 1 | 0 | 0 | TN |
| A7 | 0 | 0 | 0 | 0 | 0 | 2 | 0 | 0,00 | 0,29 | 0 | 1 | 0 | 0 | TN |
| A8 | 0 | 0 | 0 | 0 | 0 | 0 | 0 | 0,00 | 0,00 | 1 | 1 | 0 | 0 | TN |
| A9 | 0 | 0 | 0 | 0 | 0 | 0 | 0 | 36,28 | 0,00 | 1 | 1 | 0 | 0 | TN |
| A10 | 0 | 0 | 0 | 0 | 0 | 0 | 0 | 32,37 | 0,00 | 1 | 1 | 0 | 0 | TN |
| A11 | 0 | 1 | 0 | 0 | 0 | 0 | 0 | 32,22 | 0,14 | 0 | 0 | 0 | 0 | TN |
| A12 | 0 | 0 | 0 | 0 | 0 | 0 | 0 | 30,48 | 0,00 | 1 | 1 | 1 | 0 | FN |
| A13 | 0 | 0 | 0 | 0 | 0 | 0 | 0 | 27,92 | 0,00 | 1 | 1 | 1 | 0 | FN |
| A14 | 0 | 1 | 0 | 0 | 0 | 0 | 0 | 27,23 | 0,14 | 0 | 0 | 1 | 0 | FN |
| A15 | 2 | 2 | 1 | 1 | 1 | 1 | 1 | 23,54 | 1,29 | 1 | 1 | 1 | 1 | TP |
| A16 | 0 | 0 | 0 | 0 | 0 | 0 | 0 | 29,15 | 0,00 | 1 | 1 | 1 | 0 | FN |
| A17 | 1 | 1 | 0 | 0 | 0 | 0 | 0 | 23,73 | 0,29 | 0 | 0 | 1 | 0 | FN |
| A18 | 0 | 0 | 0 | 0 | 0 | 1 | 0 | 31,98 | 0,14 | 0 | 1 | 1 | 1 | TP |
| A19 | 0 | 0 | 0 | 0 | 0 | 0 | 0 | 33,53 | 0,00 | 1 | 1 | 0 | 0 | TN |
| A20 | 0 | 0 | 0 | 0 | 0 | 0 | 0 | 39,01 | 0,00 | 1 | 1 | 0 | 0 | TN |
| A21 | 3 | 3 | 3 | 2 | 3 | 3 | 2 | 21,42 | 2,71 | 0 | 1 | 1 | 1 | TP |
| A22 | 0 | 0 | 0 | 0 | 0 | 0 | 0 | NA | 0,00 | 1 | 1 | 0 | 0 | TN |
| A23 | 0 | 1 | 0 | 0 | 0 | 1 | 0 | NA | 0,29 | 0 | 0 | 0 | 0 | TN |
| A24 | 0 | 0 | 0 | 0 | 0 | 0 | 0 | NA | 0,00 | 1 | 1 | 0 | 0 | TN |
| A25 | 0 | 0 | 0 | 0 | 0 | 1 | 0 | NA | 0,14 | 0 | 1 | 0 | 0 | TN |
| A26 | 0 | 0 | 0 | 0 | 0 | 0 | 0 | NA | 0,00 | 1 | 1 | 0 | 0 | TN |
| A27 | 0 | 0 | 0 | 0 | 0 | 1 | 0 | NA | 0,14 | 0 | 1 | 0 | 0 | TN |
| A28 | 0 | 0 | 0 | 0 | 0 | 0 | 0 | NA | 0,00 | 1 | 1 | 0 | 0 | TN |
| A29 | 0 | 0 | 0 | 0 | 0 | 1 | 0 | NA | 0,14 | 0 | 1 | 0 | 0 | TN |
| A30 | 0 | 0 | 0 | 0 | 0 | 1 | 0 | NA | 0,14 | 0 | 1 | 0 | 0 | TN |
| A31 | 0 | 0 | 0 | 0 | 0 | 0 | 0 | NA | 0,00 | 1 | 1 | 0 | 0 | TN |
| A32 | 0 | 0 | 0 | 0 | 0 | 0 | 0 | NA | 0,00 | 1 | 1 | 0 | 0 | TN |
| A33 | 0 | 1 | 0 | 0 | 0 | 2 | 0 | NA | 0,43 | 0 | 0 | 0 | 0 | TN |
| A34 | 0 | 1 | 0 | 0 | 0 | 2 | 0 | NA | 0,43 | 0 | 0 | 0 | 0 | TN |

Experienced investigators/pathologists per institute independently scored the stained lung tissues in a blinded fashion.
(0) No positive cells; (1) single positive cells; (2) several positive cells and at different locations within the stained tissue; (3) widespread positive staining in different cells and all over the stained tissue, Abbreviations: Path evaluating pathologist/ neuropathologist; exp expert; NA not available; xx RT-qPCR ct value >32 and was considered negative; X not classifiable; TN True negative; TP True positive; FN False negative; FP False Positive; 0 no agreement; 1 agreement

**Supplementary Table 6**. Overview of the human samples analyzed by electron microscopy

| EM case nr. | Tissue | PMI | RT-qPCR | Preparation performed by institute | Analysis performed by institute | Definitive virus found |
|---|---|---|---|---|---|---|
| 1 | Lung | 30 | +++ (6.7)* | RKI | RKI and NP Charité | Yes |
| 1 | Lung | 30 | +++ (6.7)* | NP Charité | NP Charité | Yes*** |
| 2 | Olfactory mucosa | 82 | +++ (8)* | RKI | RKI | Yes |
| 3 | Olfactory mucosa | 17 | + (2.39)* | NP Charité | NP Charité | No |
| 4 | Lung | 106 | +++ (6.14)* | NP Charité | NP Charité | No**** |
| 5 | Lung** | 30 | + (1.72)* | NP Charité | NP Charité | No |
| 5 | Medulla obl.** | 30 | + (2.03)* | NP Charité | NP Charité | No |
| 6 | Lung** | 10 | + (3.81)* | NP Charité | NP Charité | No |
| 7 | Lung | 24 | ++ | P Aachen | P Aachen | No |
| 7 | Kidney | 24 | NA | P Aachen | P Aachen | No |
| 8 | Lung | 24 | +++ | P Aachen | P Aachen | No |
| 8 | Kidney | 24 | + | P Aachen | P Aachen | No |
| 9 | Lung | 19 | ++ | P Aachen | P Aachen | No |
| 9 | Kidney | 19 | + | P Aachen | P Aachen | No |
| 10 | Lung | 29 | +++ | P Aachen | P Aachen | No |
| 10 | Kidney | 29 | NA | P Aachen | P Aachen | No |
| 11 | Lung | 28 | + | P Aachen | P Aachen | No |
| 11 | Kidney | 28 | NA | P Aachen | P Aachen | No |
| 12 | Lung | 38 | + | P Aachen | P Aachen | No |
| 12 | Kidney | 38 | NA | P Aachen | P Aachen | No |
| 13 | Lung | 48 | + | P Aachen | P Aachen | No |
| 13 | Kidney | 48 | NA | P Aachen | P Aachen | No |
| 13 | Kidney tx | 48 | NA | P Aachen | P Aachen | No |
| 14 | Lung | 28 | +++ | P Aachen | P Aachen | No |
| 14 | Trachea | 28 | NA | P Aachen | P Aachen | No |
| 15 | Lung | 17 | NA | P Aachen | P Aachen | No |
| 15 | Myocardium | 17 | NA | P Aachen | P Aachen | No |
| 16 | Kidney | 31 | NA | P Aachen | P Aachen | No |
| 16 | Myocardium | 31 | NA | P Aachen | P Aachen | No |

*) For NP Charité/RKI samples, results of the RT-qPCR were, beside the ct value, additionally calculated as $Log_{10}$ SARS-CoV-2 RNA copies/ DNA amount of 10,000 diploid nuclei; **) Large-scale datasets of these samples were published in a previous paper [4]; ***) In one digitized ultrathin section, we found several dozens of extracellular coronavirus particles (not included in the repository datasets); ****) Only one ultrathin section was digitized due to the high post-mortem interval.
Abbreviations: EM electron microscopy; RKI Robert Koch Institute; NP Charité Department of Neuropathology, Charité – Universitätsmedizin Berlin; Medulla obl. Medulla oblongata; P Aachen Department of Pathology, RWTH Aachen University Faculty of Medicine; PMI post-mortem interval

**Supplementary Table 7**. Overview of the 15 SARS-CoV-2 infected cells of the autopsy lung tissue

| Nr. | Cell type* | Total area ($\mu m^2$) | Area nucl. ($\mu m^2$) | Area cytopl. ($\mu m^2$) | CoV total | CoV intr. | Intr. CoV per $\mu m^2$ | CoV 1 | CoV 2 | CoV 3 | CoV 4 | Comment |
|---|---|---|---|---|---|---|---|---|---|---|---|---|
| 1 | NA | 93.24 | 19.75 | 73.49 | 71 | 71 | 0.97 | 47 | 16 | 8 | 0 | |
| 2 | Type 2 pneumocyte | 94.14 | 11.47 | 82.67 | 114 | 112 | 1.35 | 70 | 16 | 26 | 2 | Viroplasm; tubular structures (compare Goldsmith et al. 2004 [12]) |
| 3 | Type 2 pneumocyte | 131.28 | 21.88 | 109.40 | 69 | 66 | 0.60 | 44 | 20 | 2 | 3 | |

| | | | | | | | | | | | |
|---|---|---|---|---|---|---|---|---|---|---|---|
| 4 | Probably alveolar macrophage | 35.66 | 0.00 | 35.66 | 32 | 32 | 0.90 | 26 | 6 | 0 | 0 | |
| 5 | Type 2 pneumocyte | 150.75 | 63.18 | 87.57 | 29 | 29 | 0.33 | 24 | 5 | 0 | 0 | |
| 6 | Alveolar macrophage | 178.12 | 20.26 | 157.85 | 387 | 358 | 2.27 | 225 | 90 | 43 | 29 | |
| 7 | Alveolar macrophage | 127.33 | 13.40 | 113.93 | 620 | 620 | 5.44 | 410 | 116 | 94 | 0 | Viroplasm; CoV-containing vesicle with dark granular material (compare Goldsmith et al. 2004 [12]) |
| 8 | NA | 7.44 | 0.00 | 7.44 | 18 | 18 | 2.42 | 15 | 2 | 1 | 0 | Viroplasm; tubular structures |
| 9 | Epithelial cell, very likely type 2 pneumocyte | 145.79 | 23.24 | 122.55 | 61 | 60 | 0.49 | 37 | 16 | 7 | 1 | Viroplasm; multiple well-preserved small CoV-containing vesicles within granular, ribosome-like material (compare Goldsmith et al. 2004 [12]) |
| 10 | Probably epithelial cell, perhaps intermediate cell in transition from type 2 into type 1 | 181.38 | 79.84 | 101.53 | 162 | 104 | 1.02 | 73 | 26 | 5 | 58 | |
| 11 | NA | 56.57 | 0.00 | 56.57 | 4 | 4 | 0.07 | 4 | 0 | 0 | 0 | |
| 12 | Epithelial cell, perhaps intermediate cell in transition from type 2 into type 1 | 16.69 | 0.00 | 16.69 | 17 | 11 | 0.66 | 8 | 1 | 2 | 6 | Viroplasm; tubular structures |
| 13 | NA | 50.10 | 23.56 | 26.54 | 65 | 22 | 0.83 | 14 | 6 | 2 | 43 | |
| 14 | Probably alveolar macrophage | 56.60 | 13.00 | 43.60 | 18 | 18 | 0.41 | 15 | 1 | 2 | 0 | |
| 15 | Alveolar macrophage | 55.25 | 8.95 | 46.30 | 34 | 32 | 0.69 | 16 | 10 | 6 | 2 | Viroplasm; tubular structures |

*) Limited due to autolysis. Abbreviations: CoV coronavirus; CoV 1-4 coronavirus type 1-4; cytopl. cytoplasm; intr. intracellular; NA not available due to small size or too severe autolytic changes; nucl. nucleus; S sample; SARS-CoV-2 severe acute respiratory syndrome coronavirus 2

**Supplementary Table 8**. Publications in scientific journals demonstrating questionable ultrastructural evidence of coronavirus particles (SARS-CoV-2) in human samples, April 2020-November 2021

| Nr. | Reference | Tissue | Figure | EM-image nr. | Reported diagnosis | Our morphological description | Possible identity of the structures | Structural preservation | Image presentation/ quality | All relevant structural features of CoV | Response | Bullock et al. | Hop-fer et al. | Comment |
|---|---|---|---|---|---|---|---|---|---|---|---|---|---|---|
| 1 | Abbate et al. [31] | Kidney | 1 | 1 | Coronavirus | Coated vesicle, cytoplasm | CCV | sufficient | sufficient | no | NA | x | o | |
| 2 | Abdullaev et al. [32] | Lung | 5B | 2 | Coronavirus particles | Structures within membrane compartment | Unidentifiable structure/s | insufficient | insufficient | no | NA | o | o | |
| 2 | Abdullaev et al. | Lung | 5C | 3 | Coronavirus particles | Vesicular structures with electron dense dots | Unidentifiable structure/s | sufficient* | sufficient | no | NA | o | o | |
| 2 | Abdullaev et al. | Lymph node | 5D to F | 4 | SARS-CoV-2 virions | Vesicular structures within membrane compartment | Mitochondrion /ia | insufficient | insufficient | no | NA | o | o | |
| 2 | Abdullaev et al. | Lymph node | 5G to I | 5 | Coronavirus particles | Vesicular structures in membrane compartment | Unidentifiable structure/s | insufficient | insufficient | no | NA | o | o | |
| 3 | Achua et al. [33] | Testis | 1A | 6 | Coronavirus-like spiked viral particles | Dark particles on membranes | rER | sufficient | sufficient | no | NA | o | o | |
| 3 | Achua et al. | Testis | 1B and 2B | 7 | Coronavirus-like spiked viral particles, coronavirus-like particles | Coated vesicles, cytoplasm | CCV | sufficient | sufficient | no | NA | o | o | |
| 4 | Ackermann et al. [34] | Lung | 3D | 8 | SARS-CoV-2 | Dark circular structures, cytoplasm | Unidentifiable structure/s | insufficient | insufficient | no | Scholkmann and Nicholls [35] Ackermann et al. [36] (reply to Scholkmann and Nicholls) Miller [37] (letter) Ackermann et al. [38] (reply to Miller) | x | x | |
| 5 | Ackermann et al. (reply to Scholkmann and Nicholls) [36] | Lung | 1A and B | 9 | Viruslike particles | Circular particles within membrane structures | Unidentifiable structure/s | sufficient | insufficient | no | NA | o | x | |
| 6 | Albert et al. [39] | Heart | 3A and B | 10 | A viral particle of coronavirus, viral particle | Coated vesicle, cytoplasm | CCV | sufficient | insufficient | no | NA | o | o | |

| | | | | | | | | | | | | | | |
|---|---|---|---|---|---|---|---|---|---|---|---|---|---|---|
| 6 | Albert et al. | Heart | 4A to D | 11 | Viral particles, viral particle | Coated vesicle, cytoplasm | CCV | sufficient | insufficient | no | NA | o | o | |
| 7 | Algarroba et al. [40] | Placenta | 2 and 3 | 12 | Virion | Coated vesicles, cytoplasm | CCV | sufficient | sufficient | no | Kniss [41] Algarroba et al. [42] (reply to Kniss) Algarroba et al. [43] (letter) | x | x | |
| 7 | Algarroba et al. | Placenta | 4 | 13 | Virion | Coated vesicles, cytoplasm | CCV | sufficient | sufficient | no | see above | x | x | |
| 7 | Algarroba et al. | Placenta | 5 | 14 | Virion | Coated vesicles, cytoplasm | CCV | sufficient | sufficient | no | see above | x | x | |
| 7 | Algarroba et al. | Placenta | 6 | 15 | Virions | Coated vesicles, cytoplasm | CCV | sufficient | sufficient | no | see above | x | x | |
| 8 | Algarroba et al. (reply to Kniss) [42] | Placenta | 1A | 16 | Virions | Vesicles of different sizes | Unidentifiable structure/s | sufficient* | insufficient | no | Algarroba et al. (letter) [43] | o | x | |
| 8 | Algarroba et al. (reply to Kniss) | Placenta | 1B | 17 | Virions | Vesicular structure | Unidentifiable structure/s | sufficient* | insufficient | no | NA | o | x | |
| 8 | Algarroba et al. (reply to Kniss) | Placenta | 1C | 18 | Virions | Coated vesicles (right) and vesicular structure (left) | CCV | sufficient* | sufficient | no | NA | o | x | |
| 8 | Algarroba et al. (reply to Kniss) | Placenta | 3 | 19 | Clusters of 10 nm immunogold particles (arrows) cross-reacting with the SARS-CoV-2 spike glycoprotein antibody | Small clusters of dark, round particles (immunogold) | Immunogold, unidentifiable structure/s | insufficient | insufficient | no | NA | o | x | |
| 9 | Anandh et al. [44] | Kidney | 3 | 20 | Viral particles | Vesicular structures with relatively empty interior and surface projections | Unidentifiable structure/s | sufficient | sufficient | no | NA | o | o | |
| 10 | Araujo-Silva et al. [45] | Retina | 2B | 21 | Presumed viral particles | Vesicular structures, one in membrane compartment | Mitochondrion /ia | insufficient* | insufficient | no | NA | o | o | Structure on the right side probably an autolytic mitochondrion, on the left unidentifiable structure |
| 10 | Araujo-Silva et al. | Retina | 2C | 22 | Viral particles | Vesicular structures | Unidentifiable structure/s | insufficient* | sufficient | no | NA | o | o | |
| 10 | Araujo-Silva et al. | Retina | 3A | 23 | Presumed viral particles, double membrane particles | Rounded structure with bright center | Unidentifiable structure/s | insufficient* | insufficient | no | NA | o | o | Misleading additional description as "double membrane particles" |
| 10 | Araujo-Silva et al. | Retina | 3B | 24 | Presumed viral particles, double membrane particles | Multiple vesicular structures | Unidentifiable structure/s | insufficient* | insufficient | no | NA | o | o | Misleading additional description as "double membrane particles" |

| | | | | | | | | | | | | | | |
|---|---|---|---|---|---|---|---|---|---|---|---|---|---|---|
| 10 | Araujo-Silva et al. | Retina | 3C | 25 | Presumed viral particles, particles with dense electron grain | Multiple round structures | Unidentifiable structure/s | insufficient* | insufficient | no | NA | o | o | Misleading additional description as "double membrane particles" |
| 10 | Araujo-Silva et al. | Retina | 3D | 26 | Presumed viral particles, particles with dense electron grain inside | Vesicular and roudn structure | Unidentifiable structure/s | insufficient* | insufficient | no | NA | o | o | |
| 11 | Bain et al. [46] | ETA | 1B | 27 | Presumptive SARS-CoV-2 virion | Vesicular structure with surface structures and granular interior | Unidentifiable structure/s | sufficient* | sufficient | no | NA | o | o | |
| 11 | Bain et al. | ETA | 1C | 28 | Presumptive SARS-CoV-2 virions | Vesicular structures | Unidentifiable structure/s | sufficient* | insufficient | no | NA | o | o | |
| 11 | Bain et al. | ETA | 1D | 29 | CD14 (…) surface immunostaining and internal immunostaining of SARS-CoV-2 Nucleocapsid protein | Dark particles (immunogold double labeling) on granular background | Immunogold, unidentifiable structure/s | insufficient | insufficient | no | NA | o | o | |
| 12 | Bian et al. [13] | Nasopharynges, trachea, lungs, heart, blood vessels, spleen, small intestine, kidneys, ovary, testes | NA (no electron micrographs shown) | NA | Coronavirus particles | NA | NA | NA | NA | NA | NA | o | o | |
| 13 | Birkhead et al. [47] | Placenta | Left | 30 | Virions | Multiple vesicular structures with electron dense, partly granular interior, within membrane comparment | Unidentifiable structure/s | sufficient | sufficient | no* | NA | o | o | *) Some features of coronavirus, for FFPE-reembedded material a bit too less compact, could also be rER vesicles (compare Werion et al. and Hopfer et al. [48 11]) |

| | | | | | | | | | | | | | | |
|---|---|---|---|---|---|---|---|---|---|---|---|---|---|---|
| 13 | Birkhead et al. | Placenta | Right | 31 | Virions | Multiple vesicular structures with electron dense, partly granular interior, within membrane comparment | Unidentifiable structure/s | sufficient | sufficient | no* | NA | o | o | *) Some features of coronavirus, for FFPE-reembedded material a bit too less compact, could also be rER vesicles (compare Werion et al. and Hopfer et al. [48] [11]) |
| 14 | Bojkova et al. [49] | Heart | 4G | 32 | Putative viral particles | Coated vesicles, cytoplasm | CCV | sufficient | insufficient | no | NA | o | o | |
| 15 | Borczuk et al. [15] | Lung | 6E | 33 | Putative viral particles | Dark particles on membranes | rER | insufficient | insufficient | no | NA | x | o | |
| 15 | Borczuk et al. | Lung | 7F | 34 | Viral particle | Coated vesicular structure | Unidentifiable structure/s | sufficient | insufficient | no | NA | x | o | |
| 16 | Bosmüller et al. [50] | Lung | 4B (+inset left) | 35 | Virus particles | Vesicular structure | Unidentifiable structure/s | insufficient | insufficient | no | NA | o | x | |
| 16 | Bosmüller et al. | Lung | 4B (right; inset*) | 36 | Virus particles, viruses | Round to oval vesicular structures with fuzzy surface within membrane structure | Mitochondrion/ia | insufficient | insufficient | no | NA | o | x | *) The inset shows another region |
| 16 | Bosmüller et al. | Lung | S2 | 37 | Virus particles | Round, dark structure | Unidentifiable structure/s | insufficient | insufficient | no | NA | o | x | |
| 17 | Bradley et al. [51] | Trachea | 5A | 38 | Coronavirus-like particles termed “virus particles” | Vesicular structures | Unidentifiable structure/s | insufficient | insufficient | no | Dittmayer et al.* [5] | x | x | *) Other publications were also discussed |
| 17 | Bradley et al. | Trachea | 5B | 39 | Coronavirus-like particles termed “virus particles” | Coated vesicles | CCV | insufficient | insufficient | no | see above | x | x | |
| 17 | Bradley et al. | Lung | 5C | 40 | Coronavirus-like particles termed “virus particles” | Vesicles in vesicular structure | MVB | insufficient | insufficient | no | see above | x | x | |
| 17 | Bradley et al. | Lung | 5D | 41 | Coronavirus-like particles termed “virus particles” | Circular particles within membrane structure | Unidentifiable structure/s | insufficient | insufficient | no | see above | x | x | |
| 17 | Bradley et al. | Intestine | 5E | 42 | Coronavirus-like particles termed “virus particles” | Vesicular structures | Unidentifiable structure/s | insufficient | insufficient | no | see above | x | x | |
| 17 | Bradley et al. | Intestine | 5F | 43 | Coronavirus-like particles termed “virus particles” | Vesicular structures | Unidentifiable structure/s | insufficient | insufficient | no | see above | x | x | |
| 17 | Bradley et al. | Kidney | 5G | 44 | Coronavirus-like particles termed “virus particles” | Vesicular structures with surface projections | CCV | insufficient | insufficient | no | see above | x | x | |
| 17 | Bradley et al. | Kidney | 5H | 45 | Coronavirus-like particles termed “virus particles” | Vesicular structures with surface projections | Unidentifiable structure/s | insufficient | insufficient | no | see above | x | x | |

| | | | | | | | | | | | | | | |
|---|---|---|---|---|---|---|---|---|---|---|---|---|---|---|
| 18 | Bryce et al. [52] | Lung | 1I | 46 | Virus particles | Round to oval objects with surface projections | Cell processes | sufficient | sufficient | no | NA | o | o | |
| 18 | Bryce et al. | Lymph node | 2F | 47 | Virus particles | Round particle with tiny surface structures and core-like interior | Unidentifiable structure/s | sufficient | sufficient | no | NA | o | o | |
| 19 | Buja et al. [53] | Kidney | 12B | 48 | Viral particles | Vesicular structures within membrane structure | MVB | insufficient | insufficient | no | Giannico and Miller [54] Buja [55] (reply to Giannico and Miller) | o | o | |
| 19 | Buja et al. | Kidney | 12C | 49 | Viral particle | Vesicular structure with surface projections | Unidentifiable structure/s | insufficient | insufficient | no | see above | o | o | |
| 20 | Bulfamante et al. (brain) [56] | Medulla oblong ata | 1B and C | 50 | A spherical particle with size suspicious for a viral particle, morphology is compatible to that of SARS-CoV-2 | Vesicular structure next to myelin sheath, or protrusion | Unidentifiable structure/s | insufficient | insufficient | no | NA | o | o | |
| 20 | Bulfamante et al. (brain) | Gyrus rectus | 1D and E | 51 | Viral-like particle | Dark particles | Unidentifiable structure/s | insufficient | insufficient | no | NA | o | o | |
| 21 | Bulfamante et al. (heart) [57] | Heart | 5A and B | 52 | SARS-CoV-2 virions, viral particles | Vesicular structure with empty lumen | Unidentifiable structure/s | insufficient | insufficient | no | NA | o | o | |
| 21 | Bulfamante et al. (heart) | Heart | 5C and D | 53 | SARS-CoV-2 virions, viral particles | Dark, in part rounded structures | Unidentifiable structure/s | insufficient | insufficient | no | NA | o | o | |
| 21 | Bulfamante et al. (heart) | Heart | 5E and F | 54 | SARS-CoV-2 virions, viral particles | Vesicular structure with empty lumen | Unidentifiable structure/s | insufficient | insufficient | no | NA | o | o | |
| 22 | Calabrese et al. [58] | Lung | 4 | 55 | Putative viral particles | Vesicular structures, some with dark granula | rER | insufficient* | sufficient | no | NA | o | o | Misleading description as putative viral particles (no morphologic features of coronavirus present), the suggested use of immunogold is also not reasonable in this example |
| 23 | Canini et al. [59] | BAL | 2G | 56 | Viral-like particles | Vesicular structures within membrane structure | Unidentifiable structure/s | sufficient* | sufficient | no | NA | o | o | |
| 24 | Carsana et al. [60] | Lung | 2 | 57 | Virions | Numerous vesicular structures | Unidentifiable structure/s | insufficient | insufficient | no | NA | x* | o | *) In Bullock et al. [61] described as rare viral particles |
| 25 | Cazzato et al. [62] | Skin | 4 | 58 | Virions | Coated vesicles, cytoplasm | CCV | sufficient* | sufficient | no | NA | o | o | |

| | | | | | | | | | | | | | | |
|---|---|---|---|---|---|---|---|---|---|---|---|---|---|---|
| 25 | Cazzato et al. | Skin | 8 | 59 | Viral particles | Vesicular structures in membrane compartments | Unidentifiable structure/s | sufficient* | sufficient | no | NA | o | o | |
| 25 | Cazzato et al. | Skin | 8 (inset) | 60 | Viral particles | Vesicular structures in membrane compartments | Unidentifiable structure/s | sufficient* | sufficient | no | NA | o | o | |
| 26 | Chaudhary et al. [63] | BAL | 4 - nr.1 with arrow/s | 61 | SARS-CoV-2, SARS-CoV-2 virus | Dark structures extracellularly | Unidentifiable structure/s | insufficient | insufficient | no | NA | o | o | |
| 26 | Chaudhary et al. | BAL | 4 - nr.2 with arrow/s | 62 | SARS-CoV-2, SARS-CoV-2 virus | Dark structure | Unidentifiable structure/s | insufficient | insufficient | no | NA | o | o | |
| 26 | Chaudhary et al. | BAL | 4 - nr.3 with arrow/s | 63 | SARS-CoV-2, SARS-CoV-2 virus | Dark and bright structures | Unidentifiable structure/s | insufficient | insufficient | no | NA | o | o | |
| 26 | Chaudhary et al. | BAL | 4 - nr.4 with arrow/s | 64 | SARS-CoV-2, SARS-CoV-2 virus | Dark structures, some rounded | Unidentifiable structure/s | insufficient | insufficient | no | NA | o | o | |
| 26 | Chaudhary et al. | BAL | 4 - nr.5 with arrow/s | 65 | SARS-CoV-2, SARS-CoV-2 virus | Two dark, round structures and a possible membrane structure | Unidentifiable structure/s | insufficient | insufficient | no | NA | o | o | |
| 26 | Chaudhary et al. | BAL | 4 - nr.6 with arrow/s | 66 | SARS-CoV-2, SARS-CoV-2 virus | Dark structure | Unidentifiable structure/s | insufficient | insufficient | no | NA | o | o | |
| 26 | Chaudhary et al. | BAL | 4 - nr.7+8 with arrow/s | 67 | SARS-CoV-2, SARS-CoV-2 virus | Dark structures | Unidentifiable structure/s | insufficient | insufficient | no* | NA | o | o | *) Some features of coronavirus due to adequate isomorphic appearance of multiple grouped structures within membrane compartments and electron density, but the limited resolution/image presentation does not allow for identification as virus as the other required features cannot be resolved (membrane, RNP, surface structure) |

| | | | | | | | | | | | | | |
|---|---|---|---|---|---|---|---|---|---|---|---|---|---|
| 26 | Chaudhary et al. | BAL | 4 - nr.9 with arrow/s | 68 | SARS-CoV-2, SARS-CoV-2 virus | Dark round structures in cytoplasm, on with empty lumen | Unidentifiable structure/s | insufficient | insufficient | no | NA | o | o |
| 26 | Chaudhary et al. | BAL | 4 - nr.10 with arrow/s | 69 | SARS-CoV-2, SARS-CoV-2 virus | Dark round structures | Unidentifiable structure/s | insufficient | insufficient | no | NA | o | o |
| 26 | Chaudhary et al. | BAL | 4 - nr.11 with arrow/s | 70 | SARS-CoV-2, SARS-CoV-2 virus | One amorphous and one possible membraneous structure | Unidentifiable structure/s | insufficient | insufficient | no | NA | o | o |
| 26 | Chaudhary et al. | BAL | 4 - nr.12 with arrow/s | 71 | SARS-CoV-2, SARS-CoV-2 virus | Membraneous structures | Unidentifiable structure/s | insufficient | insufficient | no | NA | o | o |
| 26 | Chaudhary et al. | BAL | 4 - nr.13 with arrow/s | 72 | SARS-CoV-2, SARS-CoV-2 virus | Amorphous dark structure | Unidentifiable structure/s | insufficient | insufficient | no | NA | o | o |
| 26 | Chaudhary et al. | BAL | 4 - nr.14 with arrow/s | 73 | SARS-CoV-2, SARS-CoV-2 virus | Rounded dark structure | Unidentifiable structure/s | insufficient | insufficient | no | NA | o | o |
| 27 | Chen et al. [64] | Lung | 3A | 74 | Viral particles | Coated vesicle, cytoplasm | CCV | sufficient | sufficient | no | NA | o | o |
| 27 | Chen et al. | Lung | 3B | 75 | Viral particles | Coated vesicle, cytoplasm | CCV | sufficient | sufficient | no | NA | o | o |
| 28 | Colmenero et al. [65] | Skin | 4D | 76 | Coronavirus-like particles | Coated vesicle | CCV | sufficient | sufficient | no | Baeck et al. [66] Colmenero et al. [67] (reply to Baeck et al.) Brealey and Miller [68] Colmenero et al. [69] (reply to Braeley and Miller) | x | x |
| 29 | Debelenko et al. [70] | Placenta | 2E | 77 | Consistent with virions, virion | Round dark structures, cytoplasm | Unidentifiable structure/s | insufficient | insufficient | no | NA | o | o |
| 30 | Deinhardt-Emmer et al. [71] | Lung | 2B | 78 | SARS-CoV-2 virus particles | Individual vesicular structure within membrane structure | Unidentifiable structure/s | sufficient | sufficient | no | NA | o | o |
| 30 | Deinhardt-Emmer et al. | Lung | 2C | 79 | SARS-CoV-2 virus particles | Individual vesicular structure within membrane structure | Unidentifiable structure/s | sufficient | sufficient | no | NA | o | o |

| | | | | | | | | | | | | | | |
|---|---|---|---|---|---|---|---|---|---|---|---|---|---|---|
| 30 | Deinhardt-Emmer et al. | Lung | 2D | 80 | SARS-CoV-2 virus particles | Individual vesicular structure within membrane structure | Unidentifiable structure/s | sufficient | sufficient | no | NA | o | o | |
| 30 | Deinhardt-Emmer et al. | Lung | 2E | 81 | SARS-CoV-2 virus particles | Individual vesicular structure within membrane structure | Unidentifiable structure/s | sufficient | sufficient | no | NA | o | o | |
| 31 | Deshmukh et al. [72] | Kidney | 2B | 82 | Viral-like particles | Coated vesicles | CCV | sufficient | insufficient | no | NA | o | o | |
| 31 | Deshmukh et al. | Kidney | 2C | 83 | Spiked projections protruding from spherical particles | Coated vesicles | CCV | sufficient | sufficient | no | NA | o | o | |
| 32 | Dolhnikoff et al. [73] | Heart | 3A | 84 | Viral particles | Dark particles | Unidentifiable structure/s | insufficient | insufficient | no | Dittmayer et al.* [5] Dolhnikoff et al. [74] (letter) Dittmayer et al. [6] (reply to Dolhnikoff et al.) | x | o | *) Other publications were also discussed |
| 32 | Dolhnikoff et al. | Heart | 3B | 85 | Viral particle | Clusters of dark particles, focally engulfed in membraneous, also partly granular structure | rER | insufficient | sufficient | no | see above | x | o | |
| 32 | Dolhnikoff et al. | Heart | 3C | 86 | Viral particles | Dark particles | Unidentifiable structure/s | insufficient | insufficient | no | see above | x | o | |
| 32 | Dolhnikoff et al. | Heart | 3D | 87 | Viral particle | Dark particles | Unidentifiable structure/s | insufficient | sufficient | no | see above | x | o | |
| 33 | Duarte-Neto et al. (multiorgan) [17] | Lung | 1G | 88 | Viral particles | Dark particles | Unidentifiable structure/s | insufficient | insufficient | no | NA | o | o | |
| 33 | Duarte-Neto et al. (multiorgan) | Heart | 1H | 89 | Virus particle | Dark, amorphous structure | Unidentifiable structure/s | insufficient | insufficient | no | NA | o | o | |
| 33 | Duarte-Neto et al. (multiorgan) | Heart | 2I | 90 | Viral particles | Dark, amorphous structures | Unidentifiable structure/s | insufficient | insufficient | no | NA | o | o | |
| 33 | Duarte-Neto et al. (multiorgan) | Lung | 3I and J | 91 | Viral particles | Dark particles | Unidentifiable structure/s | insufficient | insufficient | no | NA | o | o | |
| 33 | Duarte-Neto et al. (multiorgan) | Intestine | 4F | 92 | Coronavirus particles | Dark, amorphous structures | Unidentifiable structure/s | insufficient | insufficient | no | NA | o | o | |
| 33 | Duarte-Neto et al. (multiorgan) | Heart | 4I | 93 | Viral particles | Dark, amorphous structures | Unidentifiable structure/s | insufficient | insufficient | no | NA | o | o | |

| | | | | | | | | | | | | | |
|---|---|---|---|---|---|---|---|---|---|---|---|---|---|
| 33 | Duarte-Neto et al. (multiorgan) | Lung | 5D | 94 | Virus | Small clusters of dark, round particles (immunogold) on amorphous dark structure, no controls | Immunogold, unidentifiable structure/s | insufficient | insufficient | no | NA | o | o |
| 33 | Duarte-Neto et al. (multiorgan) | Heart | 5G | 95 | Viral particle | Round particle within membrane compartment | Unidentifiable structure/s | insufficient | insufficient | no | NA | o | o |
| 33 | Duarte-Neto et al. (multiorgan) | Heart | 5H | 96 | Virus particle | Small clusters of dark, round particles (immunogold) on amorphous dark structure, no controls | Immunogold, unidentifiable structure/s | insufficient | insufficient | no | NA | o | o |
| 33 | Duarte-Neto et al. (multiorgan) | Brain | 6C | 97 | Viral particles | Dark, amorphous structures | Unidentifiable structure/s | insufficient | insufficient | no | NA | o | o |
| 33 | Duarte-Neto et al. (multiorgan) | Brain | 6D | 98 | Virions | Small clusters of dark, round particles (immunogold) on amorphous dark structures, no controls | Immunogold, unidentifiable structure/s | insufficient | insufficient | no | NA | o | o |
| 33 | Duarte-Neto et al. (multiorgan) | Brain | 6E | 99 | Virus-like particles, viral particles | Vesicular structures with dark dots | Unidentifiable structure/s | insufficient | insufficient | no | NA | o | o |
| 33 | Duarte-Neto et al. (multiorgan) | Brain | 6F | 100 | Virus-like particles, virus | Vesicular structures, partly with dark dots | Unidentifiable structure/s | insufficient | insufficient | no | NA | o | o |
| 34 | Duarte-Neto et al. (testis) [18] | Testis | 3A | 101 | Viral particles | Dark vesicular structures | Unidentifiable structure/s | insufficient | insufficient | no | NA | o | o |
| 34 | Duarte-Neto et al. (testis) | Testis | 3B | 102 | Viral particles | Vesicular structures | Unidentifiable structure/s | insufficient | insufficient | no | NA | o | o |
| 34 | Duarte-Neto et al. (testis) | Testis | 3C | 103 | Virus particles | Vesicular structures | Unidentifiable structure/s | insufficient | insufficient | no | NA | o | o |
| 34 | Duarte-Neto et al. (testis) | Testis | 3D | 104 | Viral particles | Vesicular structures with faint granula | Unidentifiable structure/s | insufficient | insufficient | no | NA | o | o |
| 34 | Duarte-Neto et al. (testis) | Testis | 3E | 105 | Coronavirusparticles, coronavirus particle, SARS-CoV-2 particle | Vesicular structures, in part with surface projections | Unidentifiable structure/s | insufficient | insufficient | no | NA | o | o |

| | | | | | | | | | | | | | |
|---|---|---|---|---|---|---|---|---|---|---|---|---|---|
| 34 | Duarte-Neto et al. (testis) | Testis | 3F | 106 | Coronavirusparticles, virallike particle | Dark amorphous structure | Unidentifiable structure/s | insufficient | insufficient | no | NA | o | o | |
| 34 | Duarte-Neto et al. (testis) | Testis | 3G | 107 | Viral particles, virus particle | Vesicular structures with prominent granula, within membrane compartment that shows attached granula of similar size | rER | insufficient | insufficient | no | NA | o | o | |
| 35 | Erman et al. [75] | Nasopharynx | 2B to D | 108 | SARS-CoV-2 virions | Electron dense DAB-product on multiple dark, vesicular structures | DAB, unidentifiable structure/s | insufficient | insufficient | no | NA | o | o | DAB-preembedding using punch biopsy of FFPE block |
| 35 | Erman et al. | Nasopharynx | 3A | 109 | Presumed virions | Multiple vesicular structures, cytoplasm | CCV | insufficient* | insufficient | no | NA | o | o | Nonspecific stain is demonstrated by the authors (before antigen retrival) |
| 35 | Erman et al. | Nasopharynx | 3B | 110 | Presumed virions | Multiple vesicular structures, some with surface structures, cytoplasm | CCV | insufficient* | insufficient | no | NA | o | o | Nonspecific stain is demonstrated by the authors (before antigen retrival) |
| 35 | Erman et al. | Nasopharynx | 3C | 111 | Virions | Multiple vesicular structures in membrane compartment, one with goldparticle on profile | Immunogold, unidentifiable structure/s | insufficient* | sufficient | no | NA | o | o | Specific stain is demonstrated by the authors (after antigen retrival) |
| 35 | Erman et al. | Nasopharynx | 3D | 112 | SARS-CoV-2 virion | Individual structure with multiple goldparticles on profile | Immunogold, unidentifiable structure/s | insufficient* | insufficient | no | NA | o | o | Specific stain is demonstrated by the authors (using Lowicryl section) |
| 36 | Evert et al. [76] | Lung | 2A and B | 113 | At best, "virus-like" particles, structures that may represent autolytic virus particles were identified (…) | Round structure within membrane compartment | Unidentifiable structure/s | insufficient* | sufficient | no | NA | o | o | Critically discussed by the authors, but no morphological features of coronavirus particles |

| | | | | | | | | | | | | | |
|---|---|---|---|---|---|---|---|---|---|---|---|---|---|
| 36 | Evert et al. | Lung | 2C and D | 114 | At best, "virus-like" particles, structures that may represent autolytic virus particles were identified (…) | Oval structure with irregular outlines | Unidentifiable structure/s | insufficient* | sufficient | no | NA | o | o | Critically discussed by the authors, but no morphological features of coronavirus particles |
| 37 | Facchetti et al. [77] | Placent a | 2B | 115 | Free particle, (…) morphological features consistent with coronavirus | Vesicular structure | Unidentifiable structure/s | insufficient | insufficient | no | NA | o | x | |
| 37 | Facchetti et al. | Placent a | C2 | 116 | Particle consistent with mature coronavirus | Vesicular structure | Unidentifiable structure/s | insufficient | insufficient | no | NA | o | x | |
| 37 | Facchetti et al. | Placent a | C4 | 117 | Particle with the typical morphological features of coronavirus | Vesicular structure with surface projections | Unidentifiable structure/s | insufficient | insufficient | no | NA | o | x | |
| 37 | Facchetti et al. | Placent a | D1 | 118 | Particles morphologically consistent with SARV-CoV-2 | Vesicular structures | Unidentifiable structure/s | insufficient | insufficient | no | NA | o | x | |
| 37 | Facchetti et al. | Placent a | 3A and B | 119 | SARS-CoV-2 viral particle, particle morphologically consistent with coronavirus | Round particle with prominent surface structures | Unidentifiable structure/s | insufficient | insufficient | no | NA | o | x | |
| 38 | Falasca et al. [78] | Lung | 3B | 120 | Viral particles | Vesicular structures (singly and multiple) in membrane compartments of different sizes | Unidentifiable structure/s | sufficient | insufficient | no | NA | x* | o | *) In Bullock et al. [61] described as rare viral particles |
| 38 | Falasca et al. | Lung | 3C | 121 | Viral particles | Vesicular structures (singly and multiple) in membrane compartments of different sizes | Unidentifiable structure/s | sufficient | insufficient | no | NA | x* | o | |
| 38 | Falasca et al. | Lung | 3D | 122 | Viral particles | Vesicular structures (singly and multiple) in membrane compartments of different sizes, some particles show electron dense granula | Unidentifiable structure/s | sufficient | insufficient | no | NA | x* | o | |

| | | | | | | | | | | | | | | |
|---|---|---|---|---|---|---|---|---|---|---|---|---|---|---|
| 39 | Farkash et al. [79] | Kidney | 3B | 123 | Viral arrays | Clusters of dark vesicular structures | Unidentifiable structure/s | insufficient | insufficient | no | Miller and Goldsmith [80] Delsante et al. [81] Farkash et al. [82] (reply to Miller and Goldsmith and Delsante et al.) Roufosse et al.* [83] | x | x | *) Other publications were also discussed |
| 39 | Farkash et al. | Kidney | 3C | 124 | Viruses, virus | Clusters of vesicles, some with surface projections | Unidentifiable structure/s | insufficient | insufficient | no | see above | x | x | |
| 40 | Fiel et al. [84] | Liver | 2A | 125 | Viral-like particles, virions | Multiple vesicular structures | Unidentifiable structure/s | insufficient | insufficient | no | NA | o | o | |
| 40 | Fiel et al. | Liver | 2B | 126 | Viral-like particles, virions | Multiple vesicular structures | Unidentifiable structure/s | insufficient | insufficient | no | NA | o | o | |
| 41 | Fox et al. (Circulation) [85] | Heart | C | 127 | Particles consistent with SARS-CoV-2 virus | Vesicular structures | Unidentifiable structure/s | sufficient | insufficient | no | NA | o | o | |
| 41 | Fox et al. (Circulation) | Lung | G | 128 | Particles consistent with SARS-CoV-2 virus | Multiple vesicular structures with relatively homogeneous appearing interior | Unidentifiable structure/s | sufficient | insufficient | no | NA | o | o | |
| 41 | Fox et al. (Circulation) | Kidney | H | 129 | Particles consistent with SARS-CoV-2 virus | Clusters of vesicles, some with surface projections | Unidentifiable structure/s | sufficient | insufficient | no | NA | o | o | |
| 42 | Fox et al. (Lancet Respir Med) [86] | Lung | 4E | 130 | Particles suggestive of viral infection | Coated vesicles, cytoplasm | CCV | sufficient | sufficient | no | NA | o | x | |
| 43 | Garau et al. [87] | Heart | 4A and B | 131 | Viral particles, (…) suggests coronavirus particles | Coated vesicle, cytoplasm | CCV | sufficient* | sufficient | no | NA | o | o | |
| 44 | García-Cruz et al. [88] | Pericardium | E | 132 | Viral particles | Rounded structures | Unidentifiable structure/s | insufficient | insufficient | no | NA | o | o | |
| 44 | García-Cruz et al. | Pericardium | F | 133 | Viral particles | Rounded structures | Unidentifiable structure/s | insufficient | insufficient | no | NA | o | o | |
| 45 | Garrido Ruiz et al. [89] | Skin | 2C | 134 | Viral inclusion particles | Coated vesicles, cytoplasm | CCV | sufficient | insufficient | no | NA | o | o | |
| 45 | Garrido Ruiz et al. | Skin | 3C | 135 | Viral inclusions | Coated vesicle, cytoplasm | CCV | sufficient | insufficient | no | NA | o | o | |
| 45 | Garrido Ruiz et al. | Skin | 5B | 136 | Viral inclusions | Vesicular structure | Unidentifiable structure/s | insufficient | insufficient | no | NA | o | o | |
| 46 | Gioia et al. [90] | Placenta | 4.1 and 4.2 | 137 | Virus particles | Vesicular structures with surface | CCV | sufficient | sufficient | no | NA | o | o | |

| | | | | | | | | | | | | | |
|---|---|---|---|---|---|---|---|---|---|---|---|---|---|
| | | | | | | projections, cytoplasm | | | | | | | |
| 46 | Gioia et al. | Placent a | 4.3 | 138 | Virus particles | Vesicular structures with surface projections, cytoplasm | CCV | sufficient | sufficient | no | NA | o | o |
| 47 | Grimes et al. [91] | Lung | 2A | 139 | Viral particles, viral-like particles | Numerous vesicular structures of different electron density within membrane compartment | Unidentifiable structure/s | insufficient | insufficient | no | NA | x | o |
| 47 | Grimes et al. | Lung | 2B | 140 | Typical morphology of Coronavirus (SARS-CoV-2) | Oval-shaped vesicular structure within membrane compartment | Unidentifiable structure/s | insufficient | insufficient | no | NA | x | o |
| 48 | Hakroush et al. [92] | Lung | 1I | 141 | Virus-like particles (...) fulfilling the criteria of size (diameter: 100 nm), shape and structural features (membrane, surface structures, electron dense material within the particle resembling ribonucleoprotein and cytoplasmic localisation within a membrane compartment, partial attachment to the inner membrane surface) | Vesicular structures within membrane structure, empty appearing interior | Unidentifiable structure/s | insufficient | insufficient | no | NA | o | o |
| 49 | Han et al. [93] | Colon | 2C | 142 | SARS-CoV-2 viral particles | Multiple vesicular structures | Unidentifiable structure/s | insufficient | insufficient | no | NA | o | o |
| 50 | Hooper et al. [94] | Skeleta l muscle | 1C | 143 | Virus-like particles | Vesicular structures | Unidentifiable structure/s | insufficient | insufficient | no | NA | o | o |
| 50 | Hooper et al. | Skeleta l muscle | 1D | 144 | Virus-like particles | Vesicular structures | Unidentifiable structure/s | insufficient | insufficient | no | NA | o | o |
| 50 | Hooper et al. | Skeleta l muscle | 1E | 145 | Virus-like particles | Coated vesicles | Unidentifiable structure/s | insufficient | insufficient | no | NA | o | o |
| 50 | Hooper et al. | Skeleta l muscle | 1E (inset left) | 146 | Virus-like particles | Coated vesicle | Unidentifiable structure/s | insufficient | insufficient | no | NA | o | o |
| 50 | Hooper et al. | Skeleta l muscle | 1E (inset right) | 147 | Virus-like particles | Coated vesicle | CCV | insufficient | sufficient | no | NA | o | o |

| | | | | | | | | | | | | | |
|---|---|---|---|---|---|---|---|---|---|---|---|---|---|
| 51 | Hosier et al. [95] | Placenta | 4C | 148 | Virus particles | Vesicular structures | Unidentifiable structure/s | sufficient | insufficient | no | NA | x | x | |
| 51 | Hosier et al. | Placenta | 4D | 149 | Virus particles | Vesicular structures | Unidentifiable structure/s | sufficient | insufficient | no | NA | x | x | |
| 51 | Hosier et al. | Placenta | 4E | 150 | Virus particles | Coated vesicle | Unidentifiable structure/s | sufficient | insufficient | no | NA | x | x | |
| 51 | Hosier et al. | Placenta | 4F | 151 | Virus particles | Vesicular structure in membrane compartment | Unidentifiable structure/s | sufficient | sufficient | no | NA | x | x | |
| 51 | Hosier et al. | Placenta | 4G | 152 | Virus particles | Vesicular structures | Unidentifiable structure/s | sufficient | insufficient | no | NA | x | x | |
| 51 | Hosier et al. | Placenta | 4G (inset) | 153 | Virus particles | Coated vesicle | Unidentifiable structure/s | sufficient | sufficient | no | NA | x | x | |
| 51 | Hosier et al. | Placenta | 4H | 154 | Virus particles | Vesicular structures with double-membrane aspect | Unidentifiable structure/s | sufficient | sufficient | no | NA | x | x | |
| 51 | Hosier et al. | Placenta | 4I | 155 | Viral particles | Vesicular structure with surface projections, membrane invaginations | Unidentifiable structure/s | sufficient | sufficient | no | NA | x | x | Perhaps two caveolae (top), others unidentifiable |
| 52 | Idrissi et al. [96] | Cornea | A1 to A3 | 156 | Virus-like particles, SARS-CoV-2-like particles | Round dark structures within membrane structure | Unidentifiable structure/s | insufficient | insufficient | no | NA | o | o | |
| 52 | Idrissi et al. | Cornea | B1 to B3 | 157 | Virus-like particles, SARS-CoV-2-like particles | Round dark structures within membrane structure | Unidentifiable structure/s | insufficient | insufficient | no | NA | o | o | |
| 53 | Ingravallo et al. [97] | Skin | 4 | 158 | Maturing coronaviruses, whole virions | Large, dark, vesicular structures with surface projections | Unidentifiable structure/s | insufficient* | sufficient | no | NA | o | o | Information of scale bar and size-dimensions of putative coronaviurs particles do not match; indicated in the text with 65 to 136 nm, but the large structure in Fig. 4 would be around 200 nm (scale bar indiacated with 500 nm) |
| 54 | Jacobs et al. [98] | Platelets | 3A and B | 159 | Presumptive SARS-CoV-2 virion | Vesicular structure in membrane compartment | Unidentifiable structure/s | insufficient | insufficient | no | NA | o | o | |
| 54 | Jacobs et al. | Platelets | 3C and D | 160 | Presumptive SARS-CoV-2 virion | Vesicular structure with heterogen granular interior and surface structures in membrane compartment | Unidentifiable structure/s | insufficient | sufficient | no | NA | o | o | |
| 54 | Jacobs et al. | Platelets | 3E | 161 | Presumptive SARS-CoV-2 virions | Vesicular structures | Unidentifiable structure/s | insufficient | insufficient | no | NA | o | o | |

| | | | | | | | | | | | | | | |
|---|---|---|---|---|---|---|---|---|---|---|---|---|---|---|
| 54 | Jacobs et al. | Platelets | 3F | 162 | Presumptive virions | Vesicular structures | Unidentifiable structure/s | insufficient | insufficient | no | NA | o | o | |
| 55 | Jeican et al. (ear) [99] | Middle ear mucosa | 2B | 163 | Structures suggestive of SARS-CoV-2 virus | Vesicular structures with empty appearing interior | Unidentifiable structure/s | insufficient | insufficient | no | NA | o | o | Perhaps caveolae |
| 55 | Jeican et al. (ear) | Middle ear mucosa | 2C | 164 | Structures suggestive of SARS-CoV-2 virus | Vesicular structure with realtively homogeneously appearing interior | Unidentifiable structure/s | insufficient | insufficient | no | NA | o | o | Perhaps cross section through cell process/protrusion |
| 56 | Jeican et al. (nose) [100] | Nasal mucosa | 8A and B | 165 | Structures suggestive of the SARS-CoV-2 virus | Vesicular structures with heterogeneous substructure of relative low electron density | Cell processes | sufficient | insufficient | no | NA | o | o | |
| 57 | Kadosh et al. [101] | Kidney | 2 (top left) | 166 | Coronavirus particles | Coated vesicle, cytoplasm | CCV | sufficient | sufficient | no | NA | o | o | |
| 57 | Kadosh et al. | Kidney | 2 (top right) | 167 | Coronavirus particles | Coated vesicle, cytoplasm | CCV | sufficient | sufficient | no | NA | o | o | |
| 57 | Kadosh et al. | Kidney | 2( bottom ) | 168 | Coronavirus particles | Coated vesicle, cytoplasm | CCV | sufficient | sufficient | no | NA | o | o | |
| 58 | Kanczkowski et al. [102] | Adrenal gland | C | 169 | Viral-like particles, SARS-CoV-2 virus-like particles | Multiple vesicular structures with empty appearing interior | Unidentifiable structure/s | insufficient | insufficient | no | NA | o | o | |
| 59 | Khismatullin et al. [103] | Lung | 3A | 170 | Coronavirus-like particles | Vesicular, partly dark structures in membrane comparment | Unidentifiable structure/s | insufficient | insufficient | no | NA | o | o | |
| 59 | Khismatullin et al. | Lung | 3B | 171 | Virion-like particles | Multiple dark particles | Unidentifiable structure/s | insufficient | insufficient | no | NA | o | o | |
| 59 | Khismatullin et al. | Lung | 3C | 172 | Extracellular particle with distinctive spike-like projections | Structure with dark peripheral granules and heterogeneous dark interior | Unidentifiable structure/s | insufficient | insufficient | no | NA | o | o | |
| 60 | Kissling et al. [104] | Kidney | 1E and F | 173 | Numerous spherical particles (…) may correspond to viral inclusion bodies reported (…) | Vesicular structures within membrane compartment | MVB | sufficient | insufficient | no | Calomeni et al. [105]<br>Miller and Brealey* [106]<br>Roufosse et al.* [83]<br>Kissling et al. [107] (reply to Miller and Brealy) | x | x | *) Other publication(s) were also discussed |
| 61 | Kolivras et al. [108] | Skin | 6A | 174 | Probable viral particle | Coated vesicles, cytoplasm | CCV | insufficient | insufficient | no | NA | o | o | |
| 61 | Kolivras et al. | Skin | 6B | 175 | Probable viral particles | Vesicular structures with dark particles and empty appearing interior | Unidentifiable structure/s | insufficient | insufficient | no | NA | o | o | |

| | | | | | | | | | | | | | | |
|---|---|---|---|---|---|---|---|---|---|---|---|---|---|---|
| 61 | Kolivras et al. | Skin | 6C | 176 | Probable viral particle | Coated vesicle, cytoplasm | CCV | insufficient | insufficient | no | NA | o | o | |
| 61 | Kolivras et al. | Skin | 6D | 177 | Probable viral particle | Vesicular structure with empty appearing interior | Unidentifiable structure/s | insufficient | insufficient | no | NA | o | o | |
| 61 | Kolivras et al. | Skin | 6E | 178 | Probable viral particle | Vesicular structure with empty appearing interior | Unidentifiable structure/s | insufficient | insufficient | no | NA | o | o | |
| 61 | Kolivras et al. | Skin | 6F | 179 | Probable viral particle | Coated vesicles, cytoplasm | CCV | insufficient | insufficient | no | NA | o | o | |
| 62 | Kresch et al. [109] | Penis | 1A | 180 | Coronavirus-like spiked viral particles | Vesicular structure with dark granula attached to membrane and relatively homogeneous interior | rER | sufficient | sufficient | no | NA | o | o | |
| 62 | Kresch et al. | Penis | 1B | 181 | Coronavirus-like spiked viral particles | Vesicular structure, cytoplasm | Unidentifiable structure/s | sufficient | insufficient | no | NA | o | o | |
| 63 | Lauermann et al. [110] | Cornea | 1D | 182 | Virus-like particles | Vesicular, round to oval shaped structures within membrane compartment | Mitochondrion /ia | sufficient | insufficient | no | NA | o | o | |
| 63 | Lauermann et al. | Cornea | 1E | 183 | Virus-like particles | Vesicular, round to oval shaped structures within membrane compartment | Unidentifiable structure/s | sufficient | insufficient | no | NA | o | o | |
| 64 | Lei et al. [111] | Glioblastoma | 2 (middle and right image) | 184 | SARS-CoV-2, coronavirus | Coated vesicle, cytoplasm | CCV | sufficient | sufficient | no | NA | o | o | |
| 65 | Leoni et al. [112] | Skin | 2B | 185 | Viral particles with the morphological features of corona virus particles | Multiple vesicles within membrane compartment | MVB | sufficient | insufficient | no | NA | o | o | |
| 65 | Leoni et al. | Skin | 2C (top) | 186 | Virus | Vesicle in membrane compartment | Unidentifiable structure/s | sufficient | insufficient | no | NA | o | o | |
| 66 | Li et al. [113] | Lung | 3B | 187 | Viral particles | Multiple vesicular structures within membrane compartments | MVB | insufficient | insufficient | no | NA | o | o | Structure at top perhaps severely autolytic mitochondrium |
| 66 | Li et al. | Lung | 4C | 188 | Viral particles | Vesicular structures, cytoplasm | Unidentifiable structure/s | insufficient | insufficient | no | NA | o | o | |

| | | | | | | | | | | | | | | |
|---|---|---|---|---|---|---|---|---|---|---|---|---|---|---|
| 67 | Lin et al. [114] | Kidney | 1A (top and bottom ) | 189 | Spherical particles measuring around 80 nm (arrow) and surrounded by spikes, spherical particles surrounded by crown-like projections, which were highly suspected as SARS-CoV-2 virus | Coated vesicle, cytoplasm | CCV | sufficient* | insufficient | no | NA | o | o | |
| 68 | Liu et al. [115] | Skin | 1E | 190 | Viral particles | Vesicular structures showing a partly granular interior, located within membrane compartment | Unidentifiable structure/s | insufficient* | insufficient | no | NA | o | o | |
| 69 | Livanos et al. [116] | Intestin e | 2R, S and V | 191 | Presumptive virion | Dark particle in putative membrane compartment | Unidentifiable structure/s | sufficient | insufficient | no | NA | o | o | |
| 69 | Livanos et al. | Intestin e | 2T, U and W | 192 | Presumptive SARS-CoV-2 virions, presumptive virion, virion | Multiple vesicular structures within membrane compartment, W shows a vesicle with coated surface | MVB | sufficient | sufficient | no | NA | o | o | |
| 69 | Livanos et al. | Intestin e | 2X | 193 | Presumptive SARS-CoV-2 virion | Dark particles (immunogold) on granular background | Immunogold, unidentifiable structure/s | insufficient* | insufficient | no | NA | o | o | |
| 69 | Livanos et al. | Intestin e | 2Y | 194 | Presumptive SARS-CoV-2 virion | Dark particles (immunogold) and vesicular structure with homogeneous lumen | Immunogold, unidentifiable structure/s | insufficient* | insufficient | no | NA | o | o | |
| 70 | Lubnow et al. [117] | Liver | S1E and F | 195 | Virus-like particles | Coated vesicle, cytoplasm | CCV | sufficient | sufficient | no | NA | o | o | |
| 71 | Ma et al. [118] | Testis | 1G (overvi ew to detail image s) | 196 | Coronavirus-like particles | Dark particle with surface structures | Unidentifiable structure/s | insufficient | insufficient | no | NA | o | o | |
| 71 | Ma et al. | Testis | S2C (overvi ew to detail image s) | 197 | Coronavirus-like particles | Vesicular structures | Unidentifiable structure/s | insufficient | insufficient | no | NA | o | o | |
| 72 | Martin-Cardona et al. [119] | Colon | 2E and F | 198 | Viral particles | Dark vesicular structure with relatively | Unidentifiable structure/s | insufficient | sufficient | no | NA | o | o | Perhaps CCV |

| | | | | | | | | | | | | | | |
|---|---|---|---|---|---|---|---|---|---|---|---|---|---|---|
| | | | | | | homogeneous dark interior | | | | | | | | |
| 72 | Martin-Cardona et al. | Lymphoma | 3E | 199 | Viral particles | Particle, cytoplasm | Unidentifiable structure/s | sufficient | insufficient | no | NA | o | o | |
| 72 | Martin-Cardona et al. | Lymphoma | 3F | 200 | Coronavirus particles | Vesicular structure with empty interior and surface structures, cytoplasm | CCV | sufficient | sufficient | no | NA | o | o | |
| 72 | Martin-Cardona et al. | Colon | 5E | 201 | Virus particles | Oval structure with empty interior, cytoplasm | Unidentifiable structure/s | insufficient | insufficient | no | NA | o | o | |
| 73 | Matuck et al. [120] | Parotid gland | 1B | 202 | Viral particles | Vesicular structures, extracellularly | Unidentifiable structure/s | sufficient | insufficient | no | NA | o | o | |
| 73 | Matuck et al. | Submandibular gland | 1C | 203 | Viral particles | Vesicular structures, intracellularly | Unidentifiable structure/s | sufficient | insufficient | no | NA | o | o | |
| 74 | McMullen et al. [25] | Lung | 3A | 204 | Candidate virion-like particles | Membrane bound dark particles | rER | insufficient | insufficient | no | NA | o | o | |
| 75 | Menter et al. [121] | Kidney | 4A and B | 205 | Possible virus-like particles, virus-like particles | Multiple vesicular structures within membrane compartments | MVB | sufficient | sufficient | no | NA | x | x | |
| 75 | Menter et al. | Kidney | 4C | 206 | Virus-like particles | Multiple vesicular structures within membrane compartments | MVB | sufficient | insufficient | no | NA | x | x | |
| 75 | Menter et al. | Kidney | 4D | 207 | Virus-like particles | Membrane bound dark particles | rER | sufficient | insufficient | no | NA | x | x | |
| 76 | Mitchell et al. [122] | Intestine | 1D | 208 | Viral particles | Vesicular dark, partly granular strucures within membrane compartment | Unidentifiable structure/s | sufficient* | sufficient | no | NA | o | o | |
| 77 | Morbini et al. [123] | Olfactoy epithelium | 1A | 209 | Viral particles | Round structure with prominent surface projections | Unidentifiable structure/s or virus particle | sufficient | sufficient | no* | NA | o | o | *) But suspicious |
| 77 | Morbini et al. | Olfactory bulb | 1B | 210 | Viral cytoplasmic inclusion body | Round structures within membrane compartment | Unidentifiable structure/s | sufficient | insufficient | no | NA | o | o | |
| 77 | Morbini et al. | Olfactory bulb | 1C | 211 | Viral particles | Vesicular structures between interstitial fibres | Unidentifiable structure/s | sufficient | insufficient | no | NA | o | o | |
| 78 | Nardacci et al. [124] | Lung | 6B | 212 | Viral particles | Vesicular structures, partly granular interior, within membrane compartment | MVB | sufficient | insufficient | no | NA | o | o | |

| | | | | | | | | | | | | | | |
|---|---|---|---|---|---|---|---|---|---|---|---|---|---|---|
| 78 | Nardacci et al. | Lung | 6C | 213 | Viral particles | Dark, vesicular structure with partly granular interior within membrane compartment | Unidentifiable structure/s | sufficient* | insufficient | no | NA | o | o | |
| 78 | Nardacci et al. | Lung | 6D | 214 | Virus-containing compartments | Vesicular structures in membrane compartments | Unidentifiable structure/s | insufficient* | insufficient | no | NA | o | o | |
| 79 | Paniz-Mondolfi et al. [125] | Brain | 1A | 215 | Virus particles | Vesicular structures in membrane compartment | Unidentifiable structure/s | insufficient | insufficient | no | NA | x | o | |
| 79 | Paniz-Mondolfi et al. | Brain | 1B | 216 | Viral particles | Vesicular, heterogeneous structure | Unidentifiable structure/s | insufficient | insufficient | no | NA | x | o | |
| 79 | Paniz-Mondolfi et al. | Brain | 1C and D (inset) | 217 | Viral particles, viral-like particles | Multiple vesicular structures within membrane compartment | Unidentifiable structure/s | insufficient | insufficient | no | NA | x | o | |
| 80 | Parrón et al. [126] | Testis | 3 | 218 | Spiked viral particles | Coated vesicles, cytoplasm | CCV | insufficient* | insufficient | no | NA | o | o | |
| 81 | Pérez et al. [127] | Kidney | 3B and C | 219 | Viral particles | Coated vesicle, cytoplasm | CCV | sufficient* | sufficient | no | NA | o | o | |
| 82 | Pesaresi et al. [128] | Lung | 1B | 220 | Virions | Groups of multiple vesicular structures, partly engulfed by lamellar structure | Unidentifiable structure/s | sufficient* | insufficient | no | NA | x | x | |
| 82 | Pesaresi et al. | Lung | 1C | 221 | Viral particles | Vesicular structure with attached granula (right) | rER | sufficient* | insufficient | no | NA | x | x | Glycogen (left) |
| 82 | Pesaresi et al. | Lung | 1D | 222 | Viral particles | Membrane cisternae with attached granula | rER | sufficient* | insufficient | no | NA | x | x | |
| 82 | Pesaresi et al. | Heart | 2A and B | 223 | Viral particles | Dark structures | Unidentifiable structure/s | insufficient | Insufficient | no | NA | x | x | |
| 82 | Pesaresi et al. | Kidney | 2E | 224 | Viral particles | Dark granula | Unidentifiable structure/s | insufficient | insufficient | no | NA | x | x | |
| 83 | Pirisi et al. [129] | Liver | 1B | 225 | Virus-like particles | Vesicular structure in perinuclear cisterna | Unidentifiable structure/s | sufficient | insufficient | no | NA | o | o | |
| 83 | Pirisi et al. | Liver | 1C | 226 | Virions | Vesicular structures | Unidentifiable structure/s | sufficient | insufficient | no | NA | o | o | |
| 83 | Pirisi et al. | Liver | 1D | 227 | Budding | Membrane protrusion within membrane compartment | Unidentifiable structure/s | sufficient | insufficient | no | NA | o | o | |

| | | | | | | | | | | | | | |
|---|---|---|---|---|---|---|---|---|---|---|---|---|---|
| 83 | Pirisi et al. | Liver | 1E and F | 228 | Virus-like particle | Dark structure in membrane compartment | Unidentifiable structure/s | sufficient | insufficient | no | NA | o | o | |
| 84 | Prieto-Pérez et al. [130] | Lung | 5B | 229 | Particles of size and morphology consistent with coronavirus | Vesicular structures within membrane compartments | Unidentifiable structure/s | insufficient | insufficient | no | NA | o | o | |
| 84 | Prieto-Pérez et al. | Lung | 5C | 230 | Coronavirus particles | Vesicular structures in membrane compartment | MVB | insufficient | insufficient | no | NA | o | o | |
| 85 | Qadir et al. [131] | Pancreas | 6A (top) | 231 | SARS-CoV-2 particles | Multiple vesicular structures in membrane compartment, some with partly dark interior | Unidentifiable structure/s | sufficient* | insufficient | no | NA | o | o | |
| 85 | Qadir et al. | Pancreas | 6A (bottom) | 232 | SARS-CoV-2 particles | Multiple vesicular structures in membrane comparment, some with partly granular dark interior, some also with surface structures | Unidentifiable structure/s | sufficient* | insufficient | no* | NA | o | o | *) Some features of coronavirus particles, but insufficient quality for adequate evaluation |
| 85 | Qadir et al. | Pancreas | 6B | 233 | SARS-CoV-2 particles | Multiple vesicular structure in membrane compartment | MVB | sufficient* | insufficient | no | NA | o | o | |
| 86 | Qian et al. [132] | Intestine | 3A | 234 | SARS-CoV-2 Virions, viral particles | Vesicular structures with empty appearing interior | Unidentifiable structure/s | insufficient | insufficient | no | NA | x | x | |
| 86 | Qian et al. | Intestine | 3B | 235 | SARS-CoV-2 Virions, viral particles | Vesicular particle with empty appearing interior and surface structures | Unidentifiable structure/s | insufficient | insufficient | no | NA | x | x | |
| 87 | Rassaf et al. [133] | Heart | D (top and middle image) | 236 | SARS-CoV-2, structures with high similarities to SARS virus | Coated vesicle, cytoplasm | CCV | sufficient | sufficient | no | NA | o | o | |
| 87 | Rassaf et al. | Heart | D (bottom image) | 237 | SARS-CoV-2, structures with high similarities to SARS virus | Coated vesicle, cytoplasm | CCV | sufficient | sufficient | no | NA | o | o | |
| 87 | Rassaf et al. | Heart | E | 238 | Representative particle, structures with high similarities to SARS virus | Coated vesicle, cytoplasm | CCV | sufficient | sufficient | no | NA | o | o | |
| 88 | Ren et al. [134] | Lung | 3H | 239 | Single spherical viral particles | Vesicular structures | Unidentifiable structure/s | insufficient* | insufficient | no | NA | o | o | |

| | | | | | | | | | | | | | | |
|---|---|---|---|---|---|---|---|---|---|---|---|---|---|---|
| 89 | Resta et al. (article) [135] | Placenta | 10 and 11 | 240 | Virion; likely viral particles | Coated vesicles, cytoplasm | CCV | sufficient | sufficient | no | NA | o | o | |
| 90 | Resta et al. (case report) [135] | Placenta | 3 | 241 | Likely viral particles | Round vesicular structure with empty interior | Unidentifiable structure/s | insufficient | sufficient | no | NA | o | o | |
| 91 | Santana et al. [136] | Lung | 5A to C | 242 | Viral particles | Vesicular structures with faint surface projections and focally dark luminal components (particle at bottom) | Unidentifiable structure/s | sufficient* | insufficient | no** | NA | o | o | **) One structure with some features of coronavirus particles, but not sufficient |
| 91 | Santana et al. | Lung | 5D and E | 243 | Viral particles | Vesicular structures in membrane compartment | Unidentifiable structure/s | sufficient* | insufficient | no | NA | o | o | Perhaps MVB |
| 91 | Santana et al. | Lung | 5D and F | 244 | Viral particles | Vesicular, irregular structures | Unidentifiable structure/s | sufficient* | insufficient | no | NA | o | o | |
| 92 | Schoenmakers et al. [137] | Placenta | 1D | 245 | SARS-CoV-2 particles | Rounded structure | Unidentifiable structure/s | insufficient* | insufficient | no | NA | o | o | |
| 93 | Schwab et al. [138] | Colon | 1D (C in legend ) | 246 | Viral particles | Multiple grouped dark vesicular particles | Unidentifiable structure/s | insufficient | insufficient | no* | NA | o | o | *) Suggestive of coronavirus particles in grouping, isomorphic appearance and electron density, but insufficient preservation/image quality to proof presence of CoV particles |
| 94 | Sisman et al. [139] | Placenta | 1 | 247 | Viral-like particles | Vesicular structures with relatively homogeneous interior in membrane compartment | Unidentifiable structure/s | sufficient | sufficient | no | NA | x | x | |
| 95 | Stahl et al. [140] | Colon | 1B | 248 | Coronaviruses, virion particles | Coated vesicles, cytoplasm, and vesicles within dilated cisterna of rER | CCV | sufficient | sufficient | no | Stahl et al. [141] | o | o | |
| 96 | Steenblock et al. [142] | Pancreas | 1D | 249 | Virus-like particles | Vesicular structures | Unidentifiable structure/s | insufficient | insufficient | no | NA | o | o | |
| 97 | Su et al. [143] | Kidney | 2A | 250 | Virus particles | Group of vesicular particles with surface structures, cytoplasm | CCV | sufficient* | sufficient | no | Calomeni et al. [105] ** Miller and Brealey** [106] Roufosse et al. ** [83] Su et al. [144] (reply to Miller and Brealey) | x | x | **) Other publication(s) were also discussed |
| 97 | Su et al. | Kidney | 2B | 251 | Virus particles | Group of vesicular particles with | CCV | sufficient* | sufficient | no | see above | x | x | |

| | | | | | | | | | | | | | | |
|---|---|---|---|---|---|---|---|---|---|---|---|---|---|---|
| | | | | | | surface structures, cytoplasm | | | | | | | | |
| 97 | Su et al. | Kidney | 2C | 252 | Virus particles | Vesicular structures | CCV | sufficient* | sufficient | no | see above | x | x | |
| 97 | Su et al. | Kidney | 2D | 253 | Virus particles | Vesicular particle with empty appearing interior, showing surface structures, and attaching on membrane, cytoplasm | CCV | sufficient* | sufficient | no | see above | x | x | |
| 98 | Tavazzi et al. [145] | Heart | 2A | 254 | Viral particles | Vesicular structures with surface projections, cytoplasm | CCV | insufficient | sufficient | no | Miller and Goldsmith* [80] Dittmayer et al.* [5] | x | x | *) Other publications were also discussed |
| 98 | Tavazzi et al. | Heart | 2B and C | 255 | Viral particles | Vesicular structures with surface projections, cytoplasm | CCV | insufficient | sufficient | no | see above | x | x | |
| 98 | Tavazzi et al. | Heart | 2D | 256 | Viral particles | Vesicular structure with surface projections, cytoplasm | CCV | insufficient | sufficient | no | see above | x | x | |
| 98 | Tavazzi et al. | Heart | 2E | 257 | Viral particles | Vesicular structures with surface projections, cytoplasm | CCV | insufficient | sufficient | no | see above | x | x | |
| 98 | Tavazzi et al. | Heart | 2F | 258 | Viral particles | Vesicular structure with surface projections, cytoplasm | CCV | insufficient | sufficient | no | see above | x | x | |
| 98 | Tavazzi et al. | Heart | 3A and B | 259 | Viral particles | Vesicular structures with surface projections, cytoplasm | CCV | insufficient | insufficient | no | see above | x | x | |
| 99 | Tchana-Sato et al. [146] | Heart | 3A | 260 | Particles whose morphology is compatible with coronavirus particles | Vesicular structures within putative membrane compartment | Unidentifiable structure/s | insufficient | insufficient | no | NA | o | o | |
| 99 | Tchana-Sato et al. | Heart | 3B | 261 | Particles whose morphology is compatible with coronavirus particles | Coated vesicle, cytoplasm | CCV | insufficient* | insufficient | no | NA | o | o | |
| 100 | Tuccari et al. [147] | Lung | 3B | 262 | Viral particles | Round to oval granular dark | Unidentifiable structure/s | insufficient | insufficient | no | NA | o | o | Perhaps profiles of small mitochondria |

| | | | | | | | | | | | | | |
|---|---|---|---|---|---|---|---|---|---|---|---|---|---|
| | | | | | | structures, cytoplasm | | | | | | | |
| 101 | Valdespino-Vázquez et al. [148] | Lung | 1C and D | 263 | Coronavirus particles, virions particles | Vesicular, partly irregularly shaped structures within membrane structure | Mitochondrion /ia | insufficient | insufficient | no | NA | o | o | |
| 101 | Valdespino-Vázquez et al. | Lung | 1E | 264 | Virions | Vesicular, partly irregularly shaped structures within membrane structure | Mitochondrion /ia | insufficient | insufficient | no | NA | o | o | |
| 101 | Valdespino-Vázquez et al. | Placent a | 1F and G | 265 | Virion particles, particles consistent with the typical morphological features of coronavirus | Dark rounded structures | Unidentifiable structure/s | insufficient | insufficient | no | NA | o | o | |
| 101 | Valdespino-Vázquez et al. | Placent a | 1H | 266 | Virion particles, particles consistent with the typical morphological features of coronavirus | Vesicular structure with empty lumen | Unidentifiable structure/s | insufficient | insufficient | no | NA | o | o | |
| 102 | Varga et al. [149] | Kidney | A and B | 267 | Viral inclusion bodies, viral particles | Vesicular structures with empty appearing interior | Unidentifiable structure/s | insufficient | insufficient | no | Goldsmith et al. [150] Varga et al. [151] (reply to Goldsmith et al.) Roufosse et al.* [83] | x | x | *) Other publications were also discussed |
| 102 | Varga et al. | Kidney | B (inset) | 268 | Viral particles, viral particle | Vesicular structure with granular appearance of membrane and empty appearing interior | rER | insufficient | insufficient | no | see above | x | x | |
| 103 | Wang et al. (liver) [152] | Liver | 1M | 269 | Coronavirus particles | Numerous vesicular structures, some with granular appearance of membrane, cytoplasm | rER | insufficient | insufficient | no | Bangash et al. [153] Wang et al. [154] (reply to Bangash et al.) | x | x | |
| 103 | Wang et al. (liver) | Liver | 2J | 270 | Coronavirus particles | Numerous vesicular structures, some with granular appearance of membrane, cytoplasm | rER | insufficient | insufficient | no | see above | x | x | |
| 104 | Wang et al. (lung and kidney) [155] | Lung | 1E (lung, | 271 | Virions | Round, dark, vesicular structure | Unidentifiable structure/s | insufficient | insufficient | no | NA | o | o | Appears to be osmicated sample, not described in methods |

| | | | | | | | | | | | | | | |
|---|---|---|---|---|---|---|---|---|---|---|---|---|---|---|
| | | | left image) | | | | | | | | | | | |
| 104 | Wang et al. (lung and kidney) | Lung | 1E (lung, middle and right images) | 272 | Virions | Dark particles (immunogold double labeling) on granular background | Immunogold, unidentifiable structure/s | insufficient | insufficient | no | NA | o | o | |
| 104 | Wang et al. (lung and kidney) | Kidney | 1E (kidney, left image) | 273 | Virions | Round, vesicular structure | Unidentifiable structure/s | insufficient | insufficient | no | NA | o | o | Appears to be osmicated sample, not described in methods |
| 104 | Wang et al. (lung and kidney) | Kidney | 1E (kidney, middle and right images) | 274 | Virions | Dark particles (immunogold double labeling) on granular background | Immunogold, unidentifiable structure/s | insufficient | insufficient | no | NA | o | o | |
| 105 | Werion et al. [48] | Kidney | 3A (with inset) | 275 | Particles resembling coronaviruses | Vesicular structures with dark granula beneath membrane and within lumen | rER | sufficient* | sufficient | no | NA | x | x | |
| 105 | Werion et al. | Kidney | 3B (with insets) | 276 | Particles resembling coronaviruses | Vesicular structures with dark granula beneath membrane and within lumen | rER | sufficient* | sufficient | no | NA | x | x | |
| 105 | Werion et al. | Kidney | 3C (with insets) | 277 | Particles resembling coronaviruses | Vesicular structures with dark granula beneath membrane and within lumen | rER | sufficient* | sufficient | no | NA | x | x | |
| 106 | Wu et al. [156] | Lung | 6 | 278 | Coronavirus particles | Round structures | Unidentifiable structure/s | insufficient | insufficient | no | NA | o | o | Paper written in chinese, abstract in english |
| 107 | Wu et al. (International Journal of Infectious Diseases) [157] | Lung | 2D | 279 | Virus, virions | Vesicular structures with relatively homogeneous interior | Unidentifiable structure/s | insufficient | sufficient | no | NA | o | o | |
| 108 | Wu et al. (JIM) [158] | Lung | 6F | 280 | Virions, viral particles with morphology and intravacuolar localization | Vesicular structures | Unidentifiable structure/s | insufficient | insufficient | no | NA | o | o | |

| | | | | | | | | | | | | | | |
|---|---|---|---|---|---|---|---|---|---|---|---|---|---|---|
| | | | | | coherent with coronavirus | | | | | | | | | |
| 108 | Wu et al. (JIM) | Lung | 6G | 281 | Virions, viral particles with morphology and intravacuolar localization coherent with coronavirus | Vesicular structures | Unidentifiable structure/s | insufficient | insufficient | no | NA | o | o | |
| 109 | Xiang et al. [159] | Lymph node | 2B | 282 | Coronavirus-like particles | Multiple dark vesicular structures | Unidentifiable structure/s | insufficient | insufficient | no | NA | o | o | |
| 110 | Xu et al. [160] | Lung | 4 | 283 | Coronavirus particles | Round structure | Unidentifiable structure/s | insufficient | insufficient | no | NA | o | o | Paper written in chinese, abstract in english |
| 111 | Yang et al. [161] | BAL | 3 | 284 | Viral granules | Dark structures within membrane compartments | Unidentifiable structure/s | insufficient* | insufficient | no | NA | o | o | |
| 112 | Yao et al. [162] | Lung | 4B | 285 | Coronavirus particles | Coated vesicles, cytoplasm | CCV | insufficient | insufficient | no | NA | o | o | Paper written in chinese, abstract in english |
| 113 | Yao et al. (Cell Research 2020) [163] | Lung | 1A (right panel) | 286 | Virus particles | Vesicular structures with surface projections, cytoplasm | CCV | sufficient | sufficient | no | NA | x | x | |
| 113 | Yao et al. (Cell Research 2020) | Lung | 1B | 287 | Virus particles | Vesicular structures with surface projections, cytoplasm | CCV | sufficient | sufficient | no | NA | x | x | |
| 114 | Yao et al. (Cell Research 2021) [164] | Lung | S2D | 288 | SARS-CoV-2 | Coated vesicle, cytoplasm | CCV | sufficient* | sufficient | no | NA | o | o | |
| 115 | Zhao et al. (BAL) [165] | BAL | 4A | 289 | Virions | Vesicular structures in membrane compartment | Unidentifiable structure/s | sufficient | insufficient | no | NA | o | o | |
| 115 | Zhao et al. (BAL) | BAL | 4B | 290 | Virions | Numerous vesicular structures in membrane compartments | Unidentifiable structure/s | sufficient | insufficient | no | NA | o | o | |
| 116 | Zhao et al. (liver) [166] | Liver | 3E | 291 | COVID-19 virus, virus particles (…) consistent with the morphology of SARS coronavirus | Vesicular structure within membrane formation | Mitochondrion /ia | sufficient | insufficient | no | NA | o | o | |
| 116 | Zhao et al. (liver) | Liver | 3F | 292 | COVID-19 virus, virion-like particle | Vesicular structure within membrane formation | Unidentifiable structure/s | sufficient | insufficient | no | NA | o | o | |

We adapted the structure of the table published by Bullock et al. [61] for our table and also implemented and/or modified their morphological terms to describe the particular structures. The “reported diagnosis” category lists citations of the terms that were used to describe the structures by the authors of the respective publication in the figure legend and/or text, multiple terms are separated by a “,”, text parts within the respective descriptions that were left out by us are marked with (…). *) Asterisk in field for structural preservation means that only limited ultrastructural detail was visible for validation. The term coated vesicle was also used as a morphological description. CCV is regarded as a broad category of coated vesicles and not limited to e.g. clathrin-coated vesicles. Abbreviations:

BAL bronchoalveolar lavage; CCV (clathrin-) coated vesicle; CoV coronavirus; ETA endotracheal aspirate; NA not available/applicable; rER rough endoplasmic reticulum; RNP ribonucleoprotein; SARS-CoV-2 severe acute respiratory syndrome coronavirus 2; x publication discussed; o publication not discussed

**Supplementary Table 9.** Publications in scientific journals demonstrating ultrastructural evidence of coronavirus particles in human samples (SARS-CoV-2)

| Nr. | Reference | Tissue | Paper type | Figure | EM-image nr. | Reported diagnosis | Structural preservation | Image presentation/ quality | All relevant structural features of CoV | Response | Bullock et al. | Hopfer et al. | Comment |
|---|---|---|---|---|---|---|---|---|---|---|---|---|---|
| 1 | Martines et al.[167] | Lung | Article | 4A (top) | 1 | Extracellular virions | sufficient* | sufficient | yes | NA | x | x | |
| 1 | Martines et al. | Lung | Article | 4A (bottom) | 2 | Extracellular virions | sufficient* | sufficient | yes | NA | x | x | |
| 1 | Martines et al. | Trachea** | Article | 4B, 5A and C | 3 | Extracellular virions | sufficient* | sufficient | yes | NA | x | x | FFPE (block) |
| 1 | Martines et al. | Lung | Article | 4C and D | 4 | Viral particles | sufficient | sufficient | yes | NA | x | x | |
| 1 | Martines et al. | Lung | Article | 4E | 5 | Viral particles | sufficient | sufficient | yes | NA | x | x | |
| 1 | Martines et al. | Lung | Article | 4F including inset | 6 | Viral particles | sufficient | sufficient | yes | NA | x | x | |
| 1 | Martines et al. | Trachea** | Article | 5B including inset | 7 | Viral particles | sufficient | sufficient | yes | NA | x | x | FFPE (section) |
| 1 | Martines et al. | Trachea** | Article | 5D including inset | 8 | Viral particles | sufficient* | sufficient | yes | NA | x | x | FFPE (section) |
| 2 | Meinhardt et al.[4] | Olfactory mucosa | Article | 3D to F | 9 | CoV particles | sufficient | sufficient | yes | NA | x | o | FFPE (block) |
| 3 | Dittmayer et al.[5] | Lung*** | Correspondence | A | 10 | SARS-CoV-2 particles | sufficient | sufficient | yes | NA | x | o | |
| 3 | Dittmayer et al. | Lung*** | Correspondence | B | 11 | SARS-CoV-2 particles | sufficient | sufficient | yes | NA | x | o | |
| 3 | Dittmayer et al. | Lung*** | Correspondence | C | 12 | SARS-CoV-2 particles | sufficient | sufficient | yes | NA | x | o | |
| 3 | Dittmayer et al. | Lung*** | Correspondence | D | 13 | SARS-CoV-2 particles | sufficient | sufficient | yes | NA | x | o | |
| 4 | Dittmayer et al.[6] | Lung*** | Reply | A to C | 14 | SARS-CoV-2 particles | sufficient | sufficient | yes | NA | o | o | |
| 5 | Ritschel et al.[168] | Lung*** | Review | 6A and B | 15 | Virus particles (german) | sufficient | sufficient | yes | NA | o | o | |
| 6 | Bullock et al.[61] | NA/ autopsy specimen, probably lung**** | Review | 1B | 16 | Viral particles | sufficient* | sufficient | yes | NA | NA | o | |
| 6 | Bullock et al. | NA/ autopsy specimen, probably lung | Review | 1C | 17 | Viral particles | sufficient* | sufficient | yes | NA | NA | o | |
| 6 | Bullock et al. | Trachea | Review | 3B | 18 | SARS-CoV-2 particles | sufficient* | sufficient | yes | NA | NA | o | FFPE (section) |
| 6 | Bullock et al. | Trachea | Review | 3C | 19 | SARS-CoV-2, viral particles | sufficient* | sufficient | yes | NA | NA | o | FFPE (block) |

*) Only limited ultrastructural detail was visible for validation; **) Described as “upper airway” in this paper and “trachea” in Bullock et al., we harmonized the terms for clarity; ***) Material of the same patient was used, ****) Probably material of the same patient(s) of Martines et al. was used
Abbreviations: CoV coronavirus; FFPE formalin-fixed paraffin-embedded; SARS-CoV-2 severe acute respiratory syndrome coronavirus 2

**Supplementary Table 10**. Publications in scientific journals discussing ultrastructural findings of putative SARS-CoV-2 particles in human samples (comments and replies are also listed in Supplementary Table 8)

| Nr. | Original reference | Journal | Directed to or discussing/ or category | Title |
|---|---|---|---|---|
| 1 | Ackermann et al. [38] | The New England Journal of Medicine | Miller [37] | Visualization of SARS-CoV-2 in the Lung – The authors reply |
| 2 | Ackermann et al.* [36] | The New England Journal of Medicine | Scholkmann and Nicholls [35] | Pulmonary Vascular Pathology in Covid-19 – The authors reply |
| 3 | Akilesh et al. [169] | The American Journal of Pathology | Review | Characterizing Viral Infection by Electron Microscopy Lessons from the Coronavirus Disease 2019 Pandemic |
| 4 | Algarroba et al. [43] | American Journal of Obstetrics & Gynecology | Letter to the editor | Confirmatory evidence of the visualization of severe acute respiratory syndrome coronavirus 2 invading the human placenta using electron microscopy |
| 5 | Algarroba et al.* [42] | American Journal of Obstetrics & Gynecology | Kniss [41] | Reply |
| 6 | Bangash et al. [153] | Journal of Hepatology | Wang et al. [154] | SARS-CoV-2: Is the liver merely a bystander to severe disease? |
| 7 | Baeck et al. [66] | British Journal of Dermatology | Colmenero et al. [67] | Chilblains and COVID-19: why SARS-CoV-2 endothelial infection is questioned |
| 8 | Brealey and Miller [68] | British Journal of Dermatology | Colmenero et al. [69] | SARS-CoV-2 has not been detected directly by electron microscopy in the endothelium of chilblain lesions |
| 9 | Buja [55] | Cardiovascular Pathology | Giannico and Miller [54] | Electron microscopic identification of SARS-CoV-2 |
| 10 | Bullock et al.* [61] | Emerging Infectious Diseases | Review | Difficulties in Differentiating Coronaviruses from Subcellular Structures in Human Tissues by Electron Microscopy |
| 11 | Bullock et al. [170] | Kidney International | Review | Best practices for correctly identifying coronavirus by transmission electron microscopy |
| 12 | Calomeni et al. [105] | Kidney International | Su et al. [143], Kissling et al. [104] | Multivesicular bodies mimicking SARS-CoV-2 in patients without COVID-19 |
| 13 | Cassol et al. [171] | Kidney 360 | Brief communication | Appearances Can Be Deceiving – Viral-like Inclusions in COVID-19 Negative Renal Biopsies by Electron Microscopy |
| 14 | Colmenero et al. [67] | British Journal of Dermatology | Baeck et al. [66] | Chilblains and COVID-19: why SARS-CoV-2 endothelial infection is questioned. Reply from the authors |
| 15 | Colmenero et al. [69] | British Journal of Dermatology | Brealey and Miller [68] | SARS-CoV-2 Has Not Been Detected Directly by Electron Microscopy in the Endothelium of Chilblain Lesions: reply from authors |
| 16 | Delsante et al. [81] | Journal of the American Society of Nephrology | Farkash et al. [79] | Kidney Involvement in COVID-19: Need for Better Definitions |
| 17 | Dittmayer et al.* [6] | The Lancet | Dolhnikoff et al. [74] | Authors' reply |
| 18 | Dittmayer et al.* [5] | The Lancet | Bradley et al. [51], Dolhnikoff et al. [73] | Why misinterpretation of electron micrographs in SARS-CoV-2-infected tissue goes viral |
| 19 | Dolhnikoff et al. [74] | The Lancet | Dittmayer et al. [5] | Using EM data to understand COVID-19 pathophysiology |
| 20 | Farkash et al. [82] | Journal of the American Society of Nephrology | Delsante et al. [81], Miller and Goldsmith [80] | Authors' Reply |
| 21 | Frelih et al. [172] | KIReports | Letter to the editor | SARS-CoV-2 Virions or Ubiquitous Cell Structures? Actual Dilemma in COVID-19 Era |
| 22 | Giannico and Miller [54] | Cardiovascular Pathology | Buja et al. [53] | Electron microscopy identification of SARS-COV-2: what is the evidence? |
| 23 | Goldsmith et al. [150] | The Lancet | Varga et al. [149] | Electron microscopy of SARS-CoV-2: a challenging task |
| 24 | Hopfer et al. [11] | Histopathology | Review | Hunting coronavirus by transmission electron microscopy – a guide to SARS-CoV-2-associated ultrastructural pathology in COVID-19 tissues |
| 25 | Kissling et al. [107] | Kidney International | Miller and Brealey [106] | The authors reply |
| 26 | Kniss [41] | American Journal of Obstetrics & Gynecology | Algarroba et al. [40] | Alternative interpretation to the findings reported in visualization of severe acute respiratory syndrome coronavirus 2 invading the human placenta using electron microscopy |
| 27 | Miller [37] | The New England Journal of Medicine | Ackermann et al. [34] | Visualization of SARS-CoV-2 in the Lung – To the editor |
| 28 | Miller and Goldsmith [80] | Journal of the American Society of Nephrology | Farkash et al. [79] | Caution in Identifying Coronaviruses by Electron Microscopy |
| 29 | Miller and Brealey [106] | Kidney International | Su et al. [143], Kissling et al. [104] | Visualization of putative coronavirus in kidney |
| 30 | Neil et al. [173] | Journal of Pathology | Review | Ultrastructure of cell trafficking pathways and coronavirus: how to recognise the wolf amongst the sheep |
| 31 | Ritschel et al.* [168] | Der Pathologe | Review | COVID-19 and the central and peripheral nervous system |

| | | | | |
|---|---|---|---|---|
| 32 | Roufosse et al. [83] | Kidney International | Varga et al. [149], Su et al. [143], Farkash et al. [79], Kissling et al. [104] | Electron microscopic investigations in COVID-19: not all crowns are coronas |
| 33 | Scholkmann and Nicholls [35] | The New England Journal of Medicine | Ackermann et al. [34] | Pulmonary Vascular Pathology in Covid-19 – To the editor |
| 34 | Smith et al. [174] | Kidney International | Su et al. [143] | Am I a coronavirus? |
| 35 | Stahl et al. [141] | Intensive Care Medicine | Stahl et al. [140] | Absence of SARS-CoV-2 RNA in COVID-19-associated intestinal endothelialitis |
| 36 | Su et al. [144] | Kidney International | Miller and Brealey [106] | The authors reply |
| 37 | Varga et al. [151] | The Lancet | Goldsmith et al. [150] | Authors' reply |
| 38 | Wang et al. [154] | Journal of Hepatology | Bangash et al. [153] | Reply to: Correspondence relating to "SARS-CoV-2 infection of the liver directly contributes to hepatic impairment in patients with COVID-19" |

*) Publications that also demonstrate images of (putative) coronavirus particles in human samples, see Supplementary Tables 8 and 9 for further information. Abbreviation: SARS-CoV-2 severe acute respiratory syndrome coronavirus 2

**Supplementary Video (available on www.nanotomy.org).** The video demonstrates browsing within large-scale screening and detail datasets of human autopsy lung tissue, infected cell culture and FFPE-reembedded human autopsy olfactory mucosa. Advanced visual analysis is achieved by efficient pan-and-zoom-analysis within the coherent datasets that cannot be achieved with traditional EM that lacks a high-quality live view and similar fast navigation. Also, tools for annotation and measurements facilitate analysis.

**References**

1. Pfefferle S, Huang J, Norz D, et al. Complete Genome Sequence of a SARS-CoV-2 Strain Isolated in Northern Germany. *Microbiol Resour Announc* 2020; **9**(23).
2. Laue M, Kauter A, Hoffmann T, Moller L, Michel J, Nitsche A. Morphometry of SARS-CoV and SARS-CoV-2 particles in ultrathin plastic sections of infected Vero cell cultures. *Sci Rep* 2021; **11**(1): 3515.
3. Fortunato F, Hackert T, Buchler MW, Kroemer G. Retrospective electron microscopy: Preservation of fine structure by freezing and aldehyde fixation. *Mol Cell Oncol* 2016; **3**(6): e1251382.
4. Meinhardt J, Radke J, Dittmayer C, et al. Olfactory transmucosal SARS-CoV-2 invasion as a port of central nervous system entry in individuals with COVID-19. *Nat Neurosci* 2021; **24**(2): 168-75.
5. Dittmayer C, Meinhardt J, Radbruch H, et al. Why misinterpretation of electron micrographs in SARS-CoV-2-infected tissue goes viral. *Lancet* 2020; **396**(10260): e64-e5.
6. Dittmayer C, Meinhardt J, Radbruch H, et al. Using EM data to understand COVID-19 pathophysiology - Authors' reply. *Lancet* 2021; **397**(10270): 197-8.
7. Laue M. Electron microscopy of viruses. *Methods Cell Biol* 2010; **96**: 1-20.
8. Dittmayer C, Goebel HH, Heppner FL, Stenzel W, Bachmann S. Preparation of Samples for Large-Scale Automated Electron Microscopy of Tissue and Cell Ultrastructure. *Microsc Microanal* 2021; **27**(4): 815-27.
9. Wong DWL, Klinkhammer BM, Djudjaj S, et al. Multisystemic Cellular Tropism of SARS-CoV-2 in Autopsies of COVID-19 Patients. *Cells* 2021; **10**(8).
10. Lean FZX, Lamers MM, Smith SP, et al. Development of immunohistochemistry and in situ hybridisation for the detection of SARS-CoV and SARS-CoV-2 in formalin-fixed paraffin-embedded specimens. *Sci Rep* 2020; **10**(1): 21894.
11. Hopfer H, Herzig MC, Gosert R, et al. Hunting coronavirus by transmission electron microscopy - a guide to SARS-CoV-2-associated ultrastructural pathology in COVID-19 tissues. *Histopathology* 2021; **78**(3): 358-70.
12. Goldsmith CS, Tatti KM, Ksiazek TG, et al. Ultrastructural characterization of SARS coronavirus. *Emerg Infect Dis* 2004; **10**(2): 320-6.
13. Bian XW, Team C-P. Autopsy of COVID-19 patients in China. *Natl Sci Rev* 2020; **7**(9): 1414-8.
14. Best Rocha A, Stroberg E, Barton LM, et al. Detection of SARS-CoV-2 in formalin-fixed paraffin-embedded tissue sections using commercially available reagents. *Lab Invest* 2020; **100**(11): 1485-9.
15. Borczuk AC, Salvatore SP, Seshan SV, et al. COVID-19 pulmonary pathology: a multi-institutional autopsy cohort from Italy and New York City. *Mod Pathol* 2020; **33**(11): 2156-68.
16. Carossino M, Ip HS, Richt JA, et al. Detection of SARS-CoV-2 by RNAscope((R)) in situ hybridization and immunohistochemistry techniques. *Arch Virol* 2020; **165**(10): 2373-7.
17. Duarte-Neto AN, Caldini EG, Gomes-Gouvea MS, et al. An autopsy study of the spectrum of severe COVID-19 in children: From SARS to different phenotypes of MIS-C. *EClinicalMedicine* 2021; **35**: 100850.
18. Duarte-Neto AN, Teixeira TA, Caldini EG, et al. Testicular pathology in fatal COVID-19: A descriptive autopsy study. *Andrology* 2021.
19. El Jamal SM, Pujadas E, Ramos I, et al. Tissue-based SARS-CoV-2 detection in fatal COVID-19 infections: Sustained direct viral-induced damage is not necessary to drive disease progression. *Hum Pathol* 2021; **114**: 110-9.
20. Hong J, Wang Q, Wu Q, et al. Rabbit Monoclonal Antibody Specifically Recognizing a Linear Epitope in the RBD of SARS-CoV-2 Spike Protein. *Vaccines (Basel)* 2021; **9**(8).
21. Ko CJ, Harigopal M, Gehlhausen JR, Bosenberg M, McNiff JM, Damsky W. Discordant anti-SARS-CoV-2 spike protein and RNA staining in cutaneous perniotic lesions suggests endothelial deposition of cleaved spike protein. *J Cutan Pathol* 2021; **48**(1): 47-52.
22. Lacavalla D, Santandrea G, Andreotti D, Stano R, Occhionorelli S. Case report of gastrointestinal localization of SARS-CoV-2 and open abdomen technique in an Italian emergency surgery department for gastrointestinal bleeding. *Ann Med Surg (Lond)* 2021; **66**: 102405.

23. Liu J, Babka AM, Kearney BJ, Radoshitzky SR, Kuhn JH, Zeng X. Molecular detection of SARS-CoV-2 in formalin-fixed, paraffin-embedded specimens. *JCI Insight* 2020; **5**(12).
24. Massoth LR, Desai N, Szabolcs A, et al. Comparison of RNA In Situ Hybridization and Immunohistochemistry Techniques for the Detection and Localization of SARS-CoV-2 in Human Tissues. *Am J Surg Pathol* 2021; **45**(1): 14-24.
25. McMullen P, Pytel P, Snyder A, et al. A series of COVID-19 autopsies with clinical and pathologic comparisons to both seasonal and pandemic influenza. *J Pathol Clin Res* 2021.
26. Ray A, Jain D, Goel A, et al. Clinico-pathological features in fatal COVID-19 infection: a preliminary experience of a tertiary care center in North India using postmortem minimally invasive tissue sampling. *Expert Rev Respir Med* 2021: 1-9.
27. Roden AC, Vrana JA, Koepplin JW, et al. Comparison of In Situ Hybridization, Immunohistochemistry, and Reverse Transcription-Droplet Digital Polymerase Chain Reaction for Severe Acute Respiratory Syndrome Coronavirus 2 (SARS-CoV-2) Testing in Tissue. *Arch Pathol Lab Med* 2021; **145**(7): 785-96.
28. Sauter JL, Baine MK, Butnor KJ, et al. Insights into pathogenesis of fatal COVID-19 pneumonia from histopathology with immunohistochemical and viral RNA studies. *Histopathology* 2020; **77**(6): 915-25.
29. Szabolcs M, Sauter JL, Frosina D, et al. Identification of Immunohistochemical Reagents for In Situ Protein Expression Analysis of Coronavirus-associated Changes in Human Tissues. *Appl Immunohistochem Mol Morphol* 2021; **29**(1): 5-12.
30. Teixeira Junior AAL, Neves P, Lages JS, et al. Brazilian Consortium for the Study on Renal Diseases Associated With COVID-19: A Multicentric Effort to Understand SARS-CoV-2-Related Nephropathy. *Front Med (Lausanne)* 2020; **7**: 584235.
31. Abbate M, Rottoli D, Gianatti A. COVID-19 Attacks the Kidney: Ultrastructural Evidence for the Presence of Virus in the Glomerular Epithelium. *Nephron* 2020; **144**(7): 341-2.
32. Abdullaev A, Odilov A, Ershler M, et al. Viral Load and Patterns of SARS-CoV-2 Dissemination to the Lungs, Mediastinal Lymph Nodes, and Spleen of Patients with COVID-19 Associated Lymphopenia. *Viruses* 2021; **13**(7).
33. Achua JK, Chu KY, Ibrahim E, et al. Histopathology and Ultrastructural Findings of Fatal COVID-19 Infections on Testis. *World J Mens Health* 2021; **39**(1): 65-74.
34. Ackermann M, Verleden SE, Kuehnel M, et al. Pulmonary Vascular Endothelialitis, Thrombosis, and Angiogenesis in Covid-19. *N Engl J Med* 2020; **383**(2): 120-8.
35. Scholkmann F, Nicholls J. Pulmonary Vascular Pathology in Covid-19. *N Engl J Med* 2020; **383**(9): 887-8.
36. Ackermann M, Mentzer SJ, Jonigk D. Pulmonary Vascular Pathology in Covid-19. Reply. *N Engl J Med* 2020; **383**(9): 888-9.
37. Miller SE. Visualization of SARS-CoV-2 in the Lung. *N Engl J Med* 2020; **383**(27): 2689.
38. Ackermann M, Mentzer SJ, Jonigk D. Visualization of SARS-CoV-2 in the Lung. Reply. *N Engl J Med* 2020; **383**(27): 2689-90.
39. Albert CL, Carmona-Rubio AE, Weiss AJ, Procop GG, Starling RC, Rodriguez ER. The Enemy Within: Sudden-Onset Reversible Cardiogenic Shock With Biopsy-Proven Cardiac Myocyte Infection by Severe Acute Respiratory Syndrome Coronavirus 2. *Circulation* 2020; **142**(19): 1865-70.
40. Algarroba GN, Rekawek P, Vahanian SA, et al. Visualization of severe acute respiratory syndrome coronavirus 2 invading the human placenta using electron microscopy. *Am J Obstet Gynecol* 2020; **223**(2): 275-8.
41. Kniss DA. Alternative interpretation to the findings reported in visualization of severe acute respiratory syndrome coronavirus 2 invading the human placenta using electron microscopy. *Am J Obstet Gynecol* 2020; **223**(5): 785-6.
42. Algarroba GN, Rekawek P, Vahanian SA, et al. Reply. *Am J Obstet Gynecol* 2020; **223**(5): 786-8.

43. Algarroba GN, Hanna NN, Rekawek P, et al. Confirmatory evidence of the visualization of severe acute respiratory syndrome coronavirus 2 invading the human placenta using electron microscopy. *Am J Obstet Gynecol* 2020; **223**(6): 953-4.
44. Anandh U, Gowrishankar S, Sharma A, Salama A, Dasgupta I. Kidney transplant dysfunction in a patient with COVID - 19 infection: role of concurrent Sars-Cov 2 nephropathy, chronic rejection and vitamin C-mediated hyperoxalosis: case report. *BMC Nephrol* 2021; **22**(1): 91.
45. Araujo-Silva CA, Marcos AAA, Marinho PM, et al. Presumed SARS-CoV-2 Viral Particles in the Human Retina of Patients With COVID-19. *JAMA Ophthalmol* 2021.
46. Bain WG, Penaloza HF, Ladinsky MS, et al. Lower Respiratory Tract Myeloid Cells Harbor SARS-Cov-2 and Display an Inflammatory Phenotype. *Chest* 2021; **159**(3): 963-6.
47. Birkhead M, Glass AJ, Allan-Gould H, Goossens C, Wright CA. Ultrastructural evidence for vertical transmission of SARS-CoV-2. *Int J Infect Dis* 2021; **111**: 10-1.
48. Werion A, Belkhir L, Perrot M, et al. SARS-CoV-2 causes a specific dysfunction of the kidney proximal tubule. *Kidney Int* 2020; **98**(5): 1296-307.
49. Bojkova D, Wagner JUG, Shumliakivska M, et al. SARS-CoV-2 infects and induces cytotoxic effects in human cardiomyocytes. *Cardiovasc Res* 2020; **116**(14): 2207-15.
50. Bosmuller H, Traxler S, Bitzer M, et al. The evolution of pulmonary pathology in fatal COVID-19 disease: an autopsy study with clinical correlation. *Virchows Arch* 2020; **477**(3): 349-57.
51. Bradley BT, Maioli H, Johnston R, et al. Histopathology and ultrastructural findings of fatal COVID-19 infections in Washington State: a case series. *Lancet* 2020; **396**(10247): 320-32.
52. Bryce C, Grimes Z, Pujadas E, et al. Pathophysiology of SARS-CoV-2: the Mount Sinai COVID-19 autopsy experience. *Mod Pathol* 2021.
53. Buja LM, Wolf DA, Zhao B, et al. The emerging spectrum of cardiopulmonary pathology of the coronavirus disease 2019 (COVID-19): Report of 3 autopsies from Houston, Texas, and review of autopsy findings from other United States cities. *Cardiovasc Pathol* 2020; **48**: 107233.
54. Giannico GA, Miller SE. Electron microscopy identification of SARS-COV-2: what is the evidence? *Cardiovasc Pathol* 2021; **52**: 107338.
55. Buja LM. Electron microscopic identification of SARS-CoV-2. *Cardiovasc Pathol* 2021; **52**: 107337.
56. Bulfamante G, Chiumello D, Canevini MP, et al. First ultrastructural autoptic findings of SARS - Cov-2 in olfactory pathways and brainstem. *Minerva Anestesiol* 2020; **86**(6): 678-9.
57. Bulfamante GP, Perrucci GL, Falleni M, et al. Evidence of SARS-CoV-2 Transcriptional Activity in Cardiomyocytes of COVID-19 Patients without Clinical Signs of Cardiac Involvement. *Biomedicines* 2020; **8**(12).
58. Calabrese F, Pezzuto F, Fortarezza F, et al. Pulmonary pathology and COVID-19: lessons from autopsy. The experience of European Pulmonary Pathologists. *Virchows Arch* 2020; **477**(3): 359-72.
59. Canini V, Bono F, Calzavacca P, et al. Cytopathology of bronchoalveolar lavages in COVID___19 pneumonia: A pilot study. *Cancer Cytopathol* 2021; **129**(8): 632-41.
60. Carsana L, Sonzogni A, Nasr A, et al. Pulmonary post-mortem findings in a series of COVID-19 cases from northern Italy: a two-centre descriptive study. *Lancet Infect Dis* 2020; **20**(10): 1135-40.
61. Bullock HA, Goldsmith CS, Zaki SR, Martines RB, Miller SE. Difficulties in Differentiating Coronaviruses from Subcellular Structures in Human Tissues by Electron Microscopy. *Emerg Infect Dis* 2021; **27**(4): 1023-31.
62. Cazzato G, Mazzia G, Cimmino A, et al. SARS-CoV-2 and Skin: The Pathologist's Point of View. *Biomolecules* 2021; **11**(6).
63. Chaudhary S, Rai P, Sesham K, et al. Microscopic imaging of bronchoalveolar fluids of COVID-19 positive intubated patients reveals the different level of SARS-CoV-2 infection on oral squamosal epithelial cells. *Indian Journal of Biochemistry & Biophysics* 2021; **58**: 196-207.
64. Chen XJ, Li K, Xu L, et al. Novel insight from the first lung transplant of a COVID-19 patient. *Eur J Clin Invest* 2021; **51**(1): e13443.

65. Colmenero I, Santonja C, Alonso-Riano M, et al. SARS-CoV-2 endothelial infection causes COVID-19 chilblains: histopathological, immunohistochemical and ultrastructural study of seven paediatric cases. *Br J Dermatol* 2020; **183**(4): 729-37.
66. Baeck M, Hoton D, Marot L, Herman A. Chilblains and COVID-19: why SARS-CoV-2 endothelial infection is questioned. *Br J Dermatol* 2020; **183**(6): 1152-3.
67. Colmenero I, Santonja C, Alonso-Riano M, et al. Chilblains and COVID-19: why SARS-CoV-2 endothelial infection is questioned. Reply from the authors. *Br J Dermatol* 2020; **183**(6): 1153-4.
68. Brealey JK, Miller SE. SARS-CoV-2 has not been detected directly by electron microscopy in the endothelium of chilblain lesions. *Br J Dermatol* 2021; **184**(1): 186.
69. Colmenero I, Santonja C, Alonso-Riano M, et al. SARS-CoV-2 Has Not Been Detected Directly by Electron Microscopy in the Endothelium of Chilblain Lesions: reply from authors. *Br J Dermatol* 2020.
70. Debelenko L, Katsyv I, Chong AM, Peruyero L, Szabolcs M, Uhlemann AC. Trophoblast damage with acute and chronic intervillositis: disruption of the placental barrier by severe acute respiratory syndrome coronavirus 2. *Hum Pathol* 2021; **109**: 69-79.
71. Deinhardt-Emmer S, Wittschieber D, Sanft J, et al. Early postmortem mapping of SARS-CoV-2 RNA in patients with COVID-19 and the correlation with tissue damage. *Elife* 2021; **10**.
72. Deshmukh S, Zhou XJ, Hiser W. Collapsing glomerulopathy in a patient of Indian descent in the setting of COVID-19. *Ren Fail* 2020; **42**(1): 877-80.
73. Dolhnikoff M, Ferreira Ferranti J, de Almeida Monteiro RA, et al. SARS-CoV-2 in cardiac tissue of a child with COVID-19-related multisystem inflammatory syndrome. *Lancet Child Adolesc Health* 2020; **4**(10): 790-4.
74. Dolhnikoff M, Duarte-Neto AN, Saldiva PHN, Caldini EG. Using EM data to understand COVID-19 pathophysiology. *Lancet* 2021; **397**(10270): 196-7.
75. Erman A, Wechtersbach K, Velkavrh D, Plesko J, Frelih M, Kojc N. Just Seeing Is Not Enough for Believing: Immunolabelling as Indisputable Proof of SARS-CoV-2 Virions in Infected Tissue. *Viruses* 2021; **13**(9).
76. Evert K, Dienemann T, Brochhausen C, et al. Autopsy findings after long-term treatment of COVID-19 patients with microbiological correlation. *Virchows Arch* 2021; **479**(1): 97-108.
77. Facchetti F, Bugatti M, Drera E, et al. SARS-CoV2 vertical transmission with adverse effects on the newborn revealed through integrated immunohistochemical, electron microscopy and molecular analyses of Placenta. *EBioMedicine* 2020; **59**: 102951.
78. Falasca L, Nardacci R, Colombo D, et al. Postmortem Findings in Italian Patients With COVID-19: A Descriptive Full Autopsy Study of Cases With and Without Comorbidities. *J Infect Dis* 2020; **222**(11): 1807-15.
79. Farkash EA, Wilson AM, Jentzen JM. Ultrastructural Evidence for Direct Renal Infection with SARS-CoV-2. *J Am Soc Nephrol* 2020; **31**(8): 1683-7.
80. Miller SE, Goldsmith CS. Caution in Identifying Coronaviruses by Electron Microscopy. *J Am Soc Nephrol* 2020; **31**(9): 2223-4.
81. Delsante M, Rossi GM, Gandolfini I, Bagnasco SM, Rosenberg AZ. Kidney Involvement in COVID-19: Need for Better Definitions. *J Am Soc Nephrol* 2020; **31**(9): 2224-5.
82. Farkash EA, Wilson AM, Jentzen JM. Authors' Reply. *J Am Soc Nephrol* 2020; **31**(9): 2225-6.
83. Roufosse C, Curtis E, Moran L, et al. Electron microscopic investigations in COVID-19: not all crowns are coronas. *Kidney Int* 2020; **98**(2): 505-6.
84. Fiel MI, El Jamal SM, Paniz-Mondolfi A, et al. Findings of Hepatic Severe Acute Respiratory Syndrome Coronavirus-2 Infection. *Cell Mol Gastroenterol Hepatol* 2021; **11**(3): 763-70.
85. Fox SE, Li G, Akmatbekov A, et al. Unexpected Features of Cardiac Pathology in COVID-19 Infection. *Circulation* 2020; **142**(11): 1123-5.
86. Fox SE, Akmatbekov A, Harbert JL, Li G, Quincy Brown J, Vander Heide RS. Pulmonary and cardiac pathology in African American patients with COVID-19: an autopsy series from New Orleans. *Lancet Respir Med* 2020; **8**(7): 681-6.

87. Garau G, Joachim S, Duliere GL, et al. Sudden cardiogenic shock mimicking fulminant myocarditis in a surviving teenager affected by severe acute respiratory syndrome coronavirus 2 infection. *ESC Heart Fail* 2021; **8**(1): 766-73.
88. Garcia-Cruz E, Manzur-Sandoval D, Lazcano-Diaz EA, Soria-Castro E, Jimenez-Becerra S. Cardiac Tamponade in a Patient With Myocardial Infarction and COVID-19: Electron Microscopy. *JACC Case Rep* 2020; **2**(12): 2021-3.
89. Garrido Ruiz MC, Santos-Briz A, Santos-Briz A, et al. Spectrum of Clinicopathologic Findings in COVID-19-induced Skin Lesions: Demonstration of Direct Viral Infection of the Endothelial Cells. *Am J Surg Pathol* 2021; **45**(3): 293-303.
90. Gioia CD, Zullo F, Vecchio RCB, et al. Stillbirth and fetal capillary infection by SARS-CoV-2. *Am J Obstet Gynecol MFM* 2021: 100523.
91. Grimes Z, Bryce C, Sordillo EM, et al. Fatal Pulmonary Thromboembolism in SARS-CoV-2-Infection. *Cardiovasc Pathol* 2020; **48**: 107227.
92. Hakroush S, Franz J, Larsen J, Korsten P, Winkler MS, Tampe B. Repeated false-negative tests delayed diagnosis of COVID-19 in a case with granulomatosis with polyangiitis under maintenance therapy with rituximab and concomitant influenza pneumonia. *Ann Rheum Dis* 2020.
93. Han Y, Duan X, Yang L, et al. Identification of SARS-CoV-2 inhibitors using lung and colonic organoids. *Nature* 2021; **589**(7841): 270-5.
94. Hooper JE, Uner M, Priemer DS, Rosenberg A, Chen L. Muscle Biopsy Findings in a Case of SARS-CoV-2-Associated Muscle Injury. *J Neuropathol Exp Neurol* 2021; **80**(4): 377-8.
95. Hosier H, Farhadian SF, Morotti RA, et al. SARS-CoV-2 infection of the placenta. *J Clin Invest* 2020; **130**(9): 4947-53.
96. Idrissi MO, Baudoin J, Chateau A, et al. Presence of SARS-CoV-2 in a Cornea Transplant *pathogens* 2021; **10**(8): 1-6.
97. Ingravallo G, Mazzotta F, Resta L, et al. Inflammatory Skin Lesions in Three SARS-CoV-2 Swab-Negative Adolescents: A Possible COVID-19 Sneaky Manifestation? *Pediatr Rep* 2021; **13**(2): 181-8.
98. Jacobs JL, Bain W, Naqvi A, et al. SARS-CoV-2 Viremia is Associated with COVID-19 Severity and Predicts Clinical Outcomes. *Clin Infect Dis* 2021.
99. Jeican, II, Aluas M, Lazar M, et al. Evidence of SARS-CoV-2 Virus in the Middle Ear of Deceased COVID-19 Patients. *Diagnostics (Basel)* 2021; **11**(9).
100. Jeican, II, Gheban D, Barbu-Tudoran L, et al. Respiratory Nasal Mucosa in Chronic Rhinosinusitis with Nasal Polyps versus COVID-19: Histopathology, Electron Microscopy Analysis and Assessing of Tissue Interleukin-33. *J Clin Med* 2021; **10**(18).
101. Kadosh BS, Pavone J, Wu M, Reyentovich A, Gidea C. Collapsing glomerulopathy associated with COVID-19 infection in a heart transplant recipient. *J Heart Lung Transplant* 2020; **39**(8): 855-7.
102. Kanczkowski W, Evert K, Stadtmüller M, et al. COVID-19 targets human adrenal glands. *The Lancet Diabetes & Endocrinology* 2021; **10**(1): 13-6.
103. Khismatullin RR, Ponomareva AA, Nagaswami C, et al. Pathology of lung-specific thrombosis and inflammation in COVID-19. *J Thromb Haemost* 2021; **19**(12): 3062-72.
104. Kissling S, Rotman S, Gerber C, et al. Collapsing glomerulopathy in a COVID-19 patient. *Kidney Int* 2020; **98**(1): 228-31.
105. Calomeni E, Satoskar A, Ayoub I, Brodsky S, Rovin BH, Nadasdy T. Multivesicular bodies mimicking SARS-CoV-2 in patients without COVID-19. *Kidney Int* 2020; **98**(1): 233-4.
106. Miller SE, Brealey JK. Visualization of putative coronavirus in kidney. *Kidney Int* 2020; **98**(1): 231-2.
107. Kissling S, Rotman S, Fakhouri F. The authors reply. *Kidney Int* 2020; **98**(1): 232.
108. Kolivras A, Thompson C, Pastushenko I, et al. A clinicopathological description of COVID-19-induced chilblains (COVID-toes) correlated with a published literature review. *J Cutan Pathol* 2021.
109. Kresch E, Achua J, Saltzman R, et al. COVID-19 Endothelial Dysfunction Can Cause Erectile Dysfunction: Histopathological, Immunohistochemical, and Ultrastructural Study of the Human Penis. *World J Mens Health* 2021; **39**(3): 466-9.

110. Lauermann P, Storch M, Weig M, et al. There is no intraocular affection on a SARS-CoV-2 - Infected ocular surface. *Am J Ophthalmol Case Rep* 2020; **20**: 100884.
111. Lei J, Liu Y, Xie T, et al. Evidence for residual SARS-CoV-2 in glioblastoma tissue of a convalescent patient. *Neuroreport* 2021; **32**(9): 771-5.
112. Leoni E, Cerati M, Finzi G, Lombardo M, Sessa F. COVID-19 and HHV8 first spotted together: an affair under electron microscopy. *J Eur Acad Dermatol Venereol* 2021; **35**(5): e311-e2.
113. Li S, Jiang L, Li X, et al. Clinical and pathological investigation of patients with severe COVID-19. *JCI Insight* 2020; **5**(12).
114. Lin H, Ma X, Xiao F, et al. Identification of a special cell type as a determinant of the kidney tropism of SARS-CoV-2. *FEBS J* 2021.
115. Liu J, Babka AM, Kearney BJ, Radoshitzky SR, Kuhn JH, Zeng X. Molecular Detection of SARS-CoV-2 in Formalin Fixed Paraffin Embedded Specimens. *bioRxiv* 2020.
116. Livanos AE, Jha D, Cossarini F, et al. Intestinal Host Response to SARS-CoV-2 Infection and COVID-19 Outcomes in Patients With Gastrointestinal Symptoms. *Gastroenterology* 2021; **160**(7): 2435-50 e34.
117. Lubnow M, Schmidt B, Fleck M, et al. Secondary hemophagocytic lymphohistiocytosis and severe liver injury induced by hepatic SARS-CoV-2 infection unmasking Wilson's disease: Balancing immunosuppression. *Int J Infect Dis* 2021; **103**: 624-7.
118. Ma X, Guan C, Chen R, et al. Pathological and molecular examinations of postmortem testis biopsies reveal SARS-CoV-2 infection in the testis and spermatogenesis damage in COVID-19 patients. *Cell Mol Immunol* 2021; **18**(2): 487-9.
119. Martin-Cardona A, Lloreta Trull J, Albero-Gonzalez R, et al. SARS-CoV-2 identified by transmission electron microscopy in lymphoproliferative and ischaemic intestinal lesions of COVID-19 patients with acute abdominal pain: two case reports. *BMC Gastroenterol* 2021; **21**(1): 334.
120. Matuck BF, Dolhnikoff M, Duarte-Neto AN, et al. Salivary glands are a target for SARS-CoV-2: a source for saliva contamination. *J Pathol* 2021; **254**(3): 239-43.
121. Menter T, Haslbauer JD, Nienhold R, et al. Postmortem examination of COVID-19 patients reveals diffuse alveolar damage with severe capillary congestion and variegated findings in lungs and other organs suggesting vascular dysfunction. *Histopathology* 2020; **77**(2): 198-209.
122. Mitchell JM, Rakheja D, Gopal P. SARS-CoV-2-related Hypercoagulable State Leading to Ischemic Enteritis Secondary to Superior Mesenteric Artery Thrombosis. *Clin Gastroenterol Hepatol* 2020.
123. Morbini P, Benazzo M, Verga L, et al. Ultrastructural Evidence of Direct Viral Damage to the Olfactory Complex in Patients Testing Positive for SARS-CoV-2. *JAMA Otolaryngol Head Neck Surg* 2020; **146**(10): 972-3.
124. Nardacci R, Colavita F, Castilletti C, et al. Evidences for lipid involvement in SARS-CoV-2 cytopathogenesis. *Cell Death Dis* 2021; **12**(3): 263.
125. Paniz-Mondolfi A, Bryce C, Grimes Z, et al. Central nervous system involvement by severe acute respiratory syndrome coronavirus-2 (SARS-CoV-2). *J Med Virol* 2020; **92**(7): 699-702.
126. Parron D, Gartzia A, Iturregui AM, et al. SARS-CoV-2-Associated Obliterative Arteritis Causing Massive Testicular Infarction. *Clin Pract* 2021; **11**(2): 246-9.
127. Perez A, Torregrosa I, D'Marco L, et al. IgA-Dominant Infection-Associated Glomerulonephritis Following SARS-CoV-2 Infection. *Viruses* 2021; **13**(4).
128. Pesaresi M, Pirani F, Tagliabracci A, et al. SARS-CoV-2 identification in lungs, heart and kidney specimens by transmission and scanning electron microscopy. *Eur Rev Med Pharmacol Sci* 2020; **24**(9): 5186-8.
129. Pirisi M, Rigamonti C, D'Alfonso S, et al. Liver infection and COVID-19: the electron microscopy proof and revision of the literature. *Eur Rev Med Pharmacol Sci* 2021; **25**(4): 2146-51.
130. Prieto-Perez L, Fortes J, Soto C, et al. Histiocytic hyperplasia with hemophagocytosis and acute alveolar damage in COVID-19 infection. *Mod Pathol* 2020; **33**(11): 2139-46.
131. Qadir MMF, Bhondeley M, Beatty W, et al. SARS-CoV-2 infection of the pancreas promotes thrombofibrosis and is associated with new-onset diabetes. *JCI Insight* 2021; **6**(16).

132. Qian Q, Fan L, Liu W, et al. Direct evidence of active SARS-CoV-2 replication in the intestine. *Clin Infect Dis* 2020.
133. Rassaf T, Totzeck M, Mahabadi AA, et al. Ventricular assist device for a coronavirus disease 2019-affected heart. *ESC Heart Fail* 2021; **8**(1): 162-6.
134. Ren L, Liu Q, Wang R, et al. Clinicopathologic Features of COVID-19: A Case Report and Value of Forensic Autopsy in Studying SARS-CoV-2 Infection. *Am J Forensic Med Pathol* 2021; **42**(2): 164-9.
135. Resta L, Vimercati A, Cazzato G, et al. SARS-CoV-2 and Placenta: New Insights and Perspectives. *Viruses* 2021; **13**(5).
136. Santana MF, Pinto RAA, Marcon BH, et al. Pathological findings and morphologic correlation of the lungs of autopsied patients with SARS-CoV-2 infection in the Brazilian Amazon using transmission electron microscopy. *Rev Soc Bras Med Trop* 2021; **54**: e0850.
137. Schoenmakers S, Snijder P, Verdijk RM, et al. Severe Acute Respiratory Syndrome Coronavirus 2 Placental Infection and Inflammation Leading to Fetal Distress and Neonatal Multi-Organ Failure in an Asymptomatic Woman. *J Pediatric Infect Dis Soc* 2021; **10**(5): 556-61.
138. Schwab K, Hamidi S, Chung A, et al. Occult Colonic Perforation in a Patient With Coronavirus Disease 2019 After Interleukin-6 Receptor Antagonist Therapy. *Open Forum Infect Dis* 2020; **7**(11): ofaa424.
139. Sisman J, Jaleel MA, Moreno W, et al. Intrauterine Transmission of SARS-COV-2 Infection in a Preterm Infant. *Pediatr Infect Dis J* 2020; **39**(9): e265-e7.
140. Stahl K, Brasen JH, Hoeper MM, David S. Direct evidence of SARS-CoV-2 in gut endothelium. *Intensive Care Med* 2020; **46**(11): 2081-2.
141. Stahl K, Brasen JH, Hoeper MM, David S. Absence of SARS-CoV-2 RNA in COVID-19-associated intestinal endothelialitis. *Intensive Care Med* 2021; **47**(3): 359-60.
142. Steenblock C, Richter S, Berger I, et al. Viral infiltration of pancreatic islets in patients with COVID-19. *Nat Commun* 2021; **12**(1): 3534.
143. Su H, Yang M, Wan C, et al. Renal histopathological analysis of 26 postmortem findings of patients with COVID-19 in China. *Kidney Int* 2020; **98**(1): 219-27.
144. Su H, Gao D, Yang HC, Fogo AB, Nie X, Zhang C. The authors reply. *Kidney Int* 2020; **98**(1): 232-3.
145. Tavazzi G, Pellegrini C, Maurelli M, et al. Myocardial localization of coronavirus in COVID-19 cardiogenic shock. *Eur J Heart Fail* 2020; **22**(5): 911-5.
146. Tchana-Sato V, Ancion A, Tridetti J, et al. Clinical course and challenging management of early COVID-19 infection after heart transplantation: case report of two patients. *BMC Infect Dis* 2021; **21**(1): 89.
147. Tuccari G, Caramori G, Cavallari V, Fadda G, Ieni A. Lungs of COVID-19 affected patients: an autoptic morphological profile. *Atti della Accademia Peloritana dei Pericolanti* 2020; **108**(2): 1-8.
148. Valdespino-Vazquez MY, Helguera-Repetto CA, Leon-Juarez M, et al. Fetal and placental infection with SARS-CoV-2 in early pregnancy. *J Med Virol* 2021; **93**(7): 4480-7.
149. Varga Z, Flammer AJ, Steiger P, et al. Endothelial cell infection and endotheliitis in COVID-19. *Lancet* 2020; **395**(10234): 1417-8.
150. Goldsmith CS, Miller SE, Martines RB, Bullock HA, Zaki SR. Electron microscopy of SARS-CoV-2: a challenging task. *Lancet* 2020; **395**(10238): e99.
151. Varga Z, Flammer AJ, Steiger P, et al. Electron microscopy of SARS-CoV-2: a challenging task - Authors' reply. *Lancet* 2020; **395**(10238): e100.
152. Wang Y, Liu S, Liu H, et al. SARS-CoV-2 infection of the liver directly contributes to hepatic impairment in patients with COVID-19. *J Hepatol* 2020; **73**(4): 807-16.
153. Bangash MN, Patel JM, Parekh D, et al. SARS-CoV-2: Is the liver merely a bystander to severe disease? *J Hepatol* 2020; **73**(4): 995-6.
154. Wang Y, Lu F, Zhao J. Reply to: Correspondence relating to "SARS-CoV-2 infection of the liver directly contributes to hepatic impairment in patients with COVID-19". *J Hepatol* 2020; **73**(4): 996-8.
155. Wang K, Chen W, Zhang Z, et al. CD147-spike protein is a novel route for SARS-CoV-2 infection to host cells. *Signal Transduct Target Ther* 2020; **5**(1): 283.

156. Wu JH, Li X, Huang B, et al. [Pathological changes of fatal coronavirus disease 2019 (COVID-19) in the lungs: report of 10 cases by postmortem needle autopsy]. *Zhonghua Bing Li Xue Za Zhi* 2020; **49**(6): 568-75.
157. Wu J, Yu J, Zhou S, et al. What can we learn from a COVID-19 lung biopsy? *Int J Infect Dis* 2020; **99**: 410-3.
158. Wu MA, Fossali T, Pandolfi L, et al. Hypoalbuminemia in COVID-19: assessing the hypothesis for underlying pulmonary capillary leakage. *J Intern Med* 2021; **289**(6): 861-72.
159. Xiang Q, Feng Z, Diao B, et al. SARS-CoV-2 Induces Lymphocytopenia by Promoting Inflammation and Decimates Secondary Lymphoid Organs. *Front Immunol* 2021; **12**: 661052.
160. Xu X, Chang XN, Pan HX, et al. [Pathological changes of the spleen in ten patients with coronavirus disease 2019(COVID-19) by postmortem needle autopsy]. *Zhonghua Bing Li Xue Za Zhi* 2020; **49**(6): 576-82.
161. Yang Z, Wu J, Ye F, et al. Expert consensus-based laboratory testing of SARS-CoV-2. *J Thorac Dis* 2020; **12**(8): 4378-90.
162. Yao XH, Li TY, He ZC, et al. [A pathological report of three COVID-19 cases by minimal invasive autopsies]. *Zhonghua Bing Li Xue Za Zhi* 2020; **49**(5): 411-7.
163. Yao XH, He ZC, Li TY, et al. Pathological evidence for residual SARS-CoV-2 in pulmonary tissues of a ready-for-discharge patient. *Cell Res* 2020; **30**(6): 541-3.
164. Yao XH, Luo T, Shi Y, et al. A cohort autopsy study defines COVID-19 systemic pathogenesis. *Cell Res* 2021; **31**(8): 836-46.
165. Zhao J, Zhou H, Huang W, et al. Cell morphological analysis of SARS-CoV-2 infection by transmission electron microscopy. *J Thorac Dis* 2020; **12**(8): 4368-73.
166. Zhao CL, Rapkiewicz A, Maghsoodi-Deerwester M, et al. Pathological findings in the postmortem liver of patients with coronavirus disease 2019 (COVID-19). *Hum Pathol* 2021; **109**: 59-68.
167. Martines RB, Ritter JM, Matkovic E, et al. Pathology and Pathogenesis of SARS-CoV-2 Associated with Fatal Coronavirus Disease, United States. *Emerg Infect Dis* 2020; **26**(9): 2005-15.
168. Ritschel N, Radbruch H, Herden C, et al. [COVID-19 and the central and peripheral nervous system]. *Pathologe* 2021; **42**(2): 172-82.
169. Akilesh S, Nicosia RF, Alpers CE, et al. Characterizing Viral Infection by Electron Microscopy: Lessons from the Coronavirus Disease 2019 Pandemic. *Am J Pathol* 2021; **191**(2): 222-7.
170. Bullock HA, Goldsmith CS, Miller SE. Best practices for correctly identifying coronavirus by transmission electron microscopy. *Kidney Int* 2021; **99**(4): 824-7.
171. Cassol CA, Gokden N, Larsen CP, Bourne TD. Appearances Can Be Deceiving - Viral-like Inclusions in COVID-19 Negative Renal Biopsies by Electron Microscopy. *Kidney360* 2020; **1**(8).
172. Frelih M, Erman A, Wechtersbach K, Plesko J, Avsic-Zupanc T, Kojc N. SARS-CoV-2 Virions or Ubiquitous Cell Structures? Actual Dilemma in COVID-19 Era. *Kidney Int Rep* 2020; **5**(9): 1608-10.
173. Neil D, Moran L, Horsfield C, et al. Ultrastructure of cell trafficking pathways and coronavirus: how to recognise the wolf amongst the sheep. *J Pathol* 2020; **252**(4): 346-57.
174. Smith KD, Akilesh S, Alpers CE, Nicosia RF. Am I a coronavirus? *Kidney Int* 2020; **98**(2): 506-7.
